# Individual social contact data reflected SARS-CoV-2 transmission dynamics during the first wave in Germany better than population mobility data – an analysis based on the COVIMOD study

**DOI:** 10.1101/2021.03.24.21254194

**Authors:** Damilola Victoria Tomori, Nicole Rübsamen, Tom Berger, Stefan Scholz, Jasmin Walde, Ian Wittenberg, Berit Lange, Rafael Mikolajczyk, Veronika K Jaeger, André Karch

**Affiliations:** Institute of Epidemiology and Social Medicine, University of Münster, Münster, Germany; Immunization Unit, Robert Koch-Institute, Berlin, Germany; Institute for Medical Epidemiology, Biostatistics and Informatics, University of Halle, Halle, Germany; Department of Epidemiology, Helmholtz Centre for Infection Research, Braunschweig, Germany; German Center for Infection Research, Braunschweig, Germany

**Author notes:** **Corresponding author** Veronika K. Jaeger, PhD, Institute of Epidemiology and Social Medicine University of Münster, Domagkstraße 3, 48149 Münster. contributed equally as last authors (Veronika K Jaeger, André Karch).

**Keywords:** Contact patterns, Contact surveys, SARS-CoV-2, COVID-19, Pandemic

## Abstract

**Background:** The effect of contact reduction measures on infectious disease transmission can only be assessed indirectly and with considerable delay. However, individual social contact data and population mobility data can offer near real-time proxy information.

**Aim:** To compare social contact data and population mobility data with respect to their ability to predict transmission dynamics during the first wave of the SARS-CoV-2 pandemic in Germany.

**Methods:** We quantified the change in social contact patterns derived from self-reported contact survey data collected by the German COVIMOD study from 04/2020-06/2020 (compared to the pre-pandemic period), and estimated the percentage mean reduction in the effective reproduction number R(t) over time. We compared these results to the ones based on R(t) estimates from open-source mobility data and to R(t) values provided by the German Public Health Institute.

**Results:** We observed the largest reduction in social contacts (90%, compared to pre-pandemic data) in late April corresponding to the strictest contacts reduction measures. Thereafter, the reduction in contacts dropped continuously to a minimum of 73% in late June. R(t) estimates based on social contacts underestimated measured R(t) values slightly in the time of strictest contact reduction measures but predicted R(t) well thereafter. R(t) estimates based on mobility data overestimated R(t) considerably throughout the study.

**Conclusions:** R(t) prediction accuracy based on contact survey data was superior to the one based on population mobility data, indicating that measuring changes in mobility alone is not sufficient for understanding changes in transmission dynamics triggered by public health measures.

## Introduction

The role of social contacts for the spread of respiratory infections has been discussed extensively in the year 2020 due to the global outbreak of severe acute respiratory syndrome coronavirus type 2 (SARS-CoV-2) [1,2]. As of March 2021, over 100 million confirmed cases and over 2.5 million deaths have been recorded worldwide [2]. SARS-CoV-2 is primarily transmitted via droplets and aerosols, so that person-to-person contacts are a strong determinant of transmission dynamics [2–4]. Non-pharmaceutical interventions (NPIs) focusing on the reduction of person-to-person contacts are one of the cornerstones of the pandemic response. In the middle of March 2020, Germany mandated school and kindergarten closures, postponed academic semesters, prohibited visiting of nursing homes and restricted the number of people allowed at public and private gatherings in an attempt to protect the vulnerable groups [5]. In the following weeks, contact reduction measures were implemented on a population level by regulating the maximum number of close social contacts outside one’s household and by closing non-essential shops as well as places for leisure activities [5]. After a considerable reduction in reported case numbers, federal governments decided to ease these restrictions gradually starting at the beginning of May 2020.

Social contact patterns are known to be a critical factor for the transmission dynamics of respiratory infections [4,6–10]. However, empirical social contact data have been scarce before the emergence of SARS-CoV-2 [11–13]. One exception is the POLYMOD study, a large-scale survey that described social mixing patterns in eight European countries [12]. In 2005/2006, POLYMOD measured contacts of more than 7,000 participants across eight European countries [12]. Contact patterns observed in POLYMOD have been widely used to parametrize various mathematical models of infectious disease dynamics [3,4,12,14].

During the SARS-CoV-2 pandemic, contact surveys were initiated in several countries to understand the effect of contact precaution measures on social contact patterns [3,4,10,15,16]. While contact surveys offer a direct approach to social contact patterns, they are time- and cost-intensive, and need to be initiated actively. Mobile phone-based mobility data offer a complementary approach to infer changes in contact patterns in a population. Google and Apple granted free access to anonymized mobility data in a global attempt to provide insights into the change of mobility during the pandemic given different physical distancing policies [17,18]. Several SARS-CoV-2 modelling studies assumed that aggregated mobility data can be used as a proxy for the actual number and intensity of contacts of individuals in a defined population, although mobility data measure only certain dimensions of contact behaviour. In this article, we present survey-based social contact data for the first wave of the pandemic in Germany, and assess their ability to predict transmission dynamics (measured by effective reproduction number (R(t)) estimates) when compared to open source population mobility data from Google and Apple [17–19].

## Methods

### Contact surveys

#### Pandemic contact survey – COVIMOD

The contact survey COVIMOD was initiated in April 2020. We commissioned the market research company IPSOS-Mori to conduct the survey based on participants of the online-panel i-say.com. To ensure the samples’ broad representativeness of the German population, participants were recruited by sending email invitations to existing members of the panel based on age, sex, and regional quotas. To gain information on children’s social contacts, a defined subgroup of adult participants with under-aged children (<18 years of age) living in their household were invited to provide information as a proxy for their children. This approach, however, resulted in the sample being no longer representative of the German population as we under-sampled the middle-aged participants who instead filled out the questionnaire for their children. The first COVIMOD survey wave was launched on the 30/04/2020 corresponding to the time of the strictest contact reduction measures in Germany. Survey waves two to four were launched during a time of a gradual easing of the contact reduction measures in May and June 2020. For wave one, a sample of 1.500 participants was commissioned with IPSOS-Mori with an expected response rate of 85% for the next survey waves. Before the launch of survey wave four, the sample was increased by 1,000 additional participants.

The COVIMOD questionnaire includes questions on demographics, current behaviours, attitudes towards SARS-CoV-2, as well as the social contacts of participants. Participants were asked to provide each social contact between 5am the preceding day and 5am the day of the survey, the age and sex of the contact, the duration they spend with each contact, the setting where the contact occurred and if the contact was a household member or not. The questionnaire can be found in Supplementary File 2.

We defined a contact in COVIMOD in line with the POLYMOD study’s definition as “people who you met in person and with whom you exchanged at least a few words, or with whom you had physical contact” [12]. During survey wave one and two, participants were asked to provide each contact separately. Instead of providing each contact one by one, some participants included a group of contacts as one contact (e.g. “customers”). For these groups, we assumed a specific number of social contacts (Supplementary File 3). From survey wave three onwards, participants were offered the opportunity to provide a number of additional contacts (group contacts) they were not able to list individually in case they had too many contacts.

As participants were offered to enter these additional contacts separately, we used different analyses approaches to work with these contacts (scenarios). The main scenario includes all reported contacts plus group contacts weighted for the German population for COVIMOD and POLYMOD. Unweighted results and those without group contacts can be found in the supplementary.

#### Pre-pandemic contact survey – POLYMOD

The European contact survey POLYMOD was used as a baseline pre-pandemic comparison. In Germany, POLYMOD was conducted paper-based with the help of a market research company in 2005/06. Further details about POLYMOD can be found elsewhere [12]. As in COVIMOD, participants in POLYMOD were also allowed to enter the number of additional contacts (group contacts) they had if participants had too many contacts to report them separately.

### Mobility data

We obtained publicly available aggregated mobility data from the Google COVID-19 Community Mobility Reports and from the COVID-19 Apple Mobility trends for the times corresponding to the COVIMOD survey waves [17,18].

Google COVID-19 Community Mobility Reports provide the percentage change in mobility from February 2020 onwards compared to the median of the corresponding weekday between 03/01/2020 and 06/02/2020. Google COVID-19 Community Mobility Reports use aggregated information about true individual movement histories to provide location-specific changes in mobility over time. Data are stratified by the destination of the movement, i.e. retail and recreation, grocery and pharmacy, transit stations, workplace, residential and parks. COVID-19 Apple Mobility trends provide information about the relative volume of requests for directions for all weeks in 2020 compared to a base volume on 13/01/2020.

### Effective reproduction number

R(t) values used in our analysis were obtained from the German Public Health Institute (Robert Koch Institute (RKI)) [19,20]. The 4-day reproduction number calculated by the RKI provides information of the transmission dynamics 8 to 13 days prior [20]. We used the reproduction number provided by the RKI 10 days after the timing of our survey waves.

### Statistical analyses

As the COVIMOD sample is not fully representative of the German population, we used data from the 2011 census to apply survey weights based on the participants’ age, sex, household size and region of residence [21]. The region of residence was not available for POLYMOD, so the POLYMOD data were only weighted according to the participants’ age, sex, and household size using the R package ‘survey’ [22].

We calculated the mean number of social contacts per participant per day, and stratified the social contacts by age group, sex, household size and the day of the week. Additionally, we assessed setting-specific contacts, i.e. home, childcare/school/university, work, public transport, others; childcare/school/university contacts were assessed in the subgroup of participants who reported to attend childcare, school or university, and work contacts were assessed in the subgroup of participants who worked full-or part-time. We calculated symmetric social contact matrices for the age-specific daily frequency of direct social contacts using the ‘socialmixr’ package in R [23]. For the calculation of the contact matrices, participants who reported more than 100 group contacts were excluded from the analysis (COVIMOD: wave three - 6 participants, wave four - 13 participants, POLYMOD: 10 participants).

We assumed that the next-generation matrix for SARS-CoV-2 is a function of the number of age-specific contacts, the duration of infectiousness and the probability that a contact leads to transmission [24]. Hence the basic reproduction number (R0) is proportional to the dominant eigenvalue of the contact matrix [25]. To be able to calculate R(t) as the result of a relative reduction in R0, we assumed that the social contact patterns before the implementation of the contact reduction measures were similar to the POLYMOD contact patterns and that the duration of infectiousness and the per-contact transmission probability remained constant. Additionally, we assumed that the transmission probability did not depend on age. Under these assumptions, the relative reduction of R(t) from R0 is equivalent to the reduction in the contact matrices’ dominant eigenvalue allowing us to estimate the reproduction number corresponding to contacts recorded in COVIMOD. We assumed R0 to follow a normal distribution with mean 2.6 and standard deviation 0.54 [3]. We drew 10,000 bootstrap samples from POLYMOD and COVIMOD to assess uncertainty.

To assess mobility, we averaged across all the movement types for the Google mobility data and for the Apple mobility data except for the movements to parks as this is expected to vary tremendously during the seasons. We only assessed mobility data for the same time intervals as the COVIMOD waves’ timings. We calculated the mean percentage change within the time interval as well as the minimum and maximum for each time interval of the COVIMOD waves for Google and Apple mobility.

Similarly, we calculated the mean and the minimum and maximum of the RKI R(t) estimates for the corresponding time intervals.

R version 4.0.2 was used for all analyses [26].

## Ethics approval

COVIMOD was approved by the ethics committee of the Medical Board Westfalen-Lippe and the University of Münster (2020-473-f-s). Informed consent was obtained from all COVIMOD participants. As only anonymised POLYMOD data were used in this study, institutional review was not required for reanalysis [12].

## Results

### Participant characteristics of POLYMOD and COVIMOD

During POLYMOD, 1,341 participants were surveyed in Germany; they recorded a total of 22,814 contacts. In the first COVIMOD wave, we surveyed 1,560 participants who recorded a total of 3,256 social contacts; this changed to 1,356 participants with a total of 4,852 contacts in the second survey wave, 1,081 participants with a total of 6,311 in the third wave, and 1,890 participants with a total of 13,346 contacts in the fourth wave.

The youngest participants in all COVIMOD waves were younger than one (the parents were surveyed as a proxy), and the oldest was 91 years of age. Between 47% and 50% of all COVIMOD participants were female (Table 1). In POLYMOD and all COVIMOD waves, the median household size of the participants was 3 (POLYMOD IQR 2-4, COVIMOD wave one IQR 2-4, wave two IQR 2-3, wave three IQR 2-3, wave four IQR 1-3). In COVIMOD survey waves one, two and three, most participants reported their social contacts on a Thursday, whereas in wave four, most contacts were reported on a Monday; less than a quarter of participants reported the contacts during the weekend (Table 1).

**Table 1.**
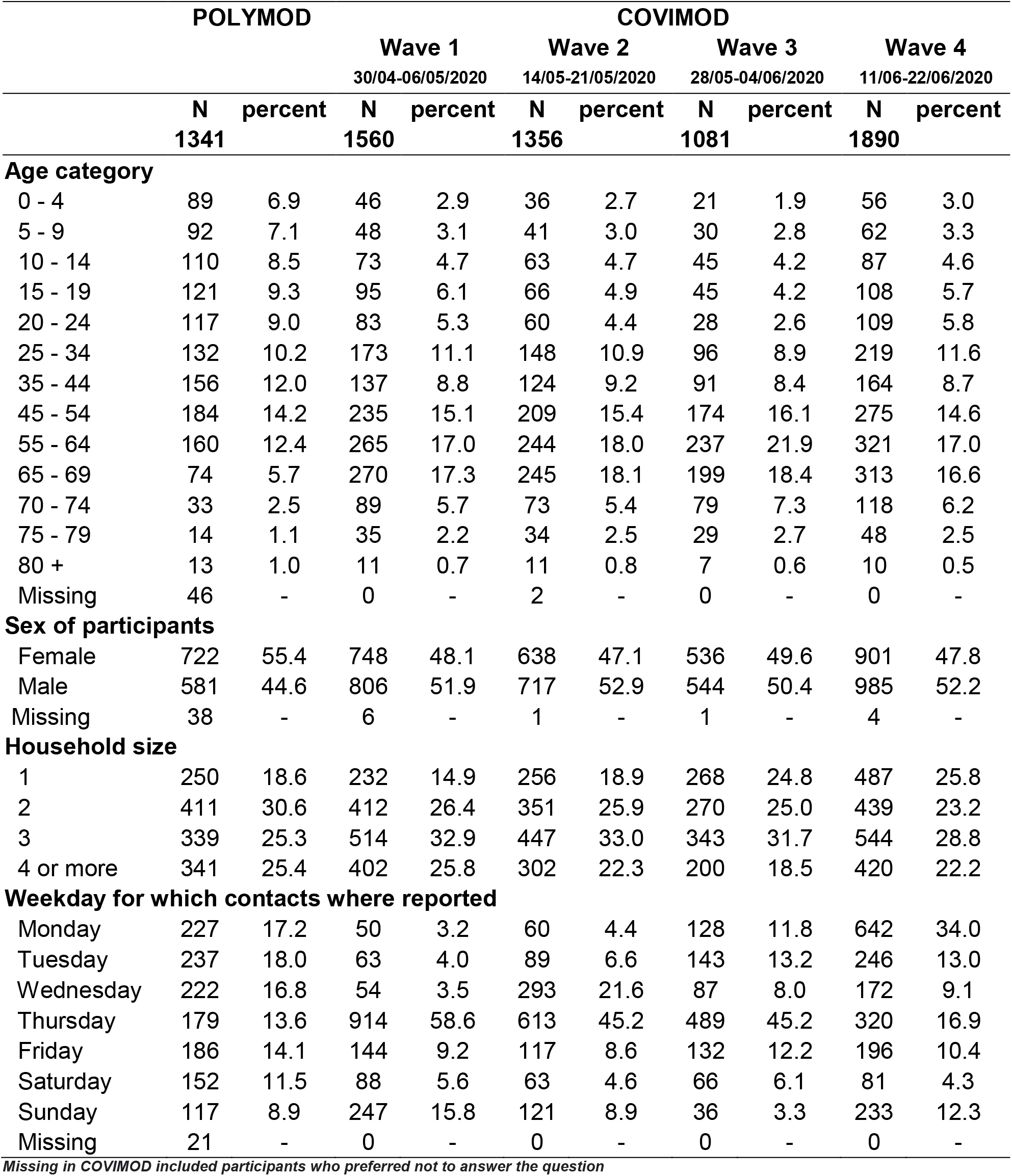
Participant characteristics in the COVIMOD survey waves one to four compared to the POLYMOD survey.

A comparison of the characteristics of German population and the POLYMOD and COVIMOD participants can be found in Supplementary File 1, Table 1. The participant characteristics after weighting can be found in Supplementary File 1, Table 1.1a. The analyses hereafter are based on the weighted data including group contacts.

### Number of social contacts

The mean number of contacts measured per participant during all COVIMOD waves (wave one, 1.9 contacts (SD 1.8), wave two, 3.5 contacts (SD 6.1), wave three, 6.5 contacts (SD 21.6), wave four, 7.3 contacts (SD 29.6)) was considerably lower in comparison to the 19.6 contacts (SD 27.7) measured in POLYMOD pre-pandemically (Figure 1a; Supplementary File 1 Table 1.2a). The reduction in the number of overall contacts between POLYMOD and COVIMOD was consistent across age, sex, household size and weekday (Figure 1; Supplementary File 1 Table 1.2a). The patterns and relative reductions observed for physical contacts (skin-to-skin) measured during COVIMOD was generally in line with those observed for all contacts (Supplementary File 1 Table 1.2b, Figure 1.5b).

**Figure 1.**
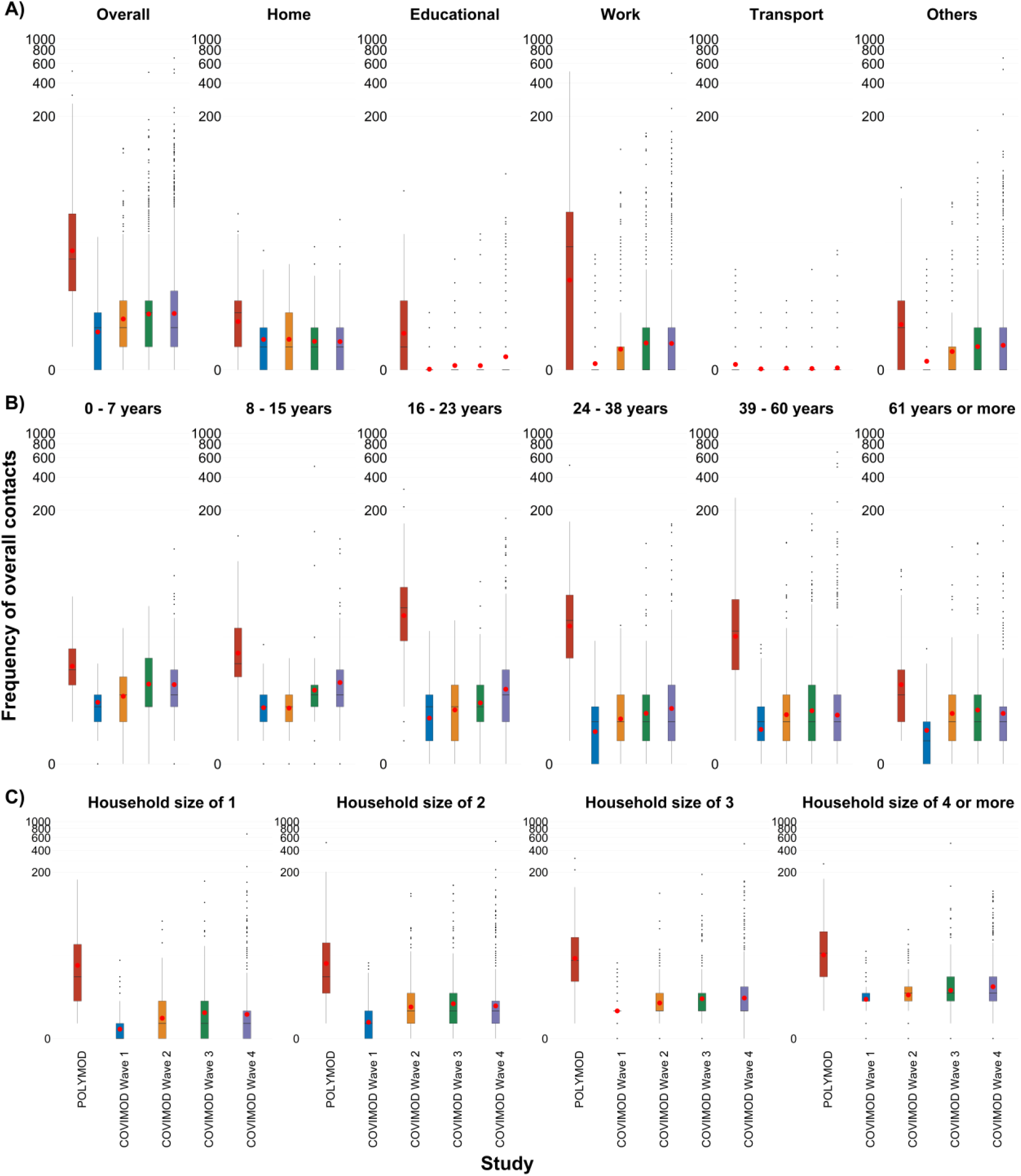
The number of all contacts during the POLYMOD and COVIMOD survey waves one to four in various settings. In the top row, the number of (A) overall contacts, (B) home contacts, (C) work contacts, (D) educational contacts, (E) public transport contacts and (F) other contacts are displayed. In the middle row, the overall number of contacts stratified by age are displayed. In the bottom row, the overall number of contacts stratified by household size are displayed. Boxes represent the 25^th^, 50^th^ and 75^th^ percentiles, the whiskers represent the 10^th^ and 90^th^ percentile. The red dot marks the mean. **Note:** The displayed educational contacts are based on the group of participants who attended an educational facility (kindergarten, school, university) and work contacts are based on the group of participants who reported to work full-/part-time. Participants with no contacts are displayed as 0 on the log-scale of the y-axis.

While the mean number of home contacts was stable across all COVIMOD waves (and just a little bit lower than in POLYMOD, Figure 1, Supplementary File 1 Table 1.2a), contacts at work and in educational settings were dramatically reduced during the first COVIMOD wave. Contacts at work increased gradually thereafter but remained much lower than in POLYMOD even for survey wave four; educational contacts started to increase only at survey wave four as schools were closed before (Figure 1, Supplementary File 1 Table 1.2c). Moreover, the distribution of contacts observed changed considerably over the different COVIMOD waves. While the maximum number of contacts reported overall and in specific settings was clearly reduced in the first COVIMOD wave, it approximated the one reported in POLYMOD already in wave two and three and reached it in wave four (although mean and median contact numbers were still clearly reduced). The number of contacts in different settings for the unweighted analyses can be found in Supplementary File 1 Tables 2.2c, 3.2c and 4.2c.

Participants in the age group 0 to 19 years reported the highest mean number of skin-to-skin contacts compared to the other age groups for POLYMOD and all COVIMOD waves (Supplementary File 1 Figure 1.5b). A similar trend was also observed in participants with a household size of more than four (Supplementary File 1 Figure 1.5c).

POLYMOD and COVIMOD participants in all age groups shared the majority of their contacts with individuals of similar age, demonstrating the expected age-assortative pattern (Figure 2; Supplementary File 1 Figures 1.6a). Since contact matrices derived from the first two COVIMOD waves were dominated by contacts at home, they reveal mainly contacts with life partners and children. This changed slowly through survey waves three and four due to the gradual increase in work and leisure time (“other”) contacts, which resulted in a broader distribution of the age of potential contact persons.

**Figure 2.**
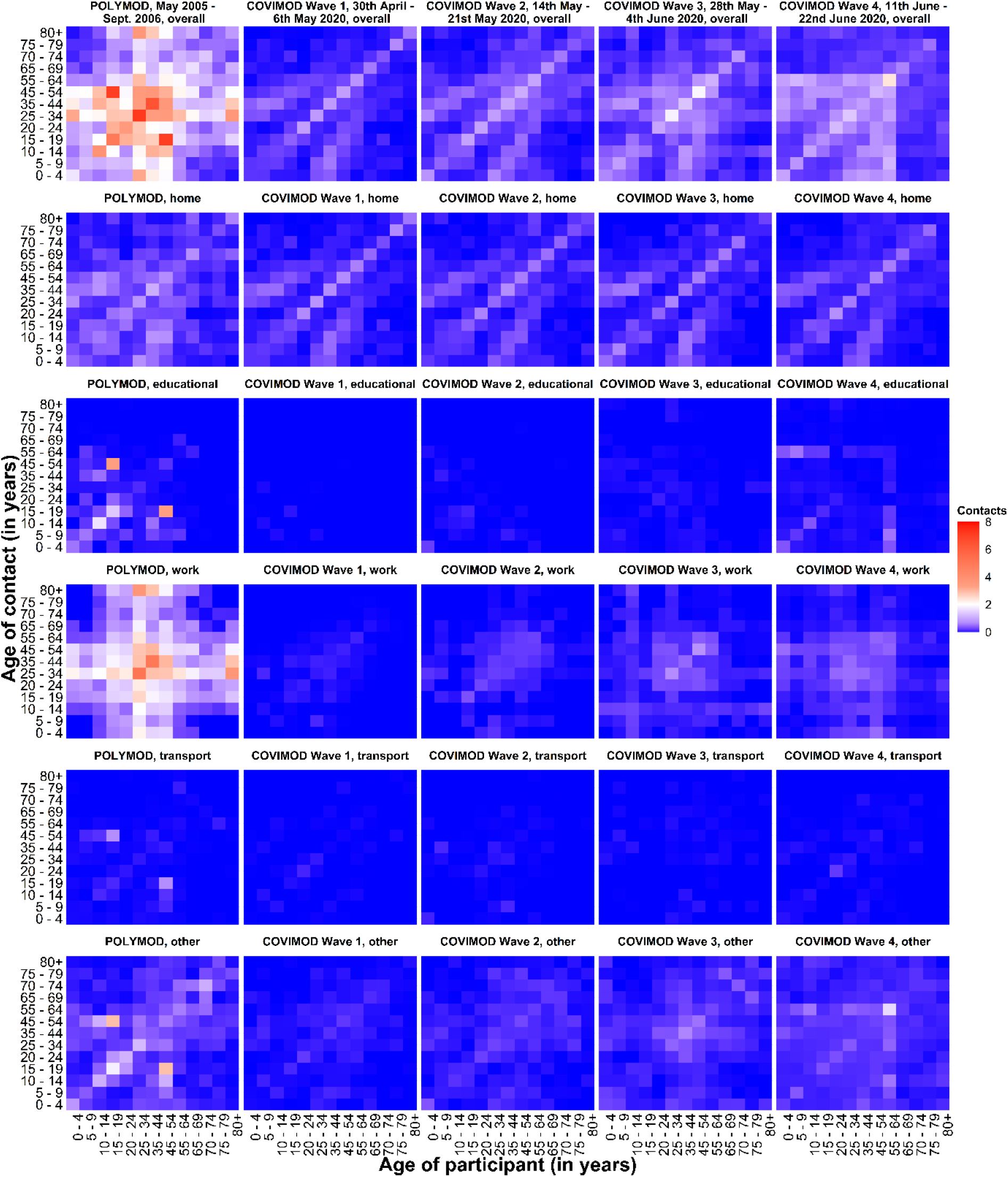
Social contact matrices with the mean total number of reported daily social contacts by participants in different age groups with individuals in other age groups in POLYMOD and COVIMOD survey waves one to four in various settings. Displayed are (A) the overall number of contacts, (B) home contacts, (C) educational contacts, (D) work contacts, (E) public transport contacts and (F) other contacts.

### Prediction of effective reproduction numbers based on contact and mobility data

In all COVIMOD waves, the mean R(t) estimated based on COVIMOD data was smaller than 1 (representing a percentage mean reduction in contacts of at least 70%); we observed the highest percentage mean reduction with 90% at the end of April (survey wave one), which corresponds to the time of the strictest contact reduction measures. Subsequently, the mean reduction decreased with time as the contact reduction measures were loosened (wave two: 86%, wave three: 80%, wave four: 73%; Figure 3; Supplementary File 1 Table 1.3). Compared to the R(t) values reported by the RKI, R(t) estimates based on COVIMOD were lower during the first survey wave, but fit quite well during wave two to four. R(t) estimates based on Google and Apple data were considerably higher than RKI R(t) estimates throughout the entire study (Figure 3; Supplementary File 1 Table 1.3). Both mobility data sources found mobility patterns similar to pre-pandemic data already during the time of survey wave one and two, while true R(t) values were still considerably below 1.

**Figure 3.**
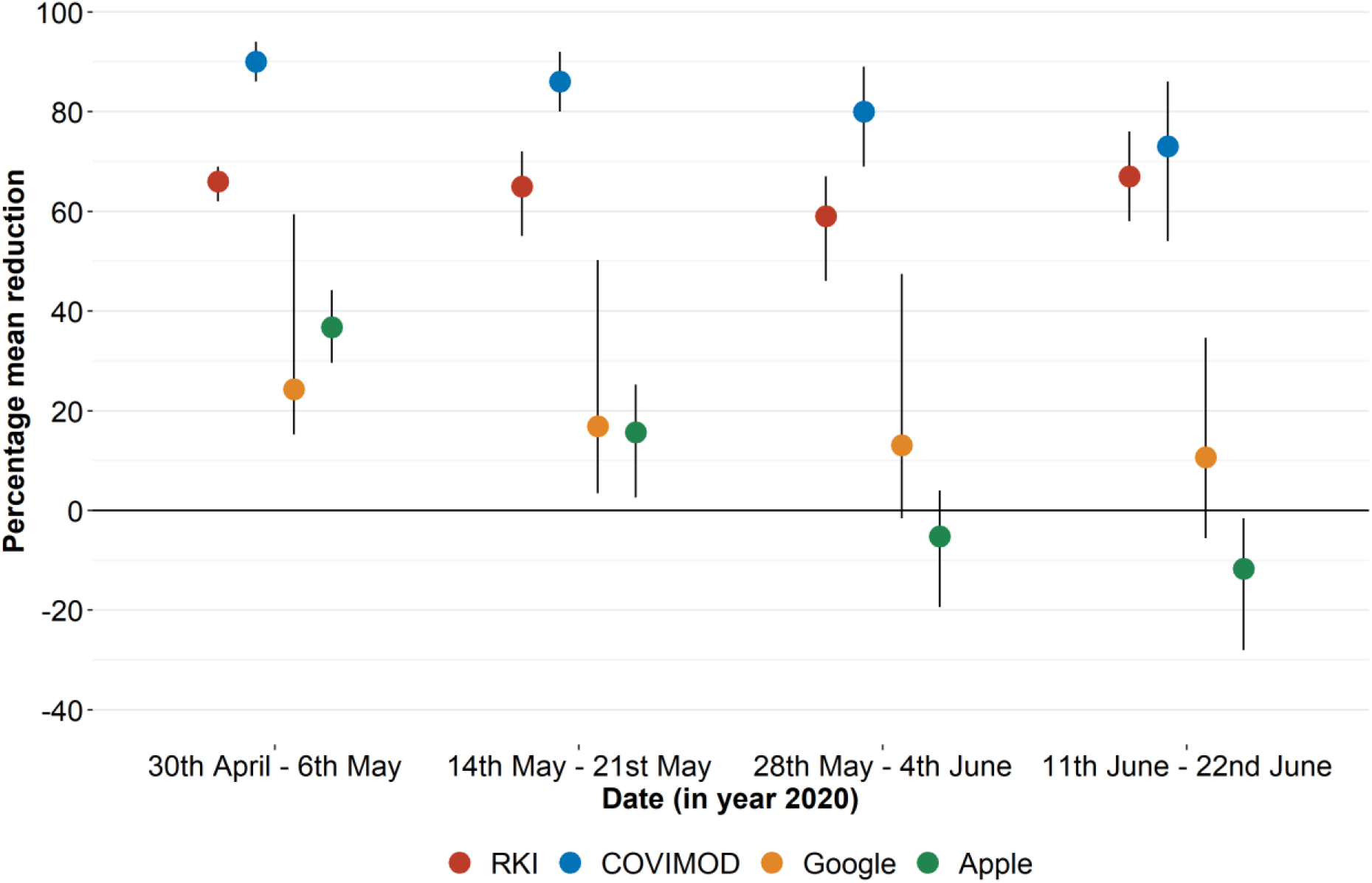
Comparison of R(t) estimates obtained based on different input data. Measured effective reproduction number at the timing of COVIMOD survey waves 1 to 4 (red), estimated reproduction number based on the reduction of social contacts at the times of COVIMOD survey waves one to four and the reduction in mobility at the times of the COVIMOD survey waves (yellow and green). Displayed are: RKI - the percentage mean and minimum and maximum reductions in the reproduction number in the assessed time intervals, COVIMOD - the calculated percentage mean reductions in the reproduction number and the 95% confidence interval, Google and Apple mobility – the percentage mean and minimum and maximum reduction (indicated by lines) in mobility in the assessed time intervals.

## Discussion

We showed in this study that non-pharmaceutical interventions introduced in Germany during the first wave of the SARS-CoV-2 pandemic were associated with a considerable reduction in social contacts reported in contact surveys. We quantified the relative reduction in contacts based on contact survey data and publicly available mobility data and found that changes in contact patterns measured in survey data predicted transmission dynamics (measured as R(t)) better than changes measured in aggregated mobility data. This indicates that deriving contact behaviour from mobility data alone is not suitable for making real-time inference on the effects of public health measures on the transmission dynamics in a population. Mobility data used in this study suggested that contact behaviour went back to normal almost instantly after the contact reduction measures were relaxed, which did not reflect the observed R(t) values. A reason for that might be that people still tried to minimise close contacts outside their own households, and maximised distance to the contacts they had, although their mobility, e.g. back to work, already reached almost pre-pandemic levels.

Consistent with studies from other European countries, we found a 73% mean reduction in contacts across the first four waves of COVIMOD [3,4]. Even though the reported number of daily contacts increased over the survey waves, it was still considerably lower than in POLYMOD, indicating sustainable behaviour change even after the end of the strictest contact reduction measures. We found an increased variance in the reported daily number of contacts as the COVIMOD waves progressed, with the maximum number of contacts increasing from 16 in survey wave one to 674 in wave four, while median contact numbers were not affected similarly. Since SARS-CoV-2 has been shown to be associated with a high variance in the number of transmissions arising from one infectious individual [27], this sharp increase in the maximum number of contacts has huge implications for the risk of superspreading events as the direct aftermath of the end of public health interventions. Participants aged 60 and above reported fewer contacts in all COVIMOD waves as well as a larger reduction to pre-pandemic values when compared to children and middle-aged persons. This should be taken into account when assessing the effects of vaccination prioritization strategies in combination with NPIs, as people in this age group are known to be more vulnerable to COVID-19 infections [15].

We further observed a smaller and more stable reduction in home contacts than in work, educational and leisure time contacts, which confirms the assertion that reduction in contacts is location-specific [3]. This is reasonable as most of the social distancing measures implemented had their main impact outside the household. We confirmed that the majority of remaining contacts under strict contact reduction measures happens between life partners and parents and children, which mirrors the huge role of this transmission setting under contact reduction measures [7,28].

Our study has several limitations. COVIMOD data are not fully representative of the German population since some adult participants with under-aged children living in their household were invited to provide information as a proxy for their children. Moreover, the elderly (>70 years) and the very young (<10 years of age) are underrepresented in COVIMOD. We tried to correct for that by introducing weights for sex, age, and household size; however, there were no relevant differences in the results of the unweighted and weighted analyses. Participants in COVIMOD were asked to record their contacts retrospectively so that different forms of information bias could have been introduced. For example, it might be challenging to remember a higher number of contacts, or the participants’ willingness to report high numbers of contacts individually might be lower as this is quite tedious and time-consuming. We tried to minimise this by allowing the participants to record group contacts. We also cannot rule out that COVIMOD attracted specifically participants who adhered to social distancing rules as these individuals might be more likely to respond to health surveys. This could have led to an overestimation of the relative reduction of contacts, and could explain the gap between R(t) values based on case numbers and R(t) values estimated from contact data. We tried to minimise this bias by using an established online panel not focusing on healthcare questions as the platform for COVIMOD. Even though contact-related questions were similarly phrased between POLYMOD and COVIMOD, POLYMOD was paper-based, and COVIMOD surveys were web-based. Previous research suggested that participants might report more contacts in paper-based surveys than in web-based surveys [11,29]. Future research will be conducted on the differences between web- and paper-based contacts during the pandemic. However, our findings are consistent with other studies that examined social contact patterns under strict contact reduction measures [3,4,15,30]. We used aggregated mobility data in our study that were freely available and have been discussed as a potential real-time proxy for SARS-CoV-2 transmission dynamics. Although we took advantage of two different data sources representing complementary ways to define mobility, our results cannot be automatically generalized to other ways of measuring mobility (e.g. based on individual movement patterns).

In summary, our study provides a comprehensive quantification of social contacts and mixing patterns relevant to the spread of SARS-CoV-2 during spring and summer 2020 in Germany. Our results indicate that population-based contact surveys provide a suitable platform for near real-time assessment of transmission dynamics for respiratory infections in a population. However, this requires considerable financial and computational efforts. Aggregated mobility data as a proxy for contacts did not show the same degree of persistent reduction, and consequently did not reflect the development of effective reproduction numbers.

## Data Availability

The data are available from the corresponding author upon valid scientific request.

## Data sharing

The data are available from the corresponding author upon valid scientific request.

## Authors’ contributions

DVT, VKJ, RTM and AK designed the study, DVT, NR and VKJ conducted the statistical analyses, DVT, VKJ and AK wrote the manuscript. All authors interpreted the study findings, contributed to the manuscript, and approved the final version of the manuscript.

## Ethics approval

COVIMOD was approved by the ethics committee of the Medical Board Westfalen-Lippe and the University of Münster, reference number 2020-473-f-s. Informed consent was obtained from all study participants of the COVIMOD study. The POLYMOD data collection was approved by national institutional review boards [12]. As only anonymised POLYMOD data were used in this study, institutional review was not required for reanalysis.

### Appendices

Supplementary File 1. Additional results. Supplementary File 2. COVIMOD questionnaire.

Supplementary File 3. Consideration of additional contacts.

Supplementary File 4. Contact matrices

