## Supplementary File 1 for "Individual social contact data reflected SARS-CoV-2 transmission dynamics during the first wave in Germany better than population mobility data – an analysis based on the COVIMOD study"


**Participant characteristics in the COVIMOD survey Waves 1 to 4 and in the POLYMOD survey, in comparison with the German population** ... 2

1. **WEIGHTED ANALYSIS INCLUDING GROUP CONTACTS** ... 3

1.1. **Participant characteristics.** ... 3

1.2. **Number of social contacts.** ... 4

1.3. **Reproduction number estimates of SARS-CoV-2 under contact reduction measures.** ... 7

1.4. **Boxplots of the number of overall contacts during the POLYMOD and COVIMOD survey Waves 1 to 4 in various settings.** ... 8

1.5. **Boxplots of the number of physical contacts during the POLYMOD and COVIMOD survey Waves 1 to 4 in various settings.** ... 10

1.6. **Social contact patterns**. ... 15

1.7. **Estimated reproduction number of SARS-CoV-2 under contact reduction measures.** ... 16

2. **UNWEIGHTED ANALYSIS INCLUDING GROUP CONTACTS** ... 17

2.1. **Participant characteristics.** ... 17

2.2. **Number of social contacts.** ... 18

2.3. **Reproduction number estimates of SARS-CoV-2 under contact reduction measures** ... 21

2.4. **Boxplots of the number of overall contacts during the POLYMOD and COVIMOD survey Waves 1 to 4 in various settings.** ... 22

2.5. **Boxplots of the number of physical contacts during the POLYMOD and COVIMOD survey waves 1 to 4 in various settings.** ... 27

2.6. **Social contact patterns.** ... 32

2.7. **Estimated reproduction number of SARS-CoV-2 under contact reduction measures.** ... 34

3. **WEIGHTED ANALYSIS WITHOUT GROUP CONTACTS** ... 35

3.2. **Number of social contacts.** ... 35

3.3. **Reproduction number estimates of SARS-CoV-2 under contact reduction measures.** ... 38

3.4. **Boxplots of the number of overall contacts during the POLYMOD and COVIMOD survey waves 1 to 4 in various settings.** ... 39

3.5. **Boxplots of the number of physical contacts during the POLYMOD and COVIMOD survey waves 1 to 4 in various settings.** ... 44

3.6. **Social contact patterns.** ... 49

3.7. **Estimated reproduction number of SARS-CoV-2 under contact reduction measures.** ... 51

4. **UNWEIGHTED ANALYSIS INCLUDING GROUP CONTACTS** ... 52

4.2. **Number of social contacts.** ... 52

4.3. **Reproduction number estimates of SARS-CoV-2 under contact reduction measures.** ... 55

4.4. **Boxplots of the number of overall contacts during the POLYMOD and COVIMOD survey waves 1 to 4 in various settings.** ... 56

4.5. **Boxplots of the number of physical contacts during the POLYMOD and COVIMOD survey waves 1 to 4 in various settings.** ... 61

4.6. **Social contact patterns.** ... 66

4.7. **Estimated reproduction number of SARS-CoV-2 under contact reduction measures.** ... 68

**Table 1**. Participant characteristics in the COVIMOD survey Waves 1 to 4 and in the POLYMOD survey, in comparison with the German population (https://www.zensus2011.de/DE/Zensus2011/zensus2011_node.html)

| | German population | POLYMOD | COVIMOD | | | |
|---|---|---|---|---|---|---|
| | | | Wave 1 | Wave 2 | Wave 3 | Wave 4 |
| **Age category** | | | | | | |
| 0 - 9 | 8.6% | 14.0% | 6.0% | 5.7% | 4.7% | 6.3% |
| 10 - 19 | 9.9% | 17.8% | 10.8% | 9.6% | 8.4% | 10.3% |
| 20 - 29 | 12.1% | 14.8% | 9.6% | 7.9% | 6.2% | 11.0% |
| 30 - 39 | 11.8% | 9.7% | 11.8% | 12.0% | 9.3% | 10.4% |
| 40 - 49 | 16.6% | 14.0% | 8.2% | 9.1% | 8.8% | 8.9% |
| 50 - 59 | 14.5% | 12.8% | 19.7% | 20.2% | 22.2% | 18.6% |
| 60 - 69 | 11.1% | 12.2% | 25.3% | 26.7% | 29.6% | 25.2% |
| 70 - 79 | 10.1% | 3.6% | 7.9% | 7.9% | 10.0% | 8.8% |
| 80 + | 5.3% | 1.0% | 0.7% | 0.8% | 0.7% | 0.5% |
| **Sex of participants** | | | | | | |
| Female | 48.8% | 55.4% | 48.1% | 47.1% | 49.6% | 47.8% |
| Male | 51.2% | 44.6% | 51.9% | 52.9% | 50.4% | 52.2% |
| **Household size** | | | | | | |
| 1 | 17.1% | 18.6% | 14.9% | 18.9% | 24.8% | 25.8% |
| 2 | 31.1% | 30.6% | 26.4% | 25.9% | 25.0% | 23.2% |
| 3 | 20.1% | 25.3% | 32.9% | 33.0% | 31.7% | 28.8% |
| 4 | 19.4% | 18.2% | 12.8% | 11.1% | 9.9% | 11.3% |
| 5 | 7.6% | 4.8% | 9.6% | 8.3% | 6.6% | 8.1% |
| 6 or more | 4.7% | 2.4% | 3.4% | 2.9% | 2.0% | 2.9% |

### 1. WEIGHTED ANALYSIS INCLUDING GROUP CONTACTS

**COVIMOD Wave 3 - 4 include group contacts in home, work, and educational settings while POLYMOD includes group contacts in work setting.**

#### 1.1. Participant characteristics.

1.1.a. Table 1.1a. Participant characteristics in the COVIMOD survey Waves 1 to 4 and in the POLYMOD survey for the weighted analysis including group contacts.

| | POLYMOD | COVIMOD | | | |
|---|---|---|---|---|---|
| | | Wave 1 | Wave 2 | Wave 3 | Wave 4 |
| | percent | percent | percent | percent | percent |
| **Age category** | | | | | |
| 0 - 4 | 4.1 | 3.2 | 3.2 | 2.3 | 3.7 |
| 5 - 9 | 4.9 | 3.4 | 4.1 | 3.9 | 4.4 |
| 10 - 14 | 5.4 | 3.8 | 4.1 | 4.3 | 5.0 |
| 15 - 19 | 4.9 | 6.7 | 5.5 | 5.1 | 5.6 |
| 20 - 24 | 8.1 | 6.4 | 5.2 | 3.3 | 5.7 |
| 25 - 34 | 9.9 | 12.7 | 13.0 | 14.2 | 14.1 |
| 35 - 44 | 14.4 | 12.9 | 12.6 | 11.5 | 11.4 |
| 45 - 54 | 17.6 | 17.8 | 18.0 | 16.2 | 16.6 |
| 55 - 64 | 11.7 | 11.8 | 12.7 | 16.1 | 12.3 |
| 65 - 69 | 5.4 | 10.1 | 10.1 | 10.4 | 9.2 |
| 70 - 74 | 6.9 | 7.0 | 6.9 | 8.7 | 7.1 |
| 75 - 79 | 2.6 | 2.9 | 3.3 | 2.8 | 3.5 |
| 80 + | 4.1 | 1.3 | 1.4 | 1.3 | 1.4 |
| **Sex of participants** | | | | | |
| Female | 51.1 | 50.5 | 49.9 | 51.4 | 49.5 |
| Male | 48.9 | 49.5 | 50.1 | 48.6 | 50.5 |
| **Household size** | | | | | |
| 1 | 17.6 | 15.1 | 16.6 | 18.8 | 18.2 |
| 2 | 32.1 | 35.0 | 34.2 | 36.2 | 33.8 |
| 3 | 20.0 | 22.8 | 22.7 | 22.2 | 21.6 |
| 4 or more | 30.2 | 27.1 | 26.5 | 22.8 | 26.4 |
| **Weekdays** | | | | | |
| Monday | 17.3 | 3.7 | 4.3 | 12.7 | 35.3 |
| Tuesday | 16.9 | 4.4 | 7.7 | 12.6 | 14.1 |
| Wednesday | 16.5 | 3.4 | 18.2 | 7.6 | 10.6 |
| Thursday | 14.9 | 57.1 | 48.0 | 43.4 | 14.9 |
| Friday | 13.6 | 7.6 | 8.4 | 12.7 | 8.5 |
| Saturday | 11.8 | 5.9 | 4.6 | 6.7 | 4.4 |
| Sunday | 9.0 | 17.9 | 8.7 | 4.2 | 12.2 |

### 1.2. Number of social contacts.

1.2.a. Table 1.2a. Number of recorded overall contacts per participant per day stratified by age, gender, household size, and day of the week in the COVIMOD survey Waves 1 to 4 and the POLYMOD survey for the weighted analysis including group contacts.

| | **POLYMOD** | | **COVIMOD** | | | | | | | |
|---|---|---|---|---|---|---|---|---|---|---|
| | | | **Wave1** | | **Wave 2** | | **Wave 3** | | **Wave 4** | |
| | **Mean (SD)** | **Min, Max** | **Mean (SD)** | **Min, Max** | **Mean (SD)** | **Min, Max** | **Mean (SD)** | **Min, Max** | **Mean (SD)** | **Min, Max** |
| | **19.6 (27.7)** | **1,512** | **1.9 (1.8)** | **0, 16** | **3.5 (6.1)** | **0,102** | **6.5 (21.6)** | **0,500** | **7.3 (29.6)** | **0,674** |
| **Age category** | | | | | | | | | | |
| 0 - 4 | 9.4 (6.2) | 2,30 | 3.1 (1.7) | 0,8 | 4.6 (2.5) | 0,14 | 6.6 (6.2) | 2,24 | 7.7 (11.6) | 0,89 |
| 5 - 9 | 8.5 (6.3) | 2,43 | 3.4 (1.7) | 0,8 | 3.8 (3.1) | 0,17 | 5.9 (5.8) | 0,27 | 6.0 (5.3) | 0,31 |
| 10 - 14 | 14.4 (13.9) | 2,61 | 2.9 (1.5) | 0,12 | 3.2 (2.0) | 0,9 | 18.3 (71.7) | 0,500 | 9.4 (17.1) | 0,110 |
| 15 - 19 | 31.8 (26.0) | 1,215 | 3.3 (1.9) | 0,8 | 4.3 (3.9) | 0,20 | 4.5 (5.3) | 0,27 | 9.3 (16.7) | 0,113 |
| 20 - 24 | 29.1 (34.0) | 1,310 | 1.6 (2.1) | 0,16 | 3.3 (3.1) | 0,20 | 5.8 (8.2) | 0,45 | 8.9 (20.8) | 0,169 |
| 25 - 34 | 31.3 (47.1) | 2,512 | 1.7 (1.7) | 0,13 | 3.0 (3.3) | 0,18 | 5.8 (13.2) | 0,100 | 6.7 (15.9) | 0,150 |
| 35 - 44 | 24.7 (20.6) | 1,111 | 2.0 (2.0) | 0,11 | 3.3 (3.7) | 0,33 | 8.3 (22.9) | 0,150 | 6.8 (41.7) | 0,674 |
| 45 - 54 | 26.1 (33.5) | 1,260 | 1.7 (1.6) | 0,9 | 3.7 (9.7) | 0,102 | 8.0 (22.2) | 0,131 | 11.9 (55.7) | 0,532 |
| 55 - 64 | 14.7 (21.2) | 1,150 | 1.7 (1.6) | 0,12 | 3.9 (5.5) | 0,42 | 6.0 (18.3) | 0,186 | 6.8 (17.8) | 0,127 |
| 65 - 69 | 8.6 (9.9) | 1,58 | 1.4 (1.6) | 0,11 | 4.3 (9.9) | 0,93 | 5.2 (13.3) | 0,101 | 4.6 (14.9) | 0,216 |
| 70 - 74 | 5.6 (5.2) | 1,25 | 1.2 (1.6) | 0,8 | 2.7 (3.3) | 0,25 | 2.7 (1.9) | 0,13 | 2.6 (3.4) | 0,37 |
| 75 - 79 | 4.8 (4.0) | 1,14 | 1.5 (1.2) | 0,5 | 2.6 (1.8) | 0,9 | 3.4 (4.2) | 0,32 | 2.2 (1.9) | 0,17 |
| 80 + | 3.9 (1.9) | 2,8 | 0.7 (1.0) | 0,3 | 1.2 (2.3) | 0,9 | 0.5 (1.3) | 0,7 | 0.5 (1.1) | 0,4 |
| **Sex of participants** | | | | | | | | | | |
| Female | 20.6 (31.3) | 1,512 | 1.9 (1.8) | 0,16 | 3.6 (6.6) | 0,102 | 6.8 (19.0) | 0,186 | 6.9 (29.8) | 0,493 |
| Male | 18.6 (23.3) | 1,310 | 1.9 (1.8) | 0,12 | 3.4 (5.6) | 0,93 | 6.2 (24.1) | 0,500 | 7.6 (29.4) | 0,674 |
| **Household size** | | | | | | | | | | |
| 1 | 16.0 (21.8) | 1,158 | 0.7 (1.4) | 0,12 | 1.9 (3.5) | 0,42 | 3.3 (10.9) | 0,150 | 6.1 (36.3) | 0,674 |
| 2 | 17.1 (30.7) | 1,512 | 1.3 (1.5) | 0,11 | 3.7 (8.3) | 0,101 | 7.5 (21.5) | 0,132 | 5.4 (23.3) | 0,532 |
| 3 | 20.9 (28.5) | 1,310 | 2.1 (1.4) | 0,11 | 3.5 (6.1) | 0,102 | 5.1 (12.9) | 0,186 | 9.5 (42.4) | 0,493 |
| 4 or more | 23.4 (26.3) | 2,260 | 3.3 (1.7) | 0,16 | 4.3 (3.1) | 0,32 | 9.0 (32.3) | 0,500 | 8.6 (15.1) | 0,109 |
| **Weekdays** | | | | | | | | | | |
| Monday | 18.0 (17.2) | 1,124 | 2.4 (1.7) | 0,6 | 4.1 (4.3) | 0,24 | 5.6 (12.0) | 0,60 | 8.2 (32.9) | 0,674 |
| Tuesday | 21.6 (43.6) | 1,512 | 2.5 (2.2) | 0,8 | 4.7 (9.3) | 0,102 | 4.8 (5.5) | 0,32 | 12.8 (51.6) | 0,493 |
| Wednesday | 18.5 (24.4) | 1,158 | 2.5 (2.7) | 0,13 | 3.2 (4.2) | 0,42 | 7.0 (14.8) | 0,80 | 7.2 (16.4) | 0,109 |
| Thursday | 18.3 (26.9) | 1,260 | 1.9 (1.8) | 0,16 | 3.4 (5.4) | 0,93 | 7.2 (20.8) | 0,186 | 4.2 (13.3) | 0,216 |
| Friday | 21.5 (25.0) | 1,162 | 1.9 (1.8) | 0,8 | 4.2 (4.4) | 0,32 | 9.3 (43.0) | 0,500 | 5.6 (19.6) | 0,238 |
| Saturday | 18.3 (21.7) | 1,215 | 1.7 (1.5) | 0,8 | 5.0 (14.8) | 0,101 | 2.9 (3.3) | 0,33 | 4.4 (8.6) | 0,95 |
| Sunday | 20.7 (22.0) | 1,96 | 1.8 (1.6) | 0,12 | 2.3 (2.5) | 0,21 | 4.1 (10.4) | 0,99 | 4.1 (13.4) | 0,126 |

1.2.b. Table 1.2b. Number of recorded physical contacts per participant per day stratified by age, gender, household size, and day of the week in the COVIMOD survey Waves 1 to 4 and the POLYMOD survey for the weighted analysis including group contacts.

| | **POLYMOD** | | **COVIMOD** | | | | | | | |
|---|---|---|---|---|---|---|---|---|---|---|
| | | | **Wave1** | | **Wave 2** | | **Wave 3** | | **Wave 4** | |
| | **Mean (SD)** | **Min, Max** | **Mean (SD)** | **Min, Max** | **Mean (SD)** | **Min, Max** | **Mean (SD)** | **Min, Max** | **Mean (SD)** | **Min, Max** |
| | **5.3 (4.7)** | **0,45** | **0.8 (1.2)** | **0,11** | **1.1 (3.7)** | **0,101** | **1.6 (4.7)** | **0,129** | **2.0 (14.5)** | **0,674** |
| **Age category** | | | | | | | | | | |
| 0 - 4 | 7.5 (4.9) | 0,30 | 2.7 (1.7) | 0,7 | 3.3 (2.3) | 0,8 | 4.8 (5.1) | 0,20 | 5.3 (9.0) | 0,51 |
| 5 - 9 | 6.3 (4.8) | 0,33 | 2.2 (1.7) | 0,6 | 2.2 (1.8) | 0,6 | 4.0 (4.5) | 0,22 | 3.1 (2.8) | 0,23 |
| 10 - 14 | 6.5 (4.9) | 0,25 | 1.9 (1.5) | 0,11 | 1.6 (1.7) | 0,6 | 4.7 (8.4) | 0,33 | 3.5 (4.9) | 0,32 |
| 15 - 19 | 6.5 (7.3) | 0,45 | 1.2 (1.5) | 0,8 | 1.3 (1.6) | 0,6 | 1.5 (2.2) | 0,14 | 1.9 (3.9) | 0,83 |
| 20 - 24 | 5.1 (4.1) | 0,26 | 0.6 (0.9) | 0,5 | 0.8 (1.1) | 0,3 | 0.7 (1.0) | 0,3 | 2.3 (6.1) | 0,45 |
| 25 - 34 | 5.6 (3.8) | 0,29 | 0.8 (1.0) | 0,6 | 0.8 (1.0) | 0,5 | 1.6 (3.7) | 0,21 | 2.3 (8.1) | 0,120 |
| 35 - 44 | 5.9 (4.9) | 0,36 | 0.7 (1.1) | 0,8 | 0.9 (1.1) | 0,6 | 2.6 (10.4) | 0,129 | 3.2 (40.3) | 0,674 |
| 45 - 54 | 5.9 (5.1) | 0,35 | 0.7 (1.0) | 0,4 | 1.6 (7.8) | 0,101 | 1.1 (2.1) | 0,22 | 1.2 (3.3) | 0,52 |
| 55 - 64 | 4.2 (3.4) | 0,16 | 0.6 (0.9) | 0,5 | 0.7 (1.2) | 0,10 | 0.7 (1.0) | 0,5 | 2.0 (7.8) | 0,51 |
| 65 - 69 | 4.6 (5.7) | 0,40 | 0.5 (0.8) | 0,5 | 1.0 (3.3) | 0,29 | 1.1 (2.6) | 0,25 | 0.9 (2.1) | 0,28 |
| 70 - 74 | 2.7 (3.1) | 0,13 | 0.4 (0.6) | 0,3 | 0.6 (0.8) | 0,3 | 0.6 (1.0) | 0,8 | 0.7 (1.0) | 0,17 |
| 75 - 79 | 2.7 (2.6) | 0,8 | 0.7 (0.8) | 0,3 | 0.8 (1.1) | 0,3 | 0.6 (0.6) | 0,3 | 0.7 (0.8) | 0,3 |
| 80 + | 3.0 (1.8) | 1,6 | 0.1 (0.3) | 0,1 | 0.0 (0.2) | 0,1 | 0.0 (0.1) | 0,2 | 0.0 (0.2) | 0,1 |
| **Sex of participants** | | | | | | | | | | |
| Female | 5.2 (4.7) | 0,45 | 0.9 (1.2) | 0,8 | 1.3 (4.9) | 0,101 | 1.2 (2.2) | 0,25 | 1.4 (2.8) | 0,52 |
| Male | 5.3 (4.8) | 0,44 | 0.8 (1.2) | 0,11 | 0.9 (1.8) | 0,29 | 1.9 (6.3) | 0,129 | 2.7 (20.2) | 0,674 |
| **Household size** | | | | | | | | | | |
| 1 | 3.7 (4.1) | 0,36 | 0.1 (0.4) | 0,5 | 0.2 (0.5) | 0,4 | 0.8 (8.1) | 0,129 | 2.2 (32.0) | 0,674 |
| 2 | 4.5 (4.1) | 0,45 | 0.5 (0.7) | 0,6 | 1.2 (5.9) | 0,101 | 1.1 (2.5) | 0,25 | 0.9 (2.0) | 0,38 |
| 3 | 5.4 (4.0) | 0,30 | 0.9 (1.0) | 0,8 | 1.0 (1.2) | 0,13 | 1.4 (2.2) | 0,14 | 1.9 (5.7) | 0,120 |
| 4 or more | 6.9 (5.5) | 0,44 | 1.7 (1.6) | 0,11 | 1.7 (2.0) | 0,13 | 3.1 (4.9) | 0,33 | 3.6 (7.7) | 0,83 |
| **Weekdays** | | | | | | | | | | |
| Monday | 4.9 (4.0) | 0,29 | 1.4 (1.5) | 0,6 | 1.2 (1.4) | 0,6 | 1.7 (3.0) | 0,20 | 2.8 (23.4) | 0,674 |
| Tuesday | 5.2 (4.7) | 0,40 | 1.0 (1.3) | 0,7 | 1.3 (1.7) | 0,13 | 1.1 (2.2) | 0,22 | 3.1 (10.1) | 0,120 |
| Wednesday | 5.2 (5.1) | 0,36 | 0.8 (1.5) | 0,6 | 0.9 (1.7) | 0,22 | 2.4 (6.1) | 0,33 | 1.6 (4.6) | 0,45 |
| Thursday | 5.5 (4.2) | 0,30 | 0.8 (1.1) | 0,8 | 1.0 (2.0) | 0,29 | 1.3 (2.4) | 0,25 | 1.0 (1.3) | 0,11 |
| Friday | 5.2 (4.1) | 0,23 | 0.9 (1.3) | 0,6 | 1.1 (1.8) | 0,12 | 2.6 (10.6) | 0,129 | 1.2 (2.5) | 0,23 |
| Saturday | 5.8 (5.2) | 0,44 | 1.1 (1.4) | 0,8 | 3.1 (15.0) | 0,101 | 1.2 (1.3) | 0,5 | 1.4 (2.1) | 0,13 |
| Sunday | 5.8 (6.3) | 0,45 | 0.9 (1.2) | 0,11 | 0.9 (1.2) | 0,5 | 1.1 (1.3) | 0,5 | 1.3 (2.6) | 0,83 |

1.2.c. Table 1.2c. Number of recorded overall contacts per different settings in the COVIMOD survey Waves 1 to 4 and the POLYMOD survey for the weighted analysis including group contacts.

**Note:** *The displayed educational contacts are based on the group of participants who attended an educational facility (kindergarten, school, university) and work contacts are based on the group of participants who reported to work full-/part-time.*

| | **POLYMOD** | | **COVIMOD** | | | | | | | |
|---|---|---|---|---|---|---|---|---|---|---|
| | | | **Wave1** | | **Wave2** | | **Wave3** | | **Wave4** | |
| | **Mean (SD)** | **Min, Max** | **Mean (SD)** | **Min, Max** | **Mean (SD)** | **Min, Max** | **Mean (SD)** | **Min, Max** | **Mean (SD)** | **Min, Max** |
| Overall | 19.6 (27.7) | 1,512 | 1.9 (1.8) | 0,16 | 3.5 (6.1) | 0,102 | 6.5 (21.6) | 0,500 | 7.3 (29.6) | 0,674 |
| Home | 2.9 (2.3) | 0,26 | 1.5 (1.5) | 0,12 | 1.6 (1.6) | 0,9 | 1.5 (1.5) | 0,13 | 1.5 (1.5) | 0,23 |
| Educational | 2.6 (4.3) | 0,42 | 0.0 (0.2) | 0,3 | 0.2 (1.0) | 0,10 | 0.1 (1.0) | 0,17 | 1.0 (3.9) | 0,60 |
| Work | 20.6 (32.8) | 0,509 | 0.3 (1.0) | 0,11 | 2.0 (7.3) | 0,100 | 6.1 (20.7) | 0,140 | 6.0 (32.1) | 0,491 |
| Transport | 0.3 (0.9) | 0,8 | 0.0 (0.2) | 0,3 | 0.1 (0.4) | 0,4 | 0.0 (0.3) | 0,8 | 0.1 (0.5) | 0,12 |
| Others | 2.9 (3.7) | 0,45 | 0.4 (0.9) | 0,10 | 1.0 (2.2) | 0,33 | 1.6 (5.7) | 0,149 | 2.5 (19.1) | 0,674 |

1.2.d. Table 1.2d. Number of recorded physical contacts per different settings in the COVIMOD survey Waves 1 to 4 and the POLYMOD survey for the weighted analysis including group contacts.

**Note:** *The displayed educational contacts are based on the group of participants who attended an educational facility (kindergarten, school, university) and work contacts are based on the group of participants who reported to work full-/part-time.*

| | **POLYMOD** | | **COVIMOD** | | | | | | | |
|---|---|---|---|---|---|---|---|---|---|---|
| | | | **Wave1** | | **Wave2** | | **Wave3** | | **Wave4** | |
| | **Mean (SD)** | **Min, Max** | **Mean (SD)** | **Min, Max** | **Mean (SD)** | **Min, Max** | **Mean (SD)** | **Min, Max** | **Mean (SD)** | **Min, Max** |
| Overall | 5.3 (4.7) | 0,45 | 0.8 (1.2) | 0,11 | 1.1 (3.7) | 0,101 | 1.6 (4.7) | 0,129 | 2.0 (14.5) | 0,674 |
| Home | 2.4 (2.2) | 0,26 | 0.8 (1.2) | 0,11 | 0.8 (1.2) | 0,8 | 0.9 (1.3) | 0,12 | 0.9 (1.2) | 0,8 |
| Educational | 1.4 (3.5) | 0,42 | 0.0 (0.0) | 0,1 | 0.1 (0.4) | 0,4 | 0.0 (0.3) | 0,3 | 0.3 (1.6) | 0,21 |
| Work | 1.6 (3.0) | 0,30 | 0.0 (0.1) | 0,2 | 0.5 (5.2) | 0,100 | 0.3 (1.7) | 0,25 | 0.7 (3.7) | 0,60 |
| Transport | 0.2 (0.7) | 0,6 | 0.0 (0.2) | 0,3 | 0.1 (0.3) | 0,3 | 0.0 (0.2) | 0,3 | 0.1 (0.4) | 0,7 |
| Others | 1.9 (3.0) | 0,40 | 0.1 (0.4) | 0,5 | 0.2 (0.6) | 0,11 | 0.5 (3.7) | 0,129 | 0.7 (13.8) | 0,674 |

### 1.3. Reproduction number estimates of SARS-CoV-2 under contact reduction measures.

1.3.a. Table 1.3a. Effective reproduction number at the timing of COVIMOD survey Waves 1 to 4, estimated effective reproduction number based on the reduction of social contacts at the times of COVIMOD survey Waves 1 to 4 assuming values of the basic reproduction number of Norm (2.6, SD=0.54) and the reduction in mobility at the times of the COVIMOD survey Waves, with 10000 bootstrapped samples in various settings and in different time frames for the weighted analysis including group contacts.

**Note:** *RKI - the percentage mean and minimum and maximum reductions in the reproduction number in the assessed time intervals, COVIMOD - the calculated percentage mean reductions in the reproduction number and the 95% confidence interval, Google and Apple mobility – the percentage mean and minimum and maximum reduction in mobility in the assessed time intervals.*

| | COVIMOD | | RKI | | Google | | Apple | |
|---|---|---|---|---|---|---|---|---|
| | Mean reproduction number (Min, Max) | Percent Mean Reduction Number (Min, Max) | Mean reproduction number (Min, Max) | Percent Mean Reduction Number (Min, Max) | Mean reproduction number (Min, Max) | Percent Mean Reduction Number (Min, Max) | Mean reproduction number (Min, Max) | Percent Mean Reduction Number (Min, Max) |
| **Overall** | | | | | | | | |
| 30th April - 6th May 2020 | 0.24 (0.14, 0.36) | -90.00 (-94.00, -86.00) | 0.88 (0.79, 0.97) | -66.00 (-69.00, -62.00) | - | -24.31 (-59.40, -15.20) | - | -36.77 (-44.17, -29.61) |
| 14th May - 21st May 2020 | 0.35 (0.20, 0.50) | -86.00 (-92.00, -80.00) | 0.90 (0.72, 1.15) | -65.00 (-72.00, -55.00) | - | -16.85 (-50.20, -3.40) | - | -15.64 (-25.26, -2.58) |
| 28th May - 4th June 2020 | 0.51 (0.28, 0.79) | -80.00 (-89.00, -69.00) | 1.06 (0.84, 1.39) | -59.00 (-67.00, -46.00) | - | -13.05 (-47.40, 1.60) | - | 5.18 (-4.00, 19.43) |
| 11th June - 22nd June 2020 | 0.68 (0.34, 1.18) | -73.00 (-86.00, -54.00) | 0.84 (0.60, 1.07) | -67.00 (-76.00, -58.00) | - | -10.63 (-34.60, 5.60) | - | 11.66 (1.62, 28.00) |
| **Home** | | | | | | | | |
| 30th April - 6th May 2020 | 1.40 (0.82, 2.02) | -46.00 (-68.00, -22.00) | - | - | - | 12.71 (6.00, 28.00) | - | - |
| 14th May - 21st May 2020 | 1.35 (0.78, 1.96) | -48.00 (-70.00, -24.00) | - | - | - | 8.62 (3.00, 18.00) | - | - |
| 28th May - 4th June 2020 | 1.42 (0.81, 2.09) | -45.00 (-68.00, -19.00) | - | - | - | 6.88 (0.00, 16.00) | - | - |
| 11th June - 22nd June 2020 | 1.25 (0.73, 1.81) | -51.00 (-71.00, -30.00) | - | - | - | 5.25 (-1.00, 16.00) | - | - |
| **Educational** | | | | | | | | |
| 30th April - 6th May 2020 | 0.04 (0.01, 0.11) | -98.00 (-99.00, -95.00) | - | - | - | - | - | - |
| 14th May - 21st May 2020 | 0.20 (0.04, 0.64) | -92.00 (-98.00, -75.00) | - | - | - | - | - | - |
| 28th May - 4th June 2020 | 0.13 (0.03, 0.34) | -95.00 (-98.00, -86.00) | - | - | - | - | - | - |
| 11th June - 22nd June 2020 | 0.71 (0.13, 2.16) | -72.00 (-95.00, -16.00) | - | - | - | - | - | - |
| **Work** | | | | | | | | |
| 30th April - 6th May 2020 | 0.04 (0.02, 0.08) | -98.00 (-99.00, -96.00) | - | - | - | -34.71 (-84.00, -18.00) | - | - |
| 14th May - 21st May 2020 | 0.13 (0.07, 0.22) | -95.00 (-97.00, -91.00) | - | - | - | -27.00 (-81.00, -1.00) | - | - |
| 28th May - 4th June 2020 | 0.29 (0.13, 0.53) | -88.00 (-95.00, -79.00) | - | - | - | -24.12 (-80.00, 8.00) | - | - |
| 11th June - 22nd June 2020 | 0.34 (0.17, 0.56) | -86.00 (-93.00, -78.00) | - | - | - | -16.50 (-59.00, 15.00) | - | - |
| **Transport** | | | | | | | | |
| 30th April - 6th May 2020 | 0.63 (0.11, 2.20) | -75.00 (-95.00, -15.00) | - | - | - | -42.14 (-67.00, -35.00) | - | - |
| 14th May - 21st May 2020 | 0.90 (0.14, 3.18) | -65.00 (-94.00, 23.00) | - | - | - | -31.75 (-46.00, -25.00) | - | - |
| 28th May - 4th June 2020 | 0.53 (0.10, 1.78) | -79.00 (-96.00, -31.00) | - | - | - | -25.12 (-42.00, -13.00) | - | - |
| 11th June - 22nd June 2020 | 1.24 (0.17, 4.99) | -52.00 (-93.00, 92.00) | - | - | - | -23.00 (-40.00, -11.00) | - | - |
| **Others** | | | | | | | | |
| 30th April - 6th May 2020 | 0.29 (0.13, 0.52) | -88.00 (-95.00, -80.00) | - | - | - | -28.71 (-87.00, -7.50) | - | - |
| 14th May - 21st May 2020 | 0.56 (0.26, 1.03) | -78.00 (-89.00, -60.00) | - | - | - | -17.06 (-71.00, 14.00) | - | - |
| 28th May - 4th June 2020 | 1.04 (0.46, 1.97) | -60.00 (-82.00, -24.00) | - | - | - | -11.44 (-65.50, 6.50) | - | - |
| 11th June - 22nd June 2020 | 1.47 (0.50, 3.54) | -43.00 (-80.00, 37.00) | - | - | - | -9.46 (-45.00, 12.50) | - | - |

1.4. **Figure 1.4. Boxplots of the number of overall contacts during the POLYMOD and COVIMOD survey Waves 1 to 4 in various settings. The boundaries of the boxes closest to zero indicates the 25th percentiles, the lines within the boxes marks the medians, the red dots within the boxes marks the means, and the boundary of the boxes farthest from zero indicates the 75th percentiles. Whiskers above and below the boxes indicate the 10th and 90th percentiles; black dots represent outliers. Participants with no contacts are displayed as 0 on the log-scale of the y-axis.**

1.4.a. Figure 1.4a. Displayed are (A) Female and (B) Male for the weighted analysis including group contacts in overall contacts.

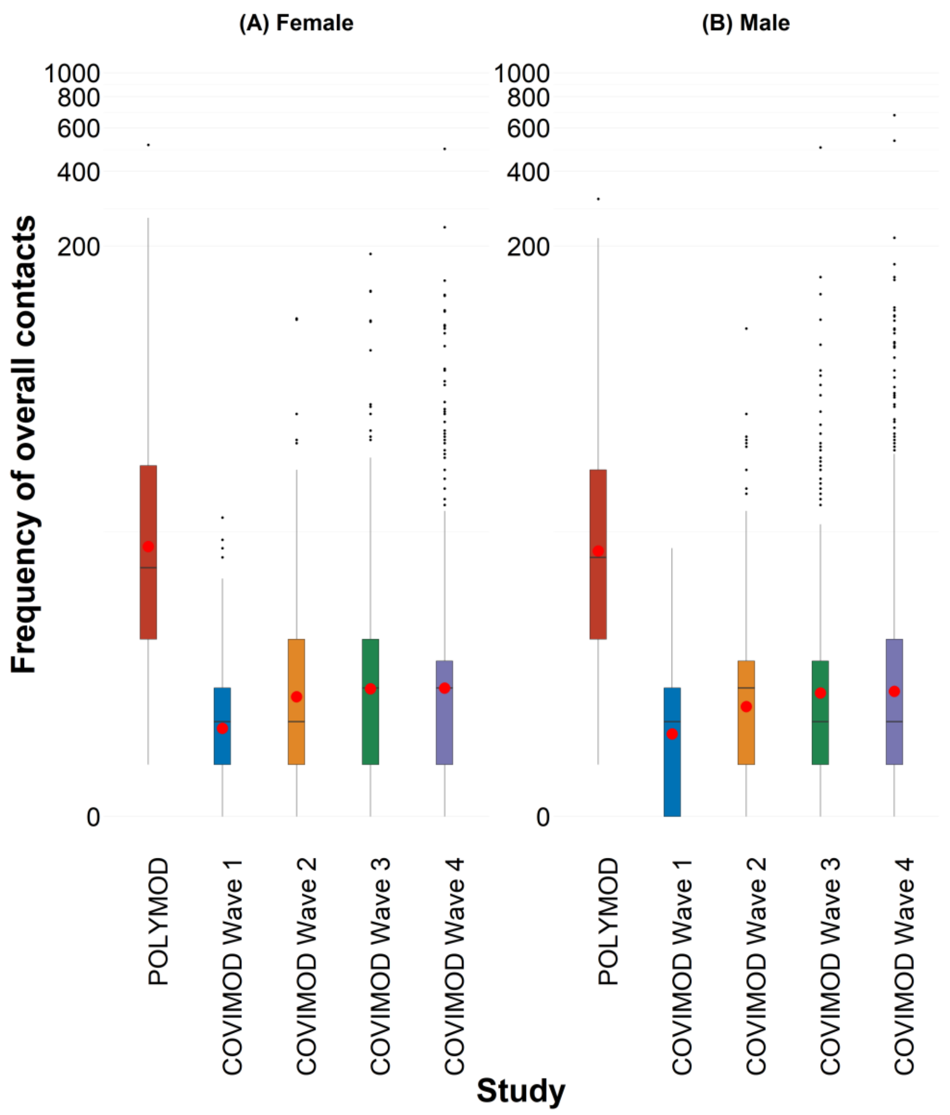

1.4.b. Figure 1.4b. Displayed are (A) Monday, (B) Tuesday, (C) Wednesday, (D) Thursday, (E) Friday, (F) Saturday and (G) Sunday for the weighted analysis including group contacts in overall contacts.

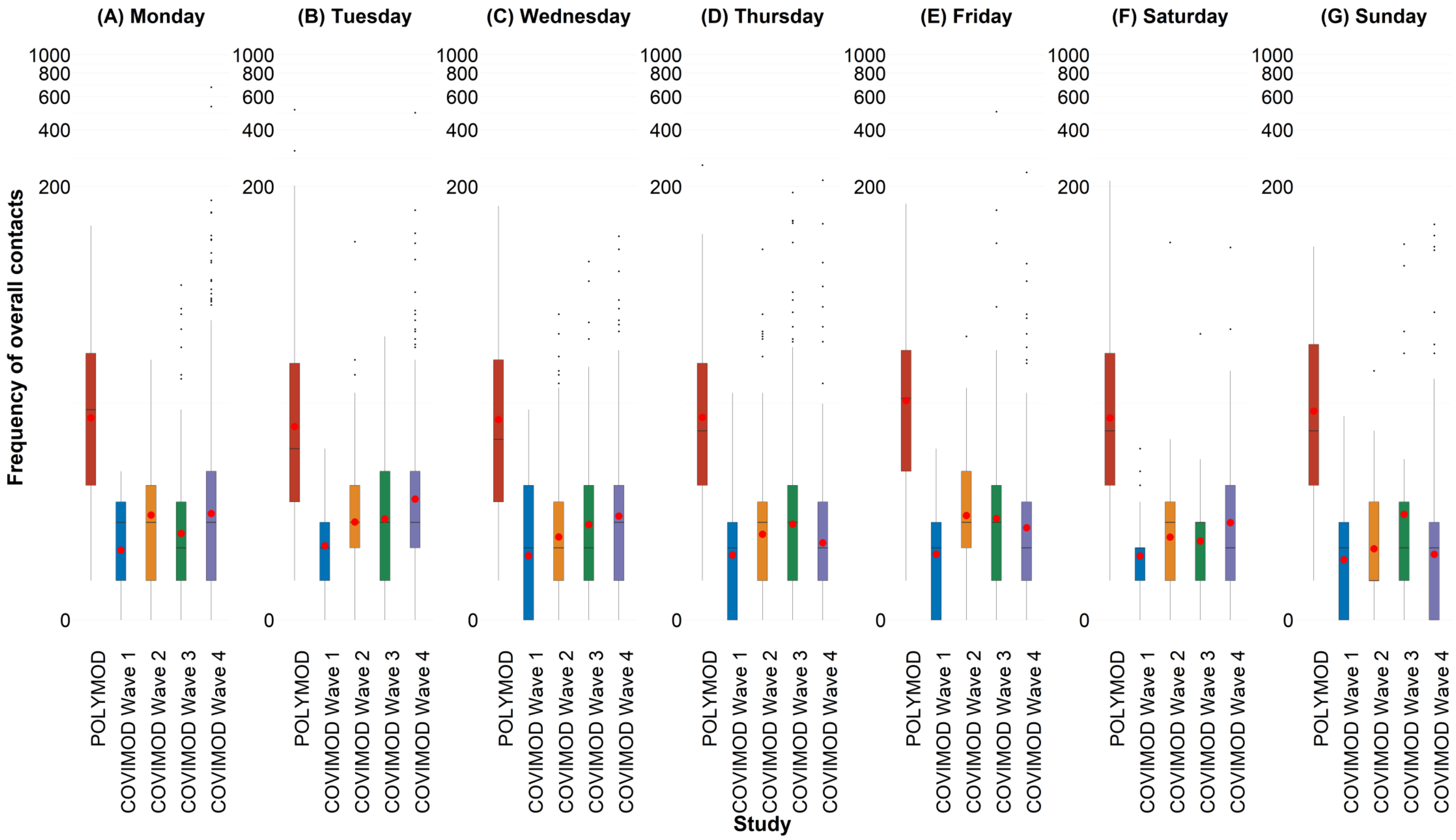

1.5. **Figure 1.5. Boxplots of the number of physical contacts during the POLYMOD and COVIMOD survey Waves 1 to 4 in various settings. The boundaries of the boxes closest to zero indicates the 25th percentiles, the lines within the boxes marks the medians, the red dots within the boxes marks the means, and the boundary of the boxes farthest from zero indicates the 75th percentiles. Whiskers above and below the boxes indicate the 10th and 90th percentiles; black dots represent outliers. Participants with no contacts are displayed as 0 on the log-scale of the y-axis**.

1.5.a. Figure 1.5a. Displayed are (A) the overall number of contacts, (B) home contacts, (C) work contacts, (D) educational contacts, (E) public transport contacts and (F) other contacts for the weighted analysis including group contacts in physical contacts.

**Note:** *The displayed educational contacts are based on the group of participants who attended an educational facility (kindergarten, school, university) and work contacts are based on the group of participants who reported to work full-/part-time.*

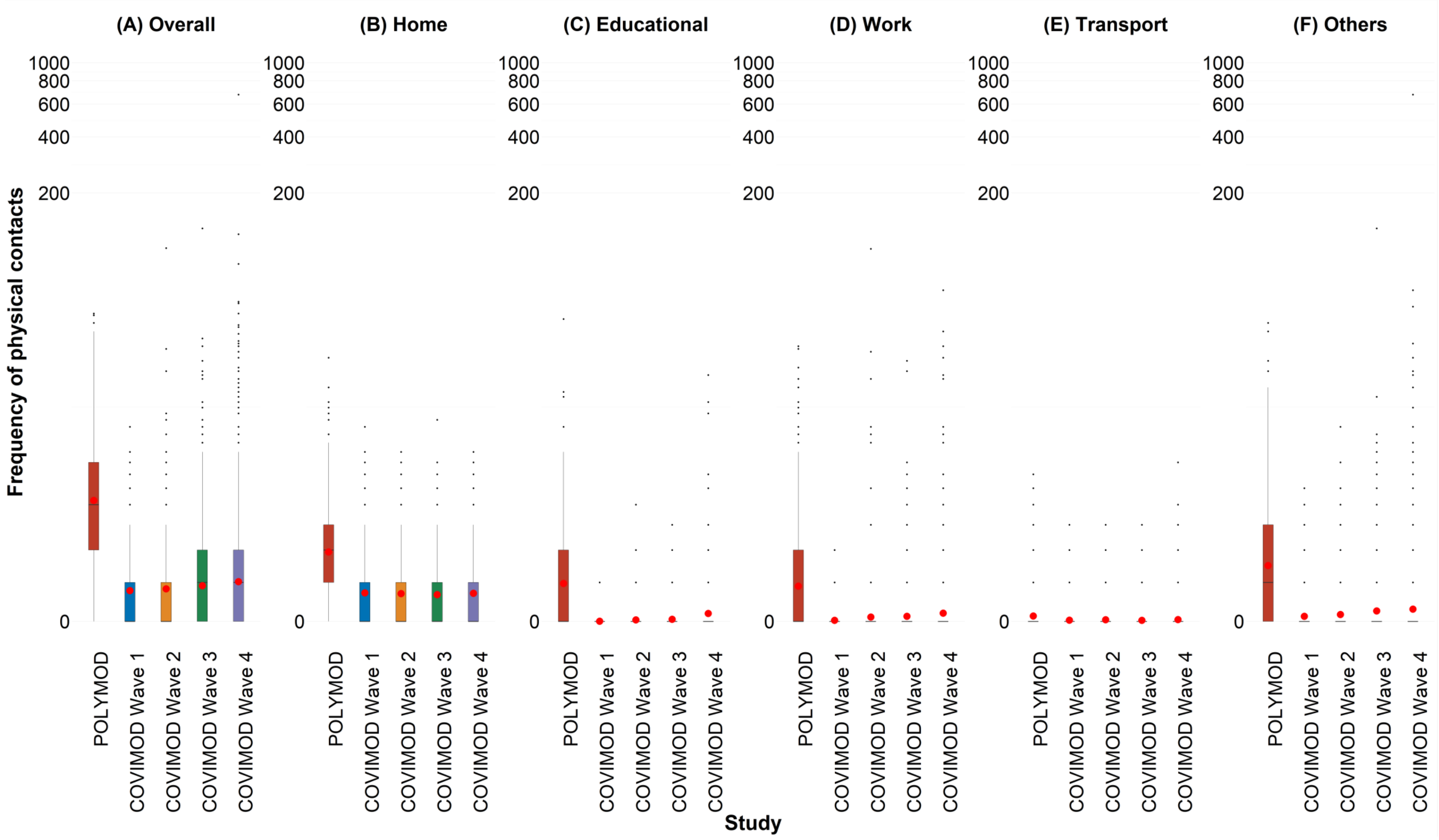

1.5.b. Figure 1.5b. Displayed are (A) 0 - 7 years, (B) 8 – 15 years, (C) 16 – 23 years, (D) 24 – 38 years, (E) 39 – 60 years and (F) 61 years or more for the weighted analysis including group contacts in physical contacts.

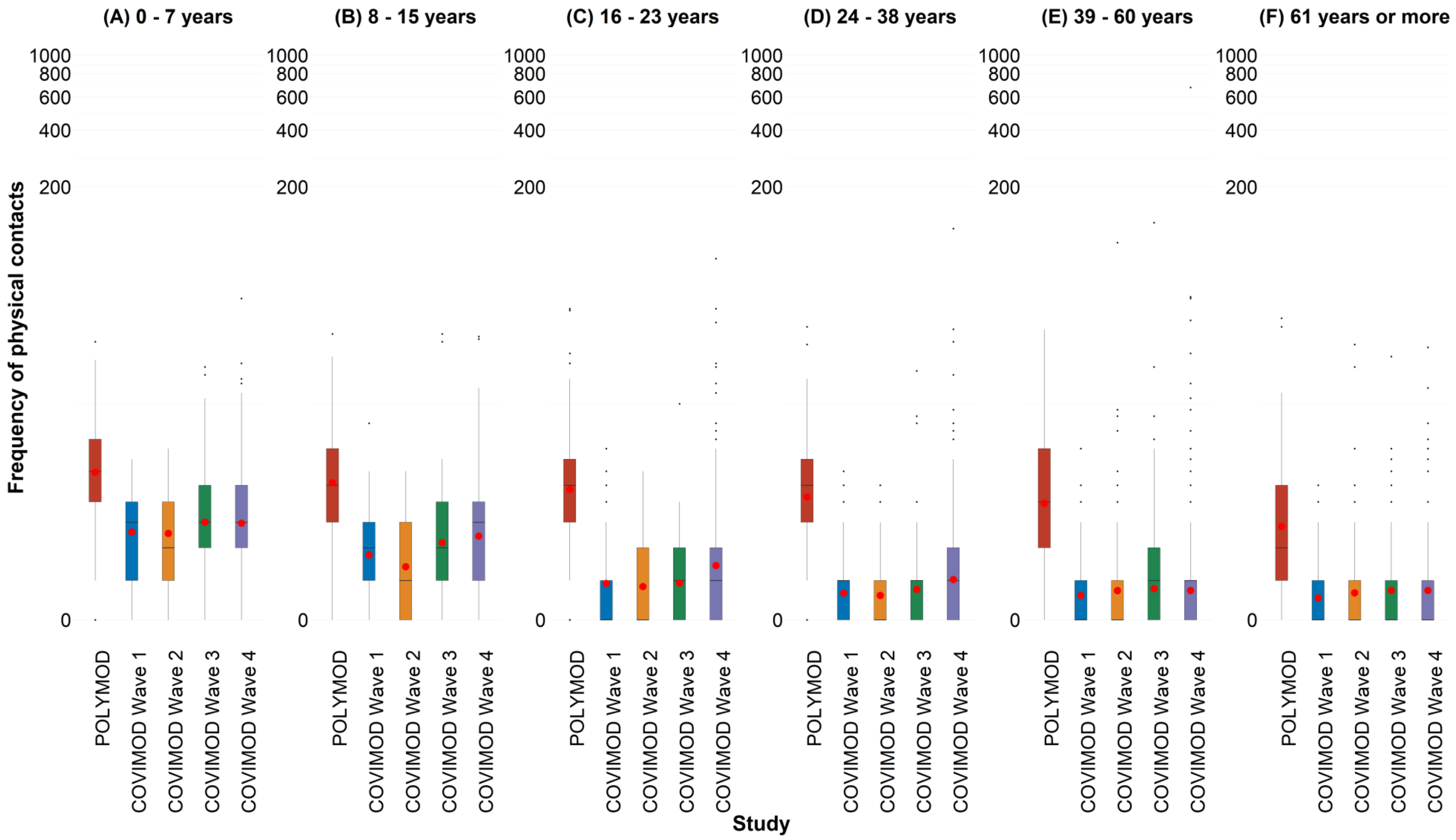

1.5.c. Figure 1.5c. Displayed are (A) Household size of 1, (B) Household size of 2, (C) Household size of 3 and (D) Household size of 4 or more for the weighted analysis including group contacts in physical contacts.

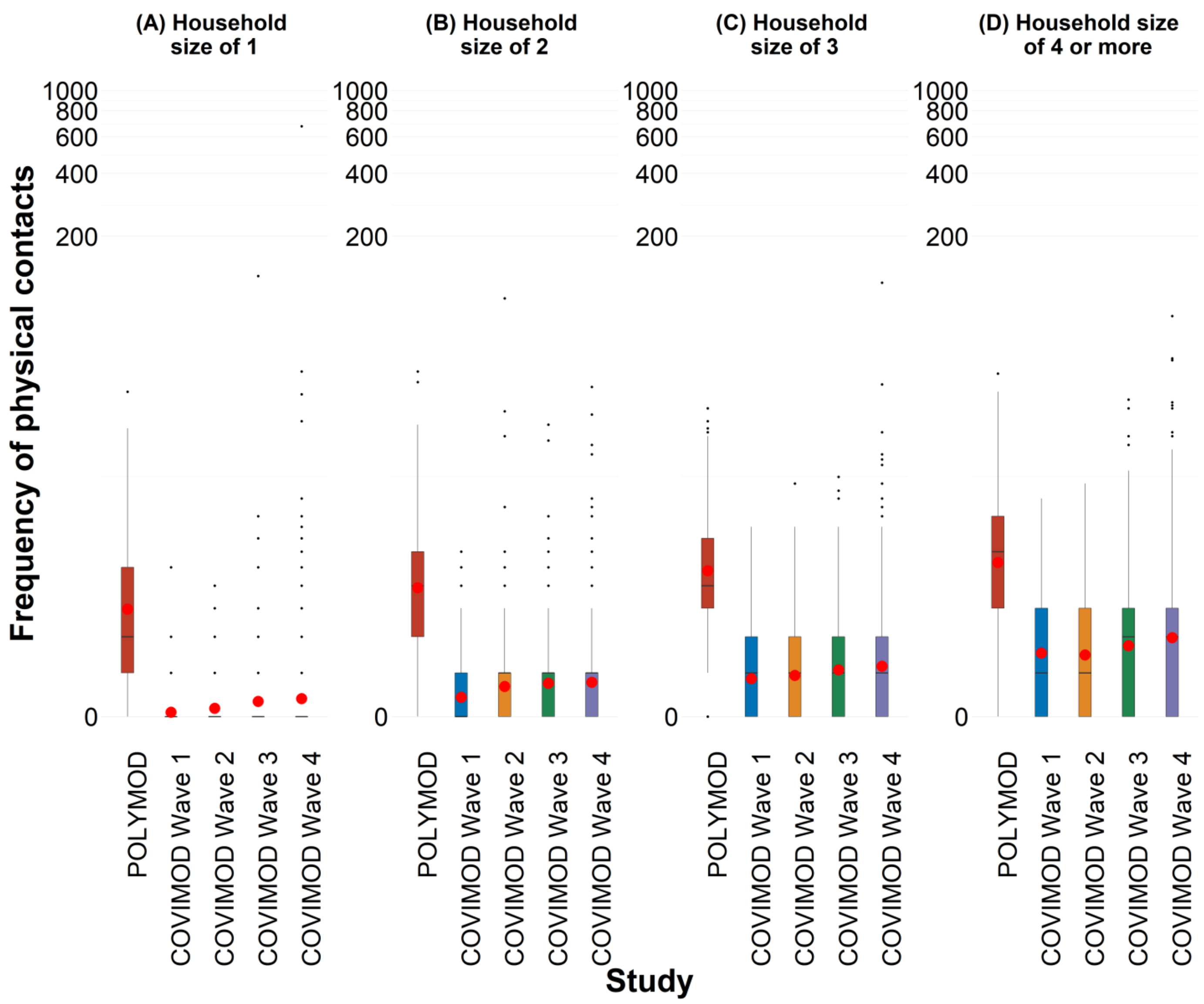

1.5.d. Figure 1.5d. Displayed are (A) Female and (B) Male for the weighted analysis including group contacts in physical contacts.

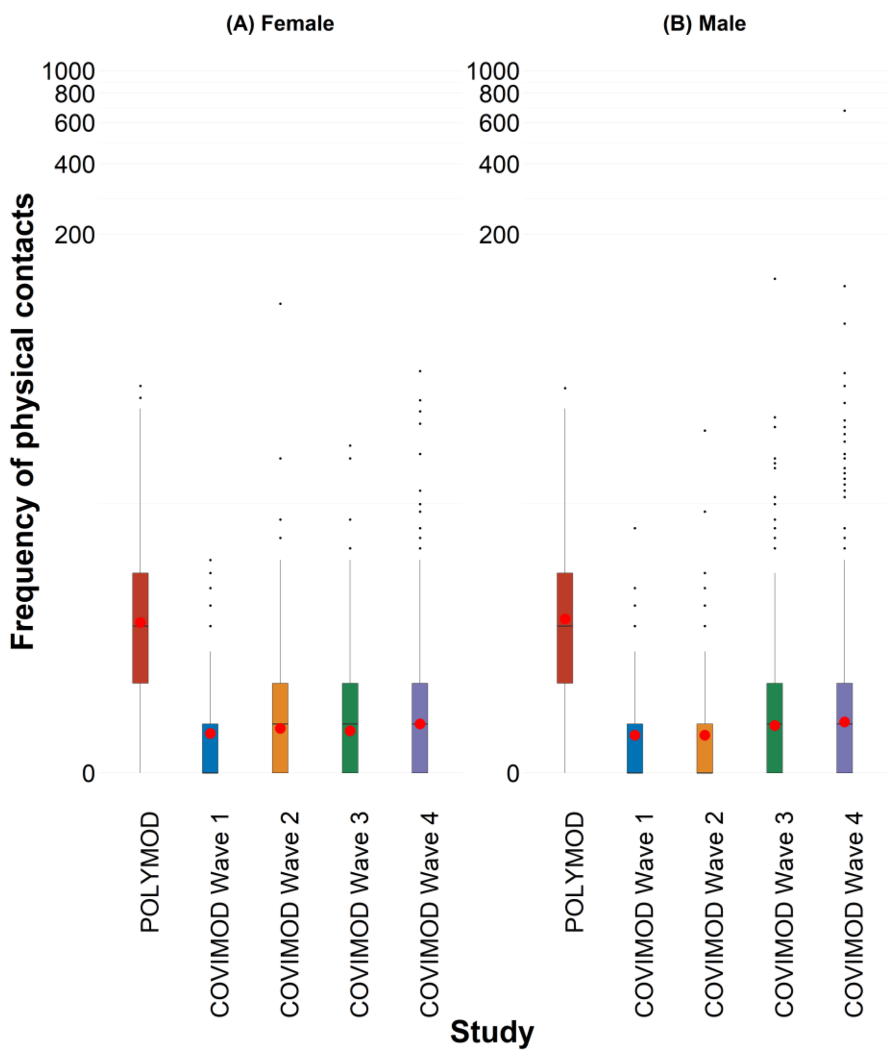

1.5.e. Figure 1.5e. Displayed are (A) Monday, (B) Tuesday, (C) Wednesday, (D) Thursday, (E) Friday, (F) Saturday and (G) Sunday for the weighted analysis including group contacts in physical contacts.

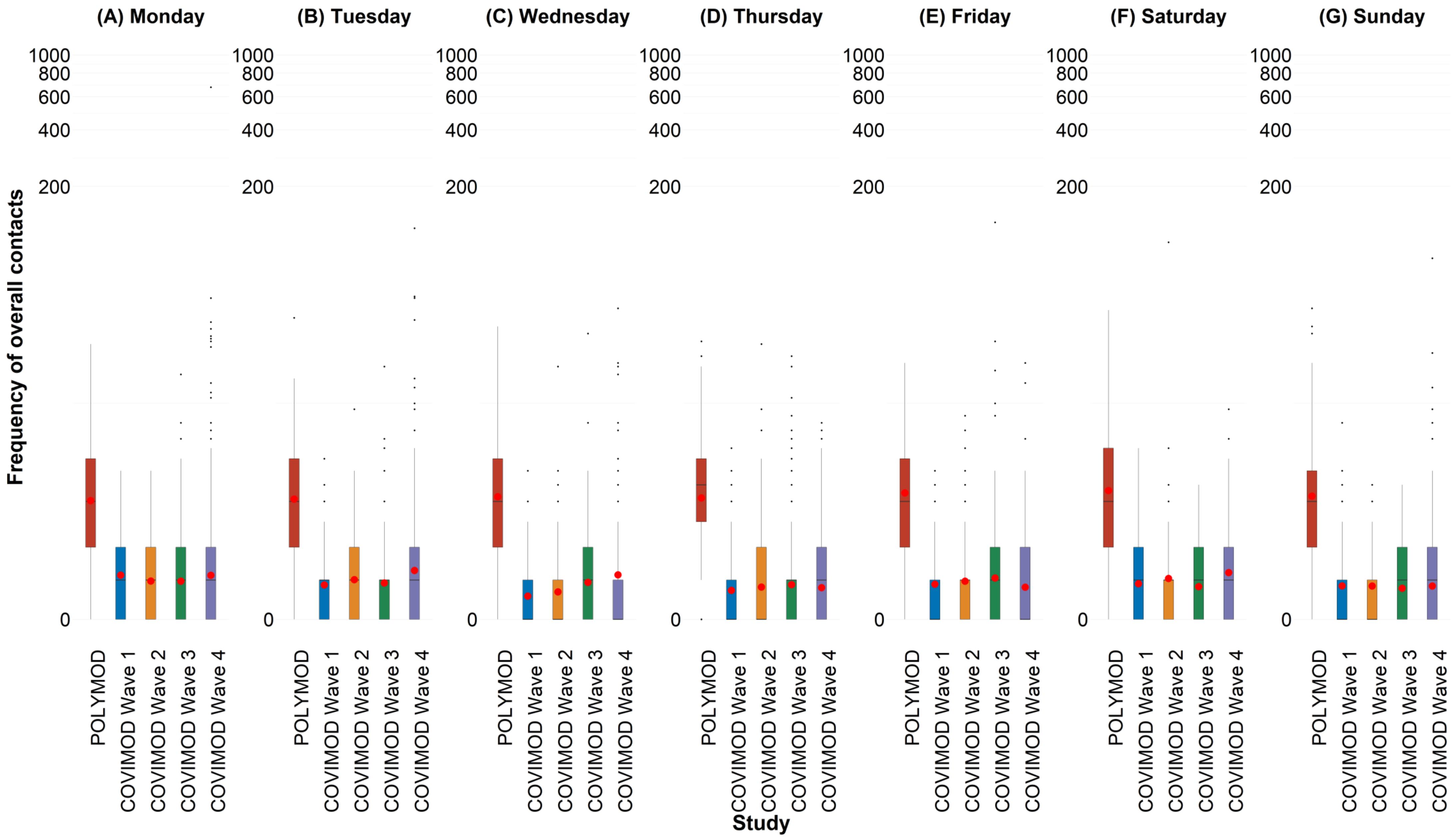

### 1.6. **Social contact patterns**.

1.6.a. Figure 1.6a. Social contact matrices with the mean physical number of reported daily social contacts by participants in different age groups with individuals in other age groups in POLYMOD and COVIMOD survey waves 1 to 4 in various settings. Displayed are (A) the overall number of contacts, (B) home contacts, (C) educational contacts, (D) work contacts, (E) public transport contacts and (F) other contacts for the weighted analysis including group contacts.

**Note:** *Participants with more than more than 100 group contacts were removed in COVIMOD and POLYMOD. Specifically, 6 participants and 13 participants were removed in Wave 3 and Wave 4 respectively, while 10 participants were removed in POLYMOD.*

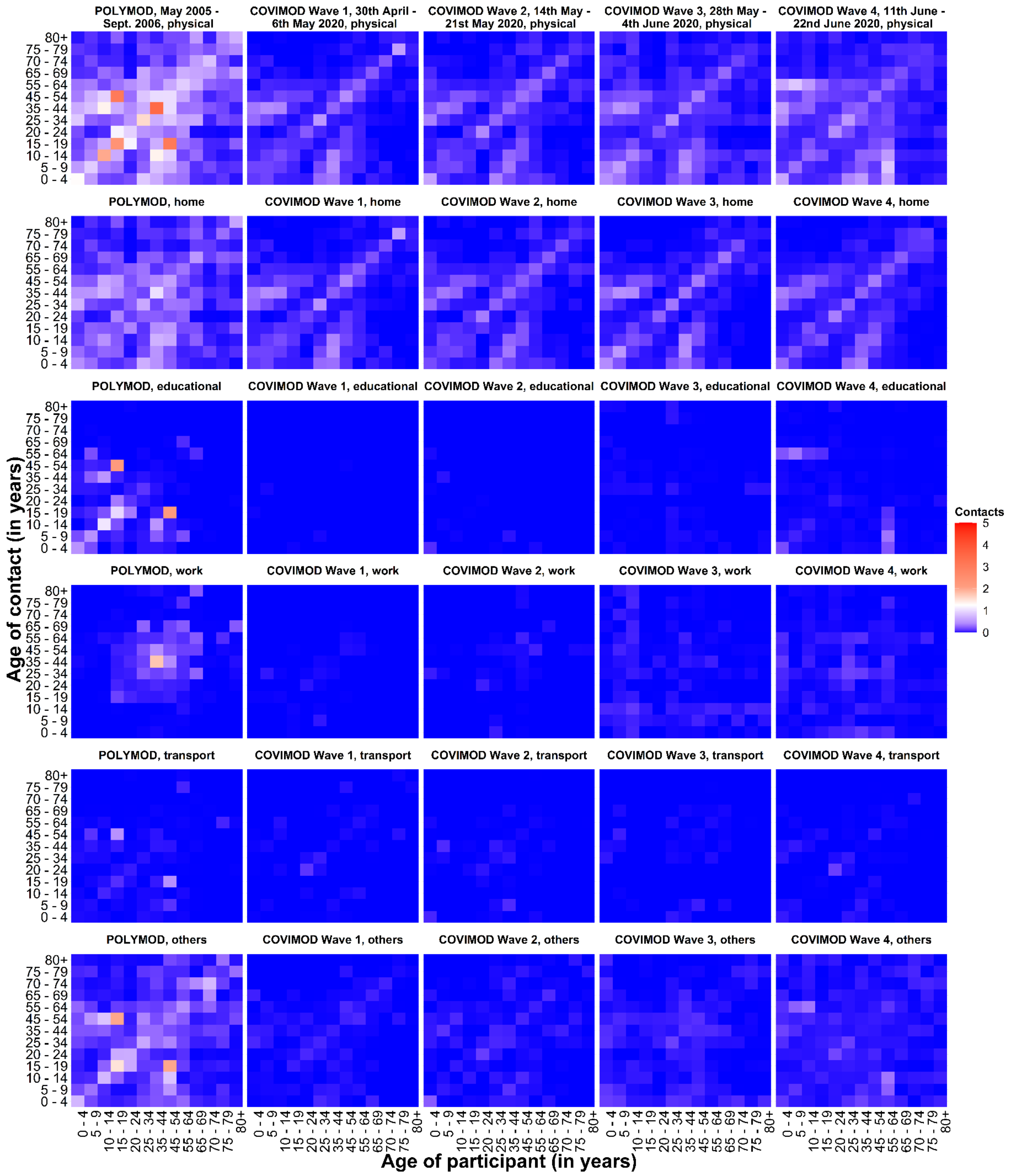

1.7. **Estimated reproduction number of SARS-CoV-2 under contact reduction measures. RKI - the percentage mean and minimum and maximum reductions in the reproduction number in the assessed time intervals, COVIMOD - the calculated percentage mean reductions in the reproduction number and the 95% confidence interval, Google and Apple mobility – the percentage mean and minimum and maximum reduction in mobility in the assessed time intervals.**

1.7.a. Figure 1.7a. Comparison of R(t) estimates obtained based on different input data. Measured effective reproduction number at the timing of COVIMOD survey Waves 1 to 4 (red), estimated reproduction number based on the reduction of social contacts at the times of COVIMOD survey Waves 1 to 4 (blue) and the reduction in mobility at the times of the COVIMOD survey Waves (yellow and green) for the weighted analysis including group contacts. Displayed are (A) 30th April – 6th May 2020, (B) 14th May – 21st May 2020, (C) 28th May – 4th June 2020 and (D) 11th June – 22nd June 2020

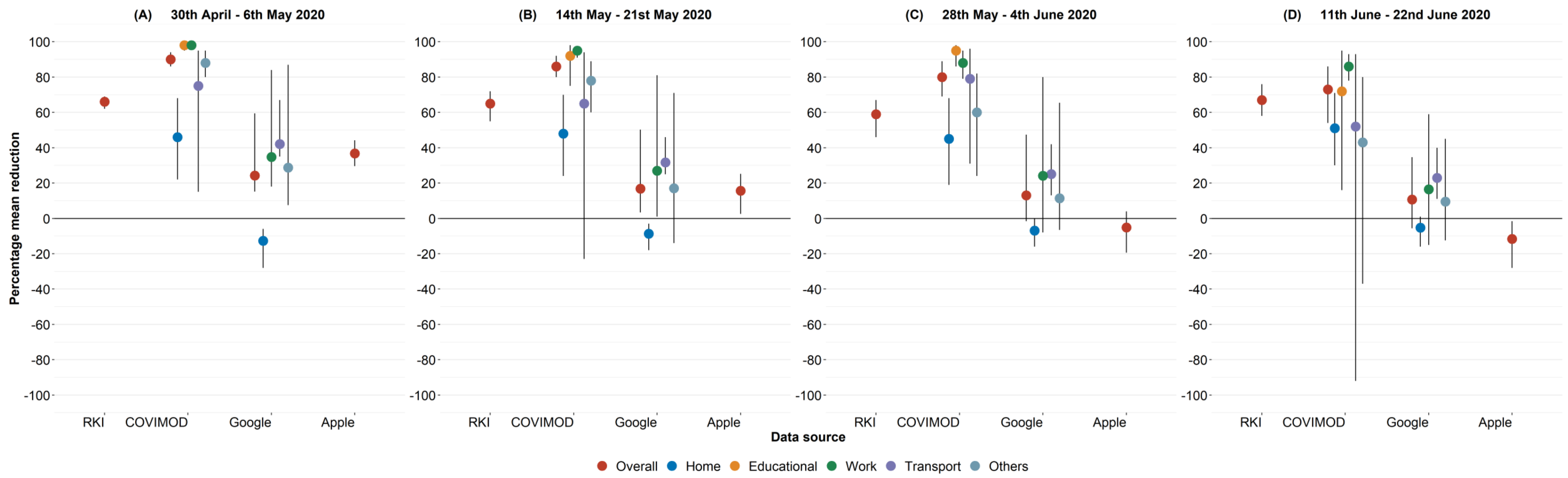

### 2. UNWEIGHTED ANALYSIS INCLUDING GROUP CONTACTS

**COVIMOD Wave 3 - 4 includes group contacts in home, work, and educational settings while POLYMOD includes group contacts in work setting.**

#### 2.1. Participant characteristics.

2.1.a. Table 2.1a. Participant characteristics in the COVIMOD survey Waves 1 to 4 and in the POLYMOD survey for the unweighted analysis including group contacts.

| | POLYMOD | | COVIMOD | | | | | | | |
|---|---|---|---|---|---|---|---|---|---|---|
| | | | Wave 1 | | Wave 2 | | Wave 3 | | Wave 4 | |
| | N | percent | N | percent | N | percent | N | percent | N | percent |
| | **1341** | **-** | **1560** | **-** | **1356** | **-** | **1081** | **-** | **1890** | **-** |
| **Age category** | | | | | | | | | | |
| 0 - 4 | 89 | 6.9 | 46 | 2.9 | 36 | 2.7 | 21 | 1.9 | 56 | 3.0 |
| 5 - 9 | 92 | 7.1 | 48 | 3.1 | 41 | 3.0 | 30 | 2.8 | 62 | 3.3 |
| 10 - 14 | 110 | 8.5 | 73 | 4.7 | 63 | 4.7 | 45 | 4.2 | 87 | 4.6 |
| 15 - 19 | 121 | 9.3 | 95 | 6.1 | 66 | 4.9 | 45 | 4.2 | 108 | 5.7 |
| 20 - 24 | 117 | 9.0 | 83 | 5.3 | 60 | 4.4 | 28 | 2.6 | 109 | 5.8 |
| 25 - 34 | 132 | 10.2 | 173 | 11.1 | 148 | 10.9 | 96 | 8.9 | 219 | 11.6 |
| 35 - 44 | 156 | 12.0 | 137 | 8.8 | 124 | 9.2 | 91 | 8.4 | 164 | 8.7 |
| 45 - 54 | 184 | 14.2 | 235 | 15.1 | 209 | 15.4 | 174 | 16.1 | 275 | 14.6 |
| 55 - 64 | 160 | 12.4 | 265 | 17.0 | 244 | 18.0 | 237 | 21.9 | 321 | 17.0 |
| 65 - 69 | 74 | 5.7 | 270 | 17.3 | 245 | 18.1 | 199 | 18.4 | 313 | 16.6 |
| 70 - 74 | 33 | 2.5 | 89 | 5.7 | 73 | 5.4 | 79 | 7.3 | 118 | 6.2 |
| 75 - 79 | 14 | 1.1 | 35 | 2.2 | 34 | 2.5 | 29 | 2.7 | 48 | 2.5 |
| 80 + | 13 | 1.0 | 11 | 0.7 | 11 | 0.8 | 7 | 0.6 | 10 | 0.5 |
| Missing | 46 | - | 0 | - | 2 | - | 0 | - | 0 | - |
| **Sex of participants** | | | | | | | | | | |
| Female | 722 | 55.4 | 748 | 48.1 | 638 | 47.1 | 536 | 49.6 | 901 | 47.8 |
| Male | 581 | 44.6 | 806 | 51.9 | 717 | 52.9 | 544 | 50.4 | 985 | 52.2 |
| Missing | 38 | - | 6 | - | 1 | - | 1 | - | 4 | - |
| **Household size** | | | | | | | | | | |
| 1 | 250 | 18.6 | 232 | 14.9 | 256 | 18.9 | 268 | 24.8 | 487 | 25.8 |
| 2 | 411 | 30.6 | 412 | 26.4 | 351 | 25.9 | 270 | 25.0 | 439 | 23.2 |
| 3 | 339 | 25.3 | 514 | 32.9 | 447 | 33.0 | 343 | 31.7 | 544 | 28.8 |
| 4 or more | 341 | 25.4 | 402 | 25.8 | 302 | 22.3 | 200 | 18.5 | 420 | 22.2 |
| Missing | 0 | - | 0 | - | 0 | - | 0 | - | 0 | - |
| **Weekdays** | | | | | | | | | | |
| Monday | 227 | 17.2 | 50 | 3.2 | 60 | 4.4 | 128 | 11.8 | 642 | 34.0 |
| Tuesday | 237 | 18.0 | 63 | 4.0 | 89 | 6.6 | 143 | 13.2 | 246 | 13.0 |
| Wednesday | 222 | 16.8 | 54 | 3.5 | 293 | 21.6 | 87 | 8.0 | 172 | 9.1 |
| Thursday | 179 | 13.6 | 914 | 58.6 | 613 | 45.2 | 489 | 45.2 | 320 | 16.9 |
| Friday | 186 | 14.1 | 144 | 9.2 | 117 | 8.6 | 132 | 12.2 | 196 | 10.4 |
| Saturday | 152 | 11.5 | 88 | 5.6 | 63 | 4.6 | 66 | 6.1 | 81 | 4.3 |
| Sunday | 117 | 8.9 | 247 | 15.8 | 121 | 8.9 | 36 | 3.3 | 233 | 12.3 |
| Missing | 21 | - | 0 | - | 0 | - | 0 | - | 0 | - |

### 2.2. Number of social contacts.

2.2.a. Table 2.2a. Number of recorded overall contacts per participant per day stratified by age, gender, household size, and day of the week in the COVIMOD survey Waves 1 to 4 and the POLYMOD survey for the unweighted analysis including group contacts.

| | POLYMOD | | | COVIMOD | | | | | | | | | | | |
|---|---|---|---|---|---|---|---|---|---|---|---|---|---|---|---|
| | | | | Wave1 | | | Wave 2 | | | Wave 3 | | | Wave 4 | | |
| | **N** | **Mean (SD)** | **Min, Max** | **N** | **Mean (SD)** | **Min, Max** | **N** | **Mean (SD)** | **Min, Max** | **N** | **Mean (SD)** | **Min, Max** | **N** | **Mean (SD)** | **Min, Max** |
| | **1341** | **20.2 (28.5)** | **1,512** | **1560** | **2.1 (1.9)** | **0,16** | **1356** | **3.6 (6.1)** | **0,102** | **1081** | **5.9 (20.3)** | **0,500** | **1890** | **7.1 (27.9)** | **0,674** |
| **Age category** | | | | | | | | | | | | | | | |
| 0 - 4 | 89 | 9.1 (6.1) | 2,30 | 46 | 3.8 (1.9) | 0,8 | 36 | 4.7 (2.7) | 0,14 | 21 | 7.3 (6.5) | 2, 24 | 56 | 12.9 (29.2) | 0,198 |
| 5 - 9 | 92 | 8.4 (5.7) | 2,43 | 48 | 3.5 (1.7) | 0,8 | 41 | 4.4 (3.2) | 0,17 | 30 | 6.9 (6.7) | 0, 27 | 62 | 6.4 (5.5) | 0, 31 |
| 10 - 14 | 110 | 13.3 (12.6) | 2,61 | 73 | 3.2 (1.8) | 0,12 | 63 | 3.3 (2.0) | 0,9 | 45 | 19.0 (76.1) | 0,500 | 87 | 9.9 (17.7) | 0,110 |
| 15 - 19 | 121 | 32.8 (28.6) | 1,215 | 95 | 3.3 (2.0) | 0,8 | 66 | 4.1 (3.7) | 0,20 | 45 | 5.1 (6.0) | 0, 27 | 108 | 11.1 (20.2) | 0,113 |
| 20 - 24 | 117 | 29.1 (32.9) | 1,310 | 83 | 2.0 (2.6) | 0,16 | 60 | 3.0 (3.3) | 0,20 | 28 | 5.7 (9.7) | 0, 45 | 109 | 9.9 (23.6) | 0,169 |
| 25 - 34 | 132 | 31.9 (50.9) | 2,512 | 173 | 1.7 (1.8) | 0,13 | 148 | 2.8 (3.2) | 0,18 | 96 | 5.2 (12.7) | 0,100 | 219 | 7.1 (19.0) | 0,150 |
| 35 - 44 | 156 | 25.0 (21.7) | 1,111 | 137 | 1.9 (2.2) | 0,11 | 124 | 3.0 (3.7) | 0,33 | 91 | 7.8 (22.3) | 0,150 | 164 | 8.1 (53.2) | 0,674 |
| 45 - 54 | 184 | 26.2 (35.1) | 1,260 | 235 | 1.8 (1.7) | 0,9 | 209 | 4.0 (10.3) | 0,102 | 174 | 4.6 (11.3) | 0,131 | 275 | 9.4 (47.0) | 0,532 |
| 55 - 64 | 160 | 13.9 (19.5) | 1,150 | 265 | 1.9 (1.7) | 0,12 | 244 | 3.7 (5.1) | 0,42 | 237 | 5.6 (16.1) | 0,186 | 321 | 4.7 (11.6) | 0,127 |
| 65 - 69 | 74 | 8.6 (10.6) | 1,58 | 270 | 1.8 (1.7) | 0,11 | 245 | 3.9 (7.6) | 0,93 | 199 | 5.2 (12.5) | 0,101 | 313 | 5.4 (16.9) | 0,216 |
| 70 - 74 | 33 | 5.8 (5.7) | 1,25 | 89 | 1.8 (1.5) | 0,8 | 73 | 3.1 (3.5) | 0,25 | 79 | 3.2 (2.5) | 0, 13 | 118 | 3.1 (4.5) | 0, 37 |
| 75 - 79 | 14 | 4.6 (3.7) | 1,14 | 35 | 1.7 (1.4) | 0,5 | 34 | 2.6 (2.1) | 0,9 | 29 | 3.9 (5.9) | 0, 32 | 48 | 3.2 (3.0) | 0, 17 |
| 80 + | 13 | 3.7 (1.8) | 2,8 | 11 | 1.1 (1.0) | 0,3 | 11 | 2.0 (2.8) | 0,9 | 7 | 2.1 (2.4) | 0, 7 | 10 | 1.7 (1.9) | 0, 4 |
| **Sex of participants** | | | | | | | | | | | | | | | |
| Female | 722 | 21.4 (31.4) | 1,512 | 748 | 2.2 (2.0) | 0,16 | 638 | 3.9 (6.8) | 0,102 | 536 | 5.7 (15.1) | 0,186 | 901 | 6.5 (22.3) | 0,493 |
| Male | 581 | 19.1 (24.9) | 1,310 | 806 | 2.0 (1.8) | 0,12 | 717 | 3.3 (5.3) | 0,93 | 544 | 6.0 (24.4) | 0,500 | 985 | 7.5 (31.7) | 0,674 |
| **Household size** | | | | | | | | | | | | | | | |
| 1 | 250 | 18.3 (23.7) | 1,158 | 232 | 0.8 (1.5) | 0,12 | 256 | 2.1 (4.2) | 0,42 | 268 | 3.6 (11.2) | 0,150 | 487 | 5.9 (34.6) | 0,674 |
| 2 | 411 | 19.9 (34.1) | 1,512 | 412 | 1.3 (1.6) | 0,11 | 351 | 3.8 (8.4) | 0,101 | 270 | 6.3 (17.6) | 0,132 | 439 | 6.8 (30.4) | 0,532 |
| 3 | 339 | 20.9 (27.9) | 1,310 | 514 | 2.2 (1.5) | 0,11 | 447 | 3.6 (6.0) | 0,102 | 343 | 5.3 (12.6) | 0,186 | 544 | 7.4 (27.2) | 0,493 |
| 4 or more | 341 | 21.3 (24.6) | 2,260 | 402 | 3.5 (1.9) | 0,16 | 302 | 4.5 (3.5) | 0,32 | 200 | 9.3 (36.9) | 0,500 | 420 | 8.6 (14.3) | 0,109 |
| **Weekdays** | | | | | | | | | | | | | | | |
| Monday | 227 | 17.4 (17.2) | 1,124 | 50 | 2.2 (1.7) | 0,6 | 60 | 4.3 (4.1) | 0,24 | 128 | 4.6 (9.2) | 0, 60 | 642 | 8.7 (37.1) | 0,674 |
| Tuesday | 237 | 22.0 (45.8) | 1,512 | 63 | 2.4 (2.0) | 0,8 | 89 | 5.0 (11.2) | 0,102 | 143 | 4.6 (5.4) | 0, 32 | 246 | 9.8 (34.8) | 0,493 |
| Wednesday | 222 | 19.7 (23.8) | 1,158 | 54 | 2.3 (2.5) | 0,13 | 293 | 3.3 (4.4) | 0,42 | 87 | 5.6 (11.9) | 0, 80 | 172 | 7.2 (15.5) | 0,109 |
| Thursday | 179 | 20.2 (27.8) | 1,260 | 914 | 2.1 (1.9) | 0,16 | 613 | 3.4 (5.5) | 0,93 | 489 | 5.7 (15.8) | 0,186 | 320 | 4.5 (15.6) | 0,216 |
| Friday | 186 | 21.8 (22.7) | 1,162 | 144 | 2.1 (1.8) | 0,8 | 117 | 4.4 (4.5) | 0,32 | 132 | 10.0 (46.0) | 0,500 | 196 | 5.9 (19.1) | 0,238 |
| Saturday | 152 | 19.5 (24.5) | 1,215 | 88 | 2.0 (1.6) | 0,8 | 63 | 4.2 (12.6) | 0,101 | 66 | 2.9 (4.1) | 0, 33 | 81 | 5.6 (11.7) | 0, 95 |
| Sunday | 117 | 20.9 (21.9) | 1,96 | 247 | 1.9 (1.9) | 0,12 | 121 | 2.5 (2.8) | 0,21 | 36 | 8.8 (20.7) | 0, 99 | 233 | 5.1 (19.1) | 0,198 |

2.2.b. Table 2.2b. Number of recorded physical contacts per participant per day stratified by age, gender, household size, and day of the week in the COVIMOD survey Waves 1 to 4 and the POLYMOD survey for the unweighted analysis including group contacts.

| | POLYMOD | | | COVIMOD | | | | | | | | | | | |
|---|---|---|---|---|---|---|---|---|---|---|---|---|---|---|---|
| | | | | Wave1 | | | Wave 2 | | | Wave 3 | | | Wave 4 | | |
| | N | Mean (SD) | Min, Max | N | Mean (SD) | Min, Max | N | Mean (SD) | Min, Max | N | Mean (SD) | Min, Max | N | Mean (SD) | Min, Max |
| | **1341** | **5.5 (4.8)** | **0,45** | **1560** | **0.9 (1.3)** | **0,11** | **1356** | **1.1 (3.2)** | **0,101** | **1081** | **1.3 (4.7)** | **0,129** | **1890** | **2.0 (16.3)** | **0,674** |
| **Age category** | | | | | | | | | | | | | | | |
| 0 - 4 | 89 | 7.3 (5.0) | 0,30 | 46 | 3.3 (1.9) | 0,7 | 36 | 3.3 (2.2) | 0,8 | 21 | 4.5 (4.7) | 0,20 | 56 | 5.2 (7.6) | 0,51 |
| 5 - 9 | 92 | 6.6 (4.4) | 0,33 | 48 | 2.6 (1.7) | 0,6 | 41 | 2.7 (1.8) | 0,6 | 30 | 3.8 (4.5) | 0,22 | 62 | 3.3 (3.3) | 0,23 |
| 10 - 14 | 110 | 6.4 (4.7) | 0,25 | 73 | 2.1 (1.9) | 0,11 | 63 | 1.7 (1.8) | 0,6 | 45 | 3.6 (6.4) | 0,33 | 87 | 3.5 (5.1) | 0,32 |
| 15 - 19 | 121 | 6.7 (6.9) | 0,45 | 95 | 1.4 (1.6) | 0,8 | 66 | 1.3 (1.6) | 0,6 | 45 | 1.5 (2.3) | 0,14 | 108 | 2.9 (8.5) | 0,83 |
| 20 - 24 | 117 | 5.2 (4.1) | 0,26 | 83 | 0.8 (1.1) | 0,5 | 60 | 0.7 (1.0) | 0,3 | 28 | 0.7 (0.9) | 0,3 | 109 | 2.4 (6.1) | 0,45 |
| 25 - 34 | 132 | 5.4 (3.8) | 0,29 | 173 | 0.8 (1.0) | 0,6 | 148 | 0.7 (0.9) | 0,5 | 96 | 1.2 (2.9) | 0,21 | 219 | 2.1 (8.9) | 0,120 |
| 35 - 44 | 156 | 5.8 (4.9) | 0,36 | 137 | 0.7 (1.2) | 0,8 | 124 | 0.8 (1.2) | 0,6 | 91 | 2.5 (13.5) | 0,129 | 164 | 4.8 (52.6) | 0,674 |
| 45 - 54 | 184 | 5.2 (4.8) | 0,35 | 235 | 0.7 (1.0) | 0,4 | 209 | 1.5 (7.2) | 0,101 | 174 | 1.1 (2.3) | 0,22 | 275 | 1.2 (3.9) | 0,52 |
| 55 - 64 | 160 | 4.0 (3.2) | 0,16 | 265 | 0.7 (0.9) | 0,5 | 244 | 0.8 (1.1) | 0,10 | 237 | 0.8 (1.1) | 0,5 | 321 | 1.1 (3.8) | 0,51 |
| 65 - 69 | 74 | 4.6 (6.3) | 0,40 | 270 | 0.6 (0.9) | 0,5 | 245 | 0.9 (2.5) | 0,29 | 199 | 1.0 (2.1) | 0,25 | 313 | 1.0 (2.1) | 0,28 |
| 70 - 74 | 33 | 2.6 (3.0) | 0,13 | 89 | 0.7 (0.8) | 0,3 | 73 | 0.8 (0.9) | 0,3 | 79 | 0.9 (1.3) | 0,8 | 118 | 0.8 (1.7) | 0,17 |
| 75 - 79 | 14 | 2.7 (2.6) | 0,8 | 35 | 0.8 (0.9) | 0,3 | 34 | 0.6 (1.0) | 0,3 | 29 | 0.8 (0.9) | 0,3 | 48 | 0.8 (0.9) | 0,3 |
| 80 + | 13 | 2.8 (1.8) | 1,6 | 11 | 0.3 (0.5) | 0,1 | 11 | 0.2 (0.4) | 0,1 | 7 | 0.3 (0.8) | 0,2 | 10 | 0.3 (0.5) | 0,1 |
| **Sex of participants** | | | | | | | | | | | | | | | |
| Female | 722 | 5.3 (4.8) | 0,45 | 748 | 0.9 (1.3) | 0,8 | 638 | 1.3 (4.3) | 0,101 | 536 | 1.1 (2.1) | 0,25 | 901 | 1.4 (3.3) | 0,52 |
| Male | 581 | 5.6 (4.9) | 0,44 | 806 | 0.9 (1.3) | 0,11 | 717 | 0.9 (1.7) | 0,29 | 544 | 1.6 (6.2) | 0,129 | 985 | 2.5 (22.3) | 0,674 |
| **Household size** | | | | | | | | | | | | | | | |
| 1 | 250 | 4.0 (4.4) | 0,36 | 232 | 0.1 (0.5) | 0,5 | 256 | 0.2 (0.6) | 0,4 | 268 | 0.8 (7.9) | 0,129 | 487 | 2.0 (30.7) | 0,674 |
| 2 | 411 | 4.9 (4.5) | 0,45 | 412 | 0.5 (0.7) | 0,6 | 351 | 1.1 (5.8) | 0,101 | 270 | 1.0 (2.3) | 0,25 | 439 | 1.1 (2.9) | 0,38 |
| 3 | 339 | 5.8 (4.2) | 0,30 | 514 | 0.9 (1.0) | 0,8 | 447 | 1.1 (1.2) | 0,13 | 343 | 1.3 (1.8) | 0,14 | 544 | 1.7 (5.8) | 0,120 |
| 4 or more | 341 | 6.7 (5.6) | 0,44 | 402 | 1.8 (1.8) | 0,11 | 302 | 1.8 (2.1) | 0,13 | 200 | 2.6 (4.4) | 0,33 | 420 | 3.2 (6.9) | 0,83 |
| **Weekdays** | | | | | | | | | | | | | | | |
| Monday | 227 | 5.1 (4.2) | 0,29 | 50 | 1.4 (1.6) | 0,6 | 60 | 1.1 (1.4) | 0,6 | 128 | 1.3 (2.4) | 0,20 | 642 | 2.8 (26.9) | 0,674 |
| Tuesday | 237 | 5.3 (4.4) | 0,40 | 63 | 1.0 (1.5) | 0,7 | 89 | 1.2 (1.8) | 0,13 | 143 | 1.2 (2.4) | 0,22 | 246 | 2.6 (9.4) | 0,120 |
| Wednesday | 222 | 5.6 (5.1) | 0,36 | 54 | 0.7 (1.2) | 0,6 | 293 | 0.8 (1.7) | 0,22 | 87 | 1.5 (3.9) | 0,33 | 172 | 1.9 (4.7) | 0,45 |
| Thursday | 179 | 5.4 (4.6) | 0,30 | 914 | 0.8 (1.2) | 0,8 | 613 | 1.0 (1.8) | 0,29 | 489 | 1.1 (2.2) | 0,25 | 320 | 1.0 (1.5) | 0,11 |
| Friday | 186 | 5.6 (4.2) | 0,23 | 144 | 1.0 (1.3) | 0,6 | 117 | 1.2 (1.9) | 0,12 | 132 | 2.5 (11.7) | 0,129 | 196 | 1.1 (2.4) | 0,23 |
| Saturday | 152 | 6.0 (5.4) | 0,44 | 88 | 1.1 (1.6) | 0,8 | 63 | 2.7 (12.7) | 0,101 | 66 | 0.9 (1.2) | 0,5 | 81 | 1.6 (2.3) | 0,13 |
| Sunday | 117 | 6.0 (6.7) | 0,45 | 247 | 1.0 (1.5) | 0,11 | 121 | 1.0 (1.3) | 0,5 | 36 | 0.9 (1.2) | 0,5 | 233 | 1.5 (6.0) | 0,83 |

2.2.c. Table 2.2c. Number of recorded overall contacts per different settings in the COVIMOD survey Waves 1 to 4 and the POLYMOD survey for the unweighted analysis including group contacts.

**Note:** *The displayed educational contacts are based on the group of participants who attended an educational facility (kindergarten, school, university) and work contacts are based on the group of participants who reported to work full-/part-time.*

| | POLYMOD | | | COVIMOD | | | | | | | | | | | |
|---|---|---|---|---|---|---|---|---|---|---|---|---|---|---|---|
| | | | | Wave1 | | | Wave2 | | | Wave3 | | | Wave4 | | |
| | N | Mean (SD) | Min, Max | N | Mean (SD) | Min, Max | N | Mean (SD) | Min, Max | N | Mean (SD) | Min, Max | N | Mean (SD) | Min, Max |
| Overall | 1341 | 20.2 (28.5) | 1,512 | 1560 | 2.1 (1.9) | 0,16 | 1356 | 3.6 (6.1) | 0,102 | 1081 | 5.9 (20.3) | 0,500 | 1890 | 7.1 (27.9) | 0,674 |
| Home | 1341 | 2.8 (2.3) | 0,26 | 1560 | 1.6 (1.5) | 0,12 | 1356 | 1.6 (1.6) | 0,9 | 1081 | 1.5 (1.5) | 0,13 | 1890 | 1.5 (1.7) | 0,23 |
| Educational | 199 | 2.8 (4.7) | 0,42 | 310 | 0.0 (0.2) | 0,3 | 247 | 0.2 (1.0) | 0,10 | 179 | 0.3 (1.9) | 0,17 | 385 | 1.2 (4.3) | 0,60 |
| Work | 715 | 18.5 (33.2) | 0,509 | 690 | 0.4 (1.2) | 0,11 | 613 | 2.0 (7.2) | 0,100 | 476 | 4.0 (15.1) | 0,140 | 809 | 4.6 (23.0) | 0,491 |
| Transport | 1341 | 0.3 (0.8) | 0,8 | 1560 | 0.0 (0.3) | 0,3 | 1356 | 0.1 (0.4) | 0,4 | 1081 | 0.1 (0.4) | 0,8 | 1890 | 0.1 (0.6) | 0,12 |
| Others | 1341 | 3.0 (3.8) | 0,45 | 1560 | 0.4 (1.0) | 0,10 | 1356 | 1.1 (2.4) | 0,33 | 1081 | 1.9 (6.7) | 0,149 | 1890 | 2.8 (21.2) | 0,674 |

2.2.d. Table 2.2d. Number of recorded physical contacts per different settings in the COVIMOD survey Waves 1 to 4 and the POLYMOD survey for the unweighted analysis including group contacts.

**Note:** *The displayed educational contacts are based on the group of participants who attended an educational facility (kindergarten, school, university) and work contacts are based on the group of participants who reported to work full-/part-time.*

| | POLYMOD | | | COVIMOD | | | | | | | | | | | |
|---|---|---|---|---|---|---|---|---|---|---|---|---|---|---|---|
| | | | | Wave1 | | | Wave2 | | | Wave3 | | | Wave4 | | |
| | N | Mean (SD) | Min, Max | N | Mean (SD) | Min, Max | N | Mean (SD) | Min, Max | N | Mean (SD) | Min, Max | N | Mean (SD) | Min, Max |
| Overall | 1341 | 5.5 (4.8) | 0,45 | 1560 | 0.9 (1.3) | 0,11 | 1356 | 1.1 (3.2) | 0,101 | 1081 | 1.3 (4.7) | 0,129 | 1890 | 2.0 (16.3) | 0,674 |
| Home | 1341 | 2.3 (2.2) | 0,26 | 1560 | 0.8 (1.3) | 0,11 | 1356 | 0.8 (1.2) | 0, 8 | 1081 | 0.8 (1.2) | 0, 12 | 1890 | 0.8 (1.2) | 0, 8 |
| Educational | 199 | 1.5 (3.8) | 0,42 | 310 | 0.0 (0.1) | 0,1 | 247 | 0.0 (0.3) | 0, 4 | 179 | 0.1 (0.3) | 0, 3 | 385 | 0.3 (1.8) | 0, 21 |
| Work | 715 | 1.4 (3.0) | 0,30 | 690 | 0.0 (0.2) | 0,2 | 613 | 0.4 (4.4) | 0,100 | 476 | 0.2 (1.7) | 0, 25 | 809 | 0.5 (3.2) | 0, 60 |
| Transport | 1341 | 0.2 (0.6) | 0,6 | 1560 | 0.0 (0.2) | 0,3 | 1356 | 0.0 (0.3) | 0, 3 | 1081 | 0.0 (0.2) | 0, 3 | 1890 | 0.0 (0.3) | 0, 7 |
| Others | 1341 | 2.1 (3.2) | 0,40 | 1560 | 0.1 (0.5) | 0,5 | 1356 | 0.2 (0.7) | 0, 11 | 1081 | 0.4 (4.1) | 0,129 | 1890 | 0.8 (15.7) | 0,674 |

### 2.3. Reproduction number estimates of SARS-CoV-2 under contact reduction measures

2.3.a. Table 2.3a. Effective reproduction number at the timing of COVIMOD survey Waves 1 to 4, estimated effective reproduction number based on the reduction of social contacts at the times of COVIMOD survey Waves 1 to 4 assuming values of the basic reproduction number of Norm (2.6, SD=0.54) and the reduction in mobility at the times of the COVIMOD survey Waves, with 10000 bootstrapped samples in various settings and in different time frames for the unweighted analysis including group contacts.

**Note:** *RKI - the percentage mean and minimum and maximum reductions in the reproduction number in the assessed time intervals, COVIMOD - the calculated percentage mean reductions in the reproduction number and the 95% confidence interval, Google and Apple mobility – the percentage mean and minimum and maximum reduction in mobility in the assessed time intervals.*

| | COVIMOD | | RKI | | Google | | Apple | |
|---|---|---|---|---|---|---|---|---|
| | Mean reproduction number (Min, Max) | Percent Mean Reduction Number (Min, Max) | Mean reproduction number (Min, Max) | Percent Mean Reduction Number (Min, Max) | Mean reproduction number (Min, Max) | Percent Mean Reduction Number (Min, Max) | Mean reproduction number (Min, Max) | Percent Mean Reduction Number (Min, Max) |
| **Overall** | | | | | | | | |
| 30th April - 6th May 2020 | 0.29 (0.18, 0.42) | -88.00 (-93.00, -83.00) | 0.88 (0.79, 0.97) | -66.00 (-69.00, -62.00) | - | -24.31 (-59.40, -15.20) | - | -36.77 (-44.17, -29.61) |
| 14th May - 21st May 2020 | 0.42 (0.25, 0.59) | -83.00 (-90.00, -77.00) | 0.90 (0.72, 1.15) | -65.00 (-72.00, -55.00) | - | -16.85 (-50.20, -3.40) | - | -15.64 (-25.26, -2.58) |
| 28th May - 4th June 2020 | 0.61 (0.35, 0.89) | -76.00 (-86.00, -65.00) | 1.06 (0.84, 1.39) | -59.00 (-67.00, -46.00) | - | -13.05 (-47.40, 1.60) | - | 5.18 (-4.00, 19.43) |
| 11th June - 22nd June 2020 | 0.73 (0.43, 1.05) | -71.00 (-83.00, -59.00) | 0.84 (0.60, 1.07) | -67.00 (-76.00, -58.00) | - | -10.63 (-34.60, 5.60) | - | 11.66 (1.62, 28.00) |
| **Home** | | | | | | | | |
| 30th April - 6th May 2020 | 1.60 (0.95, 2.28) | -38.00 (-63.00, -12.00) | - | - | - | 12.71 (6.00, 28.00) | - | - |
| 14th May - 21st May 2020 | 1.59 (0.93, 2.26) | -38.00 (-64.00, -13.00) | - | - | - | 8.62 (3.00, 18.00) | - | - |
| 28th May - 4th June 2020 | 1.57 (0.90, 2.26) | -39.00 (-65.00, -13.00) | - | - | - | 6.88 (0.00, 16.00) | - | - |
| 11th June - 22nd June 2020 | 1.42 (0.84, 2.02) | -45.00 (-67.00, -22.00) | - | - | - | 5.25 (-1.00, 16.00) | - | - |
| **Educational** | | | | | | | | |
| 30th April - 6th May 2020 | 0.07 (0.02, 0.16) | -97.00 (-99.00, -93.00) | - | - | - | - | - | - |
| 14th May - 21st May 2020 | 0.34 (0.12, 0.77) | -86.00 (-95.00, -70.00) | - | - | - | - | - | - |
| 28th May - 4th June 2020 | 0.51 (0.16, 1.22) | -80.00 (-93.00, -53.00) | - | - | - | - | - | - |
| 11th June - 22nd June 2020 | 1.26 (0.60, 2.22) | -51.00 (-76.00, -14.00) | - | - | - | - | - | - |
| **Work** | | | | | | | | |
| 30th April - 6th May 2020 | 0.05 (0.03, 0.07) | -98.00 (-98.00, -97.00) | - | - | - | -34.71 (-84.00, -18.00) | - | - |
| 14th May - 21st May 2020 | 0.16 (0.09, 0.24) | -93.00 (-96.00, -90.00) | - | - | - | -27.00 (-81.00, -1.00) | - | - |
| 28th May - 4th June 2020 | 0.32 (0.17, 0.51) | -87.00 (-93.00, -80.00) | - | - | - | -24.12 (-80.00, 8.00) | - | - |
| 11th June - 22nd June 2020 | 0.40 (0.22, 0.61) | -84.00 (-91.00, -76.00) | - | - | - | -16.50 (-59.00, 15.00) | - | - |
| **Transport** | | | | | | | | |
| 30th April - 6th May 2020 | 0.64 (0.30, 1.12) | -75.00 (-88.00, -56.00) | - | - | - | -42.14 (-67.00, -35.00) | - | - |
| 14th May - 21st May 2020 | 0.90 (0.43, 1.61) | -65.00 (-83.00, -38.00) | - | - | - | -31.75 (-46.00, -25.00) | - | - |
| 28th May - 4th June 2020 | 0.71 (0.32, 1.31) | -72.00 (-87.00, -49.00) | - | - | - | -25.12 (-42.00, -13.00) | - | - |
| 11th June - 22nd June 2020 | 1.17 (0.53, 2.30) | -55.00 (-79.00, -11.00) | - | - | - | -23.00 (-40.00, -11.00) | - | - |
| **Others** | | | | | | | | |
| 30th April - 6th May 2020 | 0.40 (0.23, 0.65) | -84.00 (-91.00, -75.00) | - | - | - | -28.71 (-87.00, -7.50) | - | - |
| 14th May - 21st May 2020 | 0.72 (0.42, 1.04) | -72.00 (-83.00, -60.00) | - | - | - | -17.06 (-71.00, 14.00) | - | - |
| 28th May - 4th June 2020 | 1.41 (0.77, 2.21) | -45.00 (-70.00, -15.00) | - | - | - | -11.44 (-65.50, 6.50) | - | - |
| 11th June - 22nd June 2020 | 1.53 (0.89, 2.27) | -41.00 (-65.00, -12.00) | - | - | - | -9.46 (-45.00, 12.50) | - | - |

2.4. **Figure 2.4. Boxplots of the number of overall contacts during the POLYMOD and COVIMOD survey Waves 1 to 4 in various settings. The boundaries of the boxes closest to zero indicates the 25th percentiles, the lines within the boxes marks the medians, the red dots within the boxes marks the means, and the boundary of the boxes farthest from zero indicates the 75th percentiles. Whiskers above and below the boxes indicate the 10th and 90th percentiles; black dots represent outliers. Participants with no contacts are displayed as 0 on the log-scale of the y-axis.**

2.4.a. Figure 2.4a. Displayed are (A) the overall number of contacts, (B) home contacts, (C) work contacts, (D) educational contacts, (E) public transport contacts and (F) other contacts for the unweighted analysis including group contacts in overall contacts.

**Note:** *The displayed educational contacts are based on the group of participants who attended an educational facility (kindergarten, school, university) and work contacts are based on the group of participants who reported to work full-/part-time.*

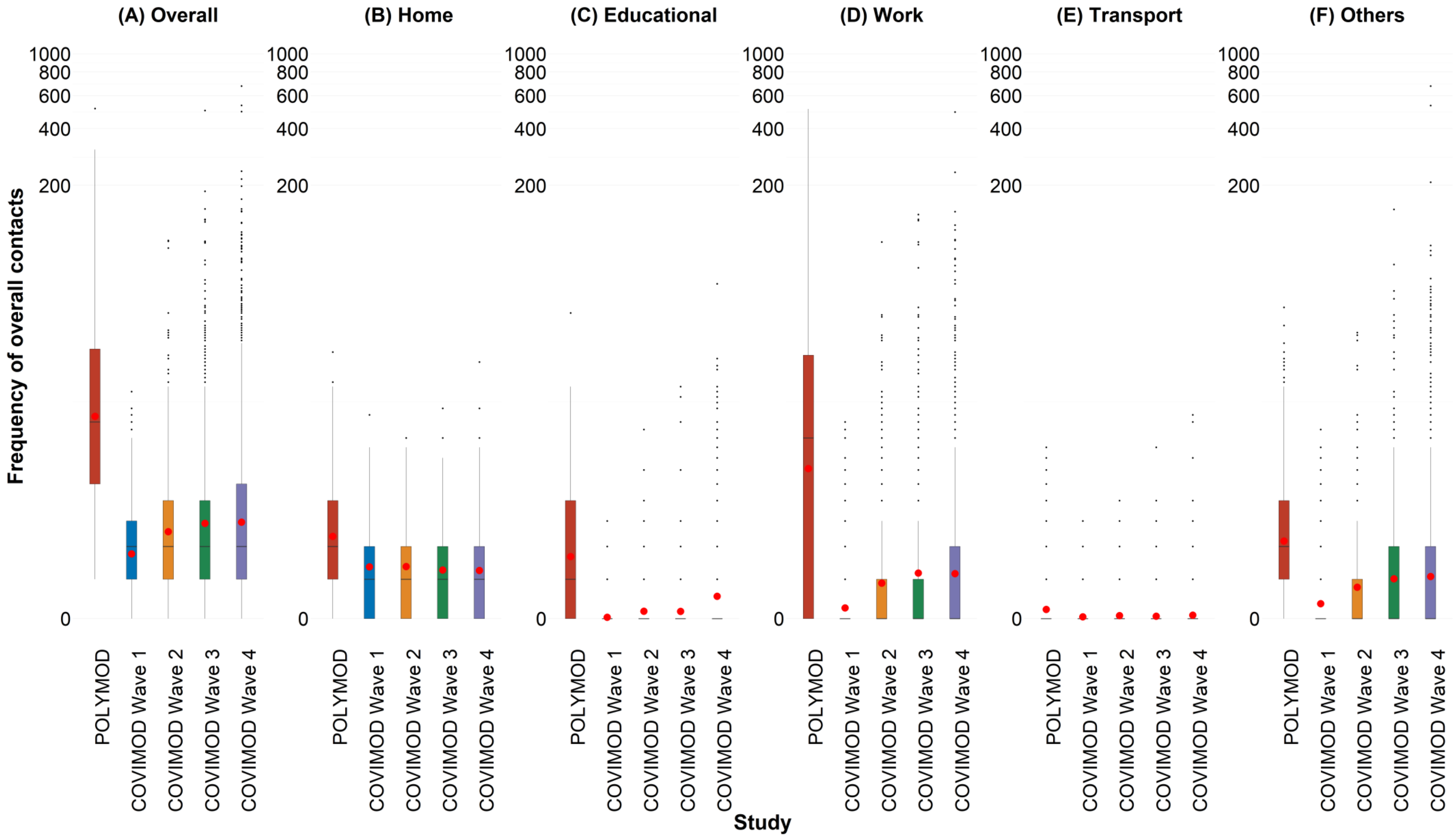

2.4.b. Figure 2.4b. Displayed are (A) 0 - 7 years, (B) 8 – 15 years, (C) 16 – 23 years, (D) 24 – 38 years, (E) 39 – 60 years and (F) 61 years or more for the unweighted analysis including group contacts in overall contacts.

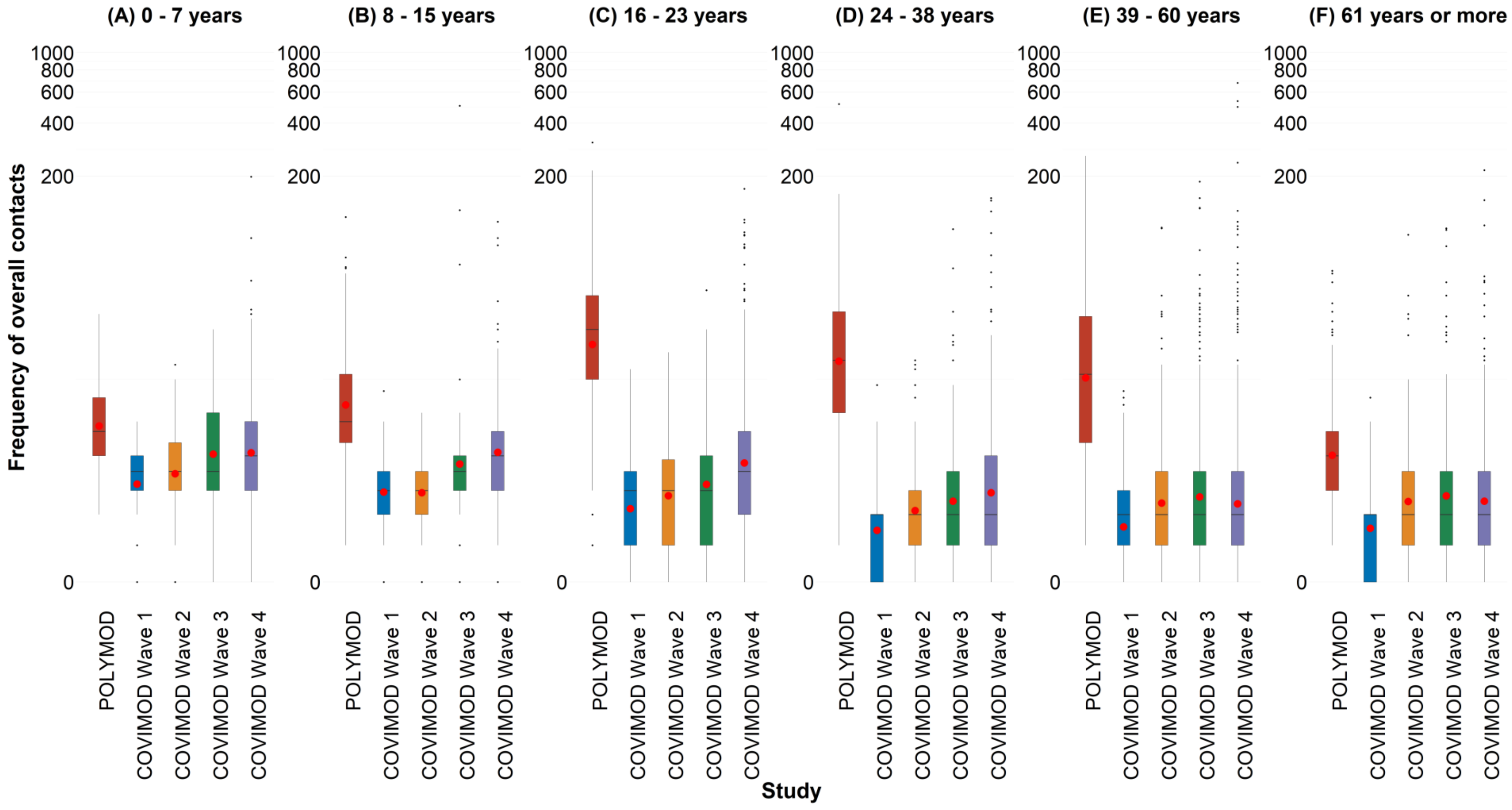

2.4.c. Figure 2.4c. Displayed are (A) Household size of 1, (B) Household size of 2, (C) Household size of 3 and (D) Household size of 4 or more for the unweighted analysis including group contacts in overall contacts.

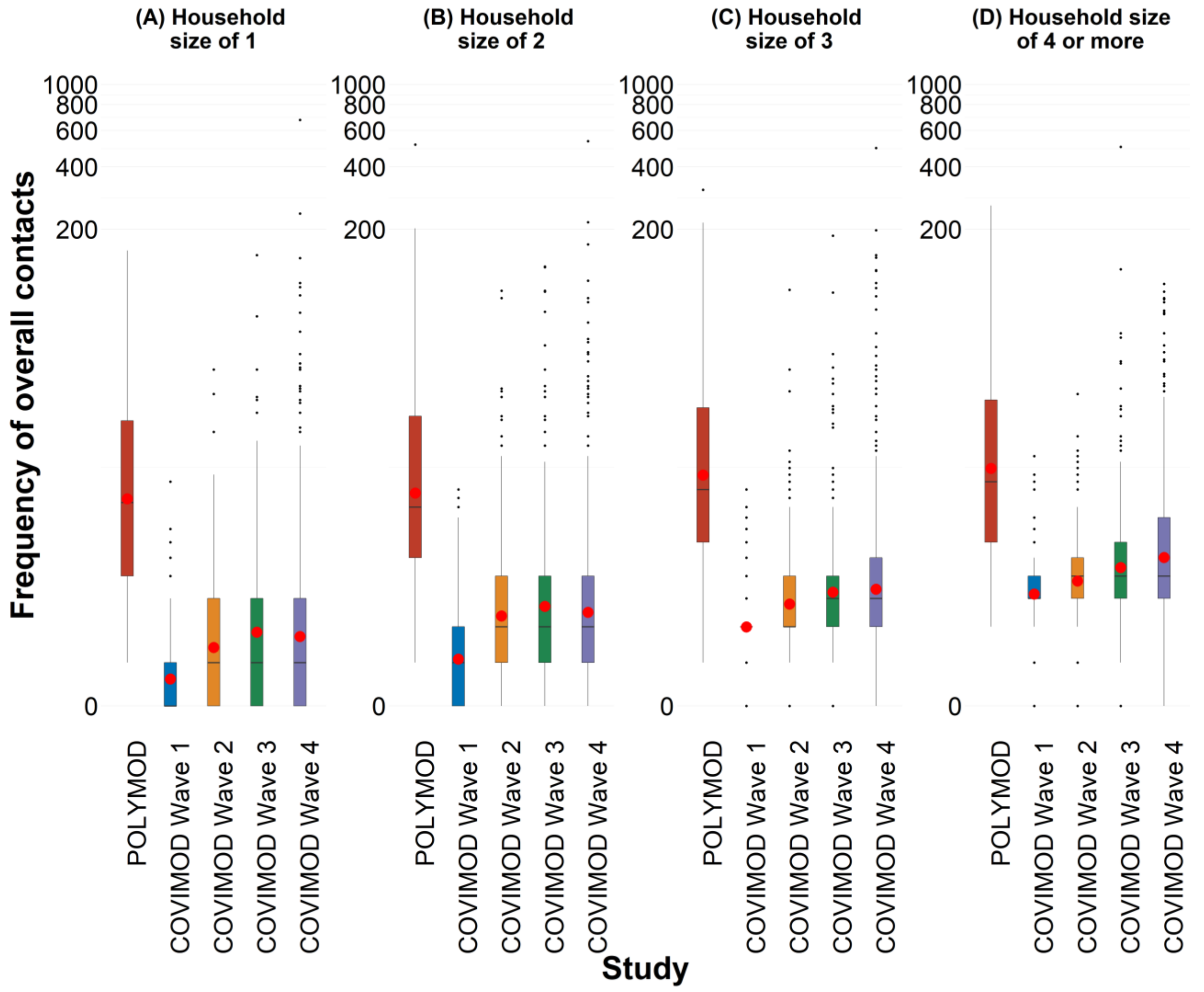

2.4.d. Figure 2.4d. Displayed are (A) Female and (B) Male for the unweighted analysis including group contacts in overall contacts.

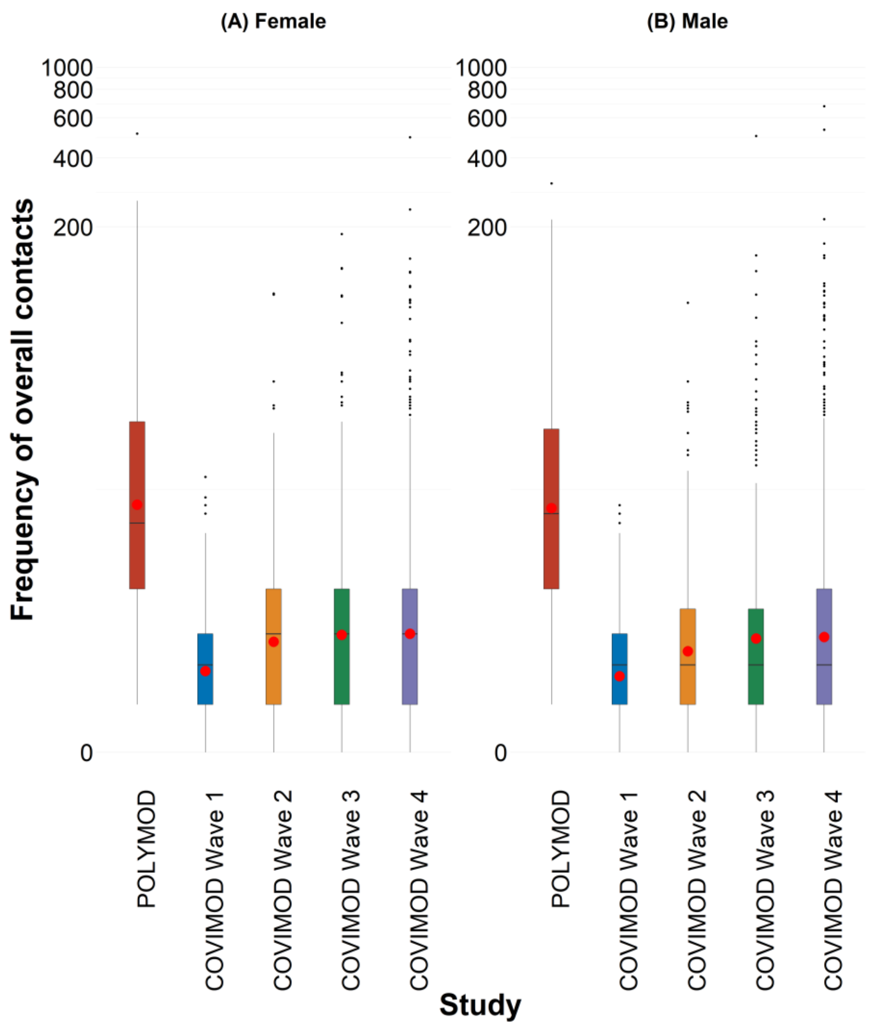

2.4.e. Figure 2.4e. Displayed are (A) Monday, (B) Tuesday, (C) Wednesday, (D) Thursday, (E) Friday, (F) Saturday and (G) Sunday for the unweighted analysis including group contacts in overall contacts.

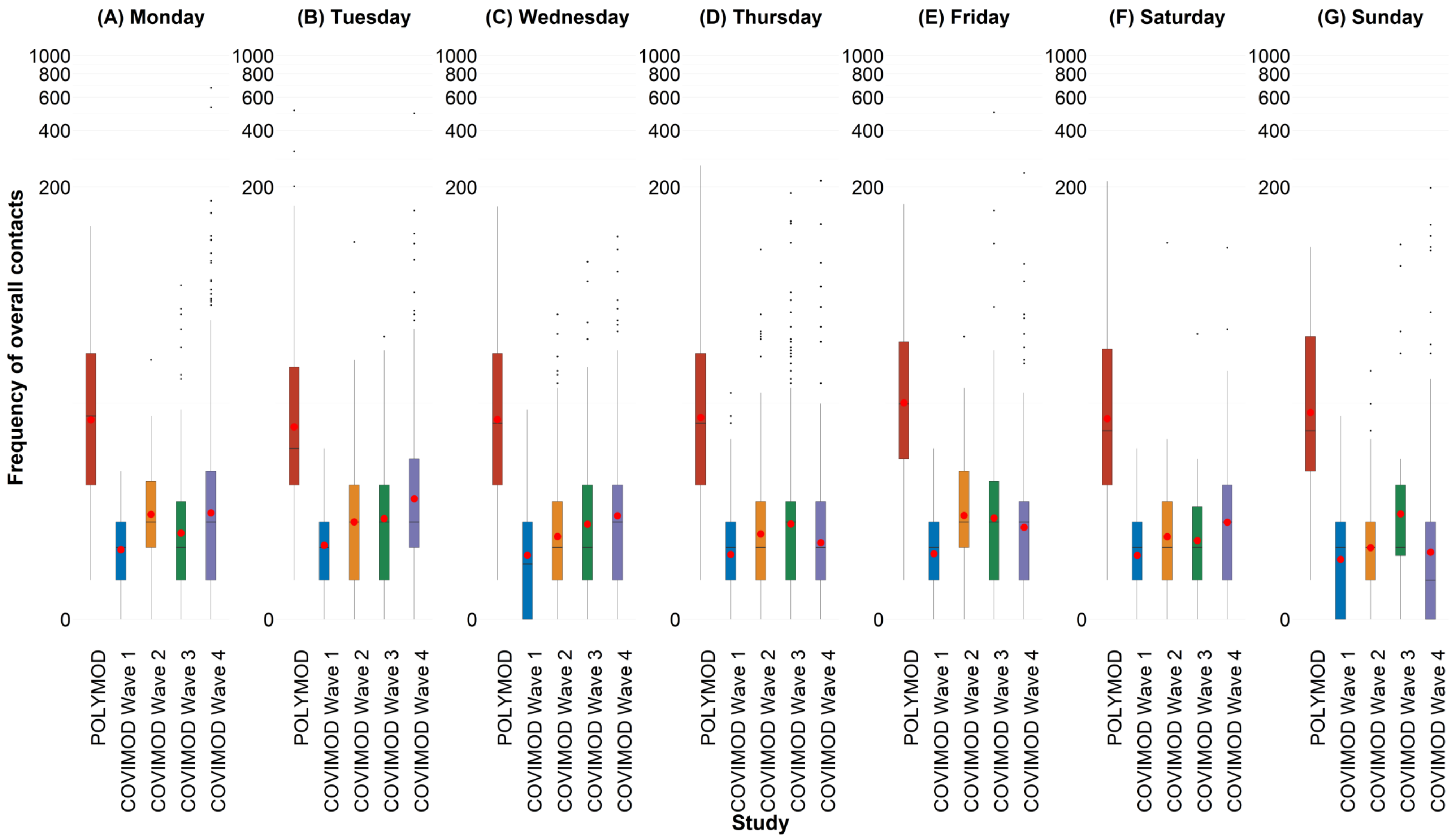

2.5. **Figure 2.5. Boxplots of the number of physical contacts during the POLYMOD and COVIMOD survey waves 1 to 4 in various settings. The boundaries of the boxes closest to zero indicates the 25th percentiles, the lines within the boxes marks the medians, the red dots within the boxes marks the means, and the boundary of the boxes farthest from zero indicates the 75th percentiles. Whiskers above and below the boxes indicate the 10th and 90th percentiles; black dots represent outliers. Participants with no contacts are displayed as 0 on the log-scale of the y-axis.**

2.5.a. Figure 2.5a. Displayed are (A) the overall number of contacts, (B) home contacts, (C) work contacts, (D) educational contacts, (E) public transport contacts and (F) other contacts for the unweighted analysis including group contacts in physical contacts.

**Note:** *The displayed educational contacts are based on the group of participants who attended an educational facility (kindergarten, school, university) and work contacts are based on the group of participants who reported to work full-/part-time.*

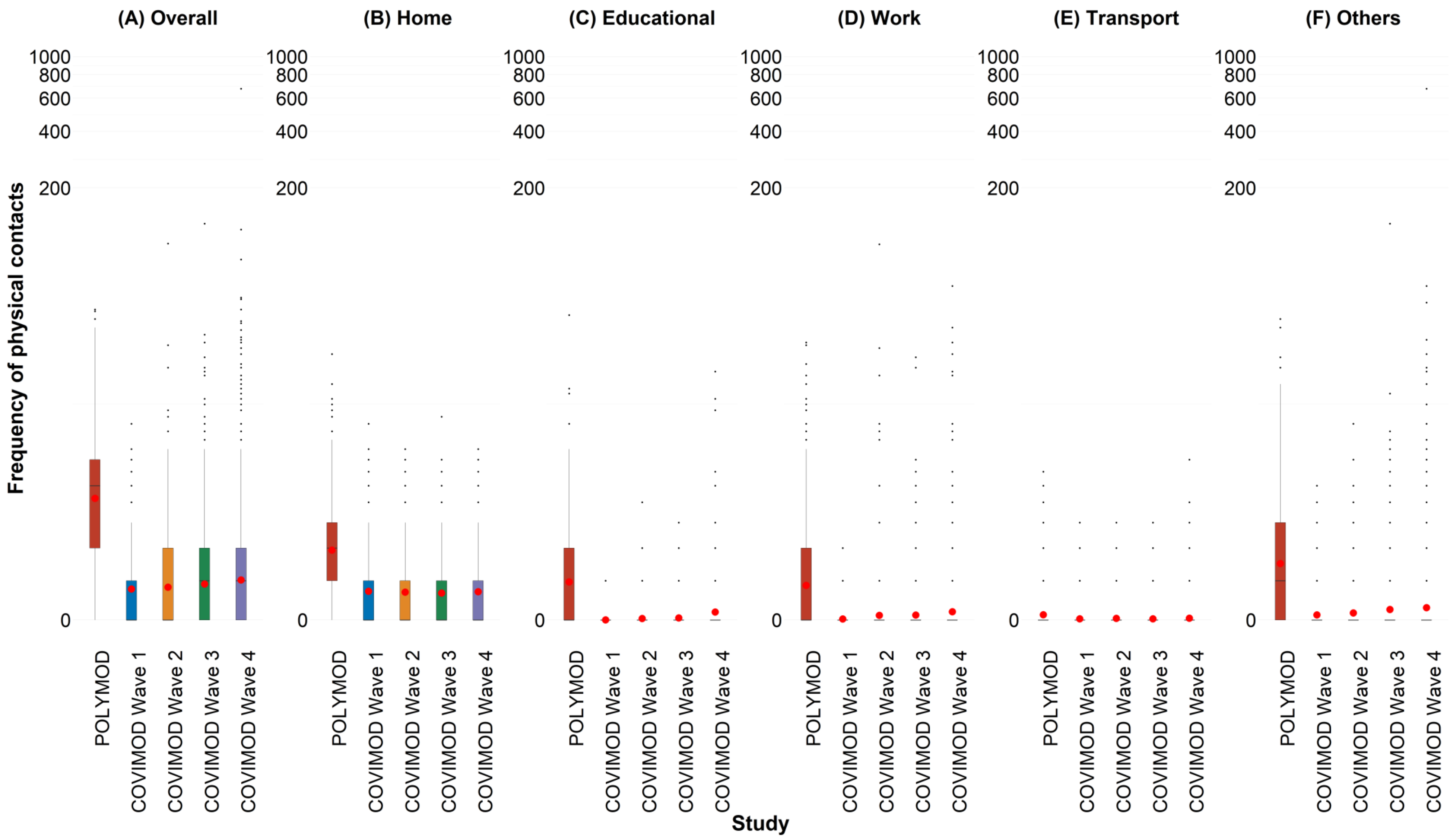

2.5.b. Figure 2.5b. Displayed are (A) 0 - 7 years, (B) 8 – 15 years, (C) 16 – 23 years, (D) 24 – 38 years, (E) 39 – 60 years and (F) 61 years or more for the unweighted analysis including group contacts in physical contacts.

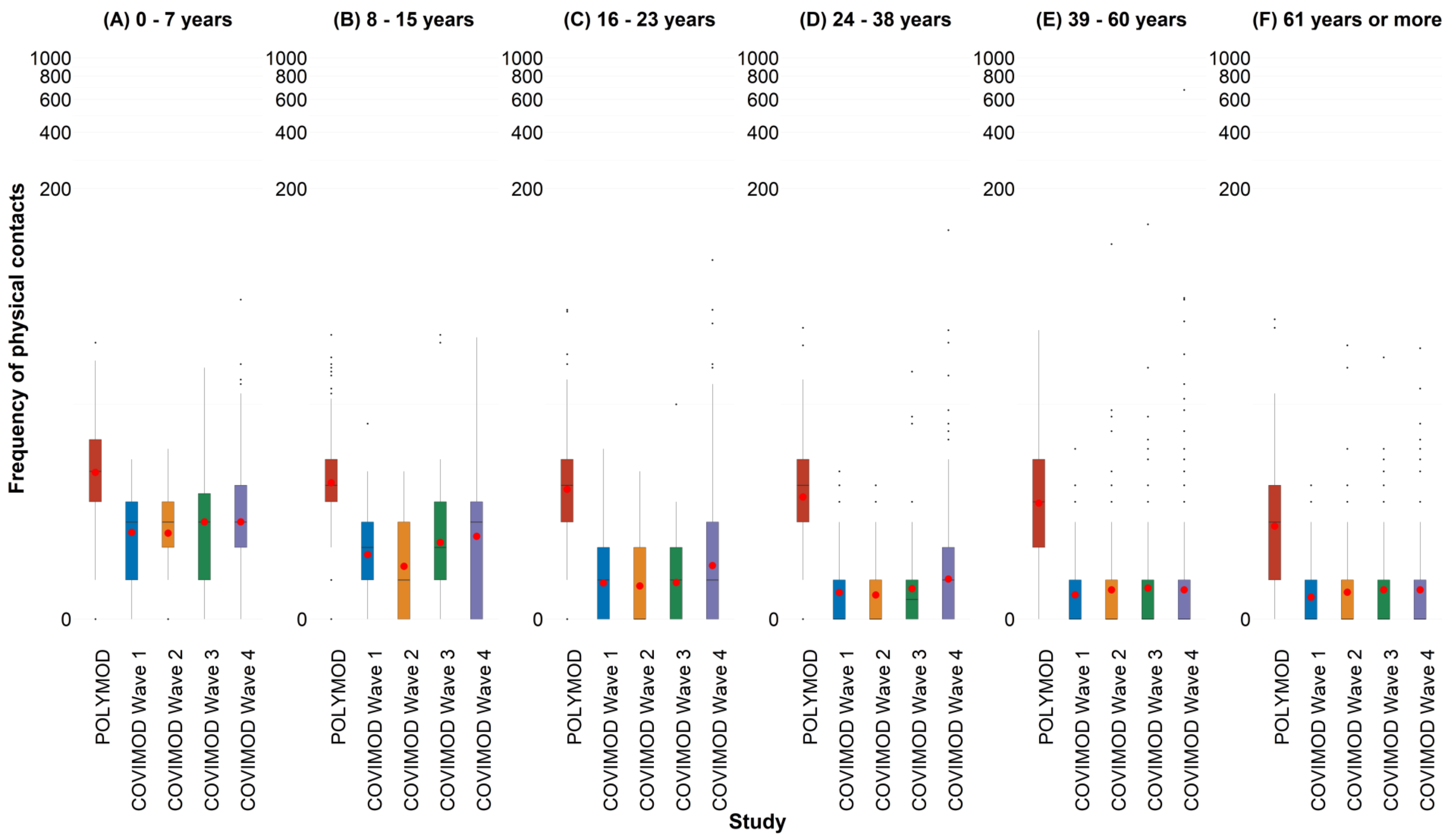

2.5.c. Figure 2.5c. Displayed are (A) Household size of 1, (B) Household size of 2, (C) Household size of 3 and (D) Household size of 4 or more for the unweighted analysis including group contacts in physical contacts.

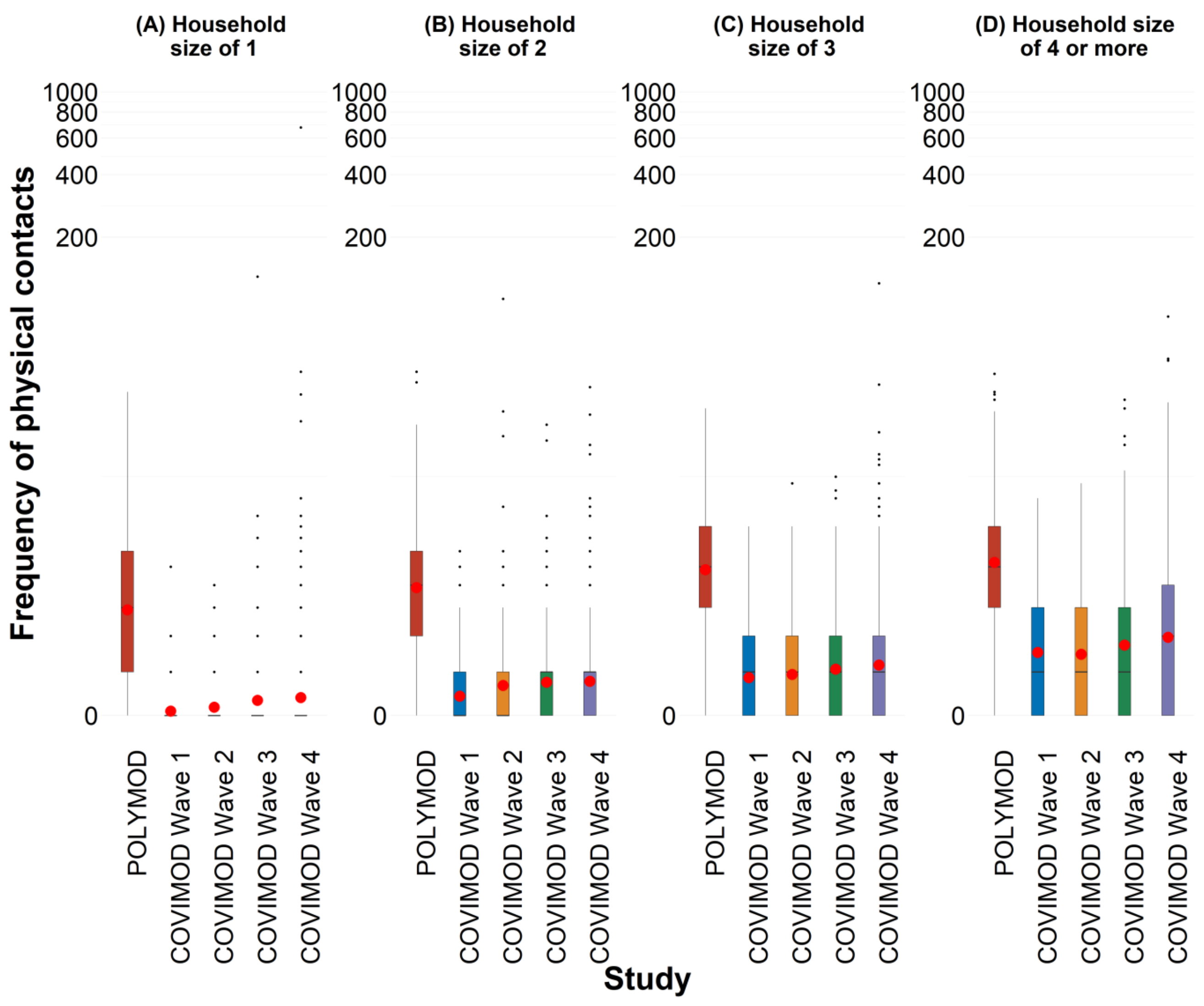

2.5.d. Figure 2.5d. Displayed are (A) Female and (B) Male for the unweighted analysis including group contacts in physical contacts.

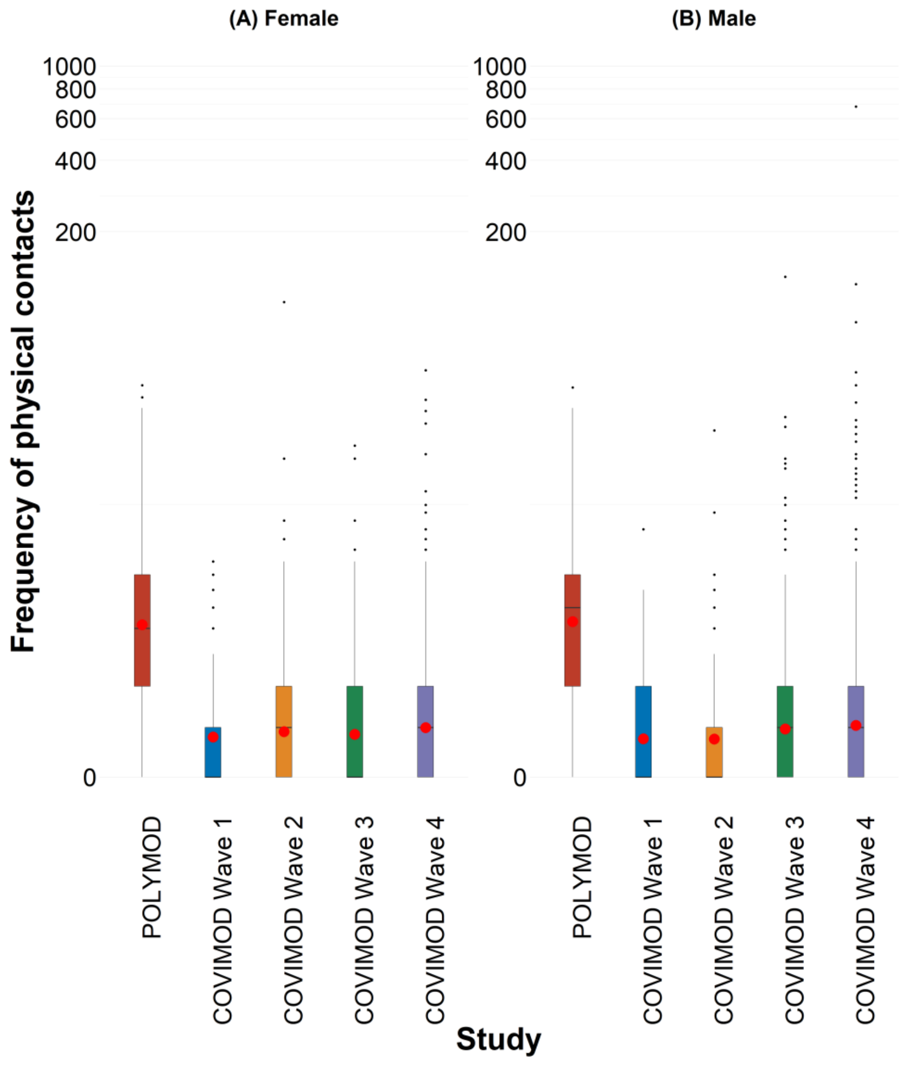

2.5.e. Figure 2.5e. Displayed are (A) Monday, (B) Tuesday, (C) Wednesday, (D) Thursday, (E) Friday, (F) Saturday and (G) Sunday for the unweighted analysis including group contacts in physical contacts.

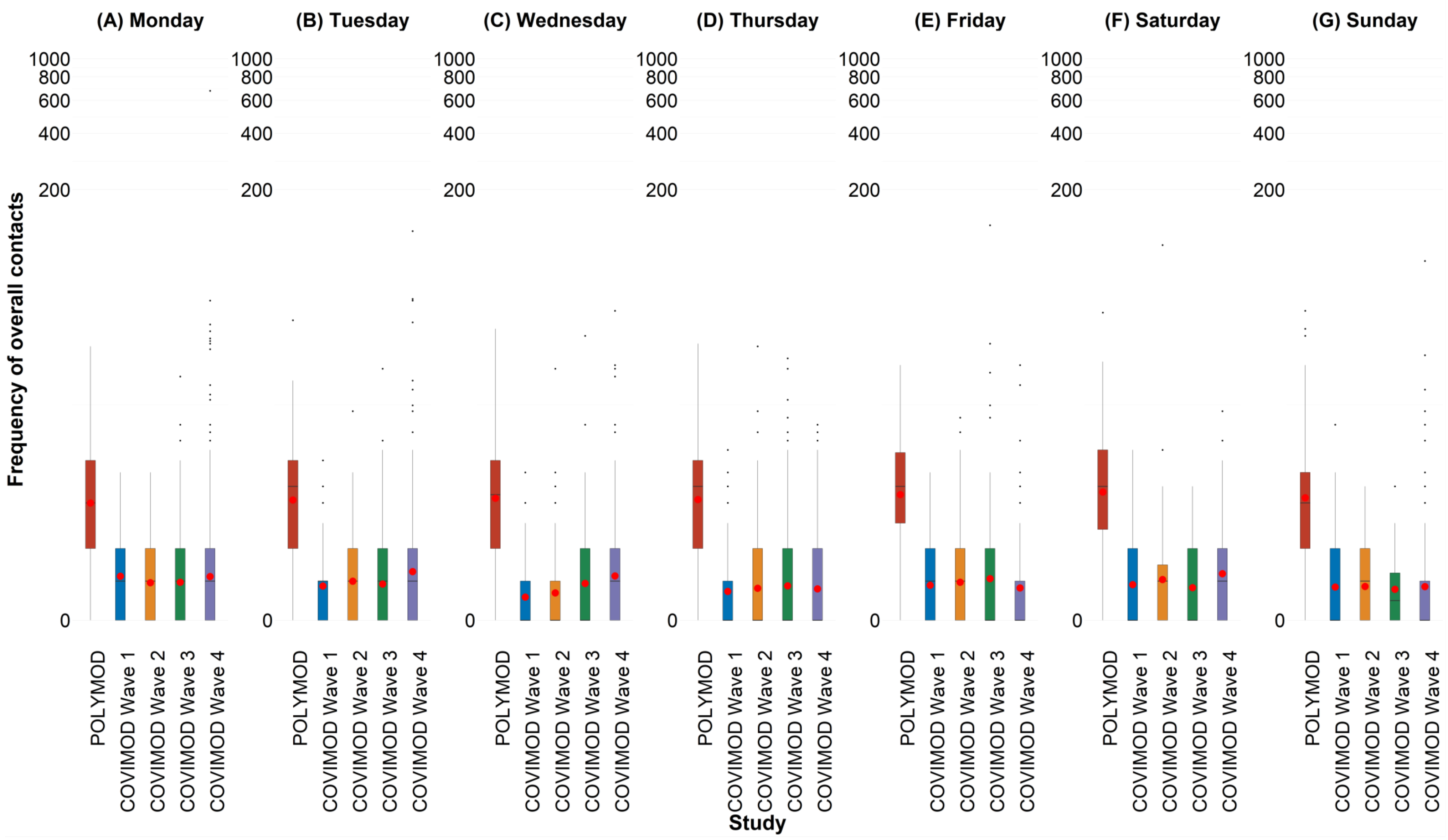

### 2.6. Social contact patterns.

2.6.a. Figure 2.6a. Social contact matrices with the mean total number of reported daily social contacts by participants in different age groups with individuals in other age groups in POLYMOD and COVIMOD survey waves 1 to 4 in various settings. Displayed are (A) the overall number of contacts, (B) home contacts, (C) educational contacts, (D) work contacts, (E) public transport contacts and (F) other contacts for the unweighted analysis including group contacts.

**Note:** *Participants with more than more than 100 group contacts were removed in COVIMOD and POLYMOD. Specifically, 6 participants and 13 participants were removed in Wave 3 and Wave 4 respectively, while 10 participants were removed in POLYMOD*

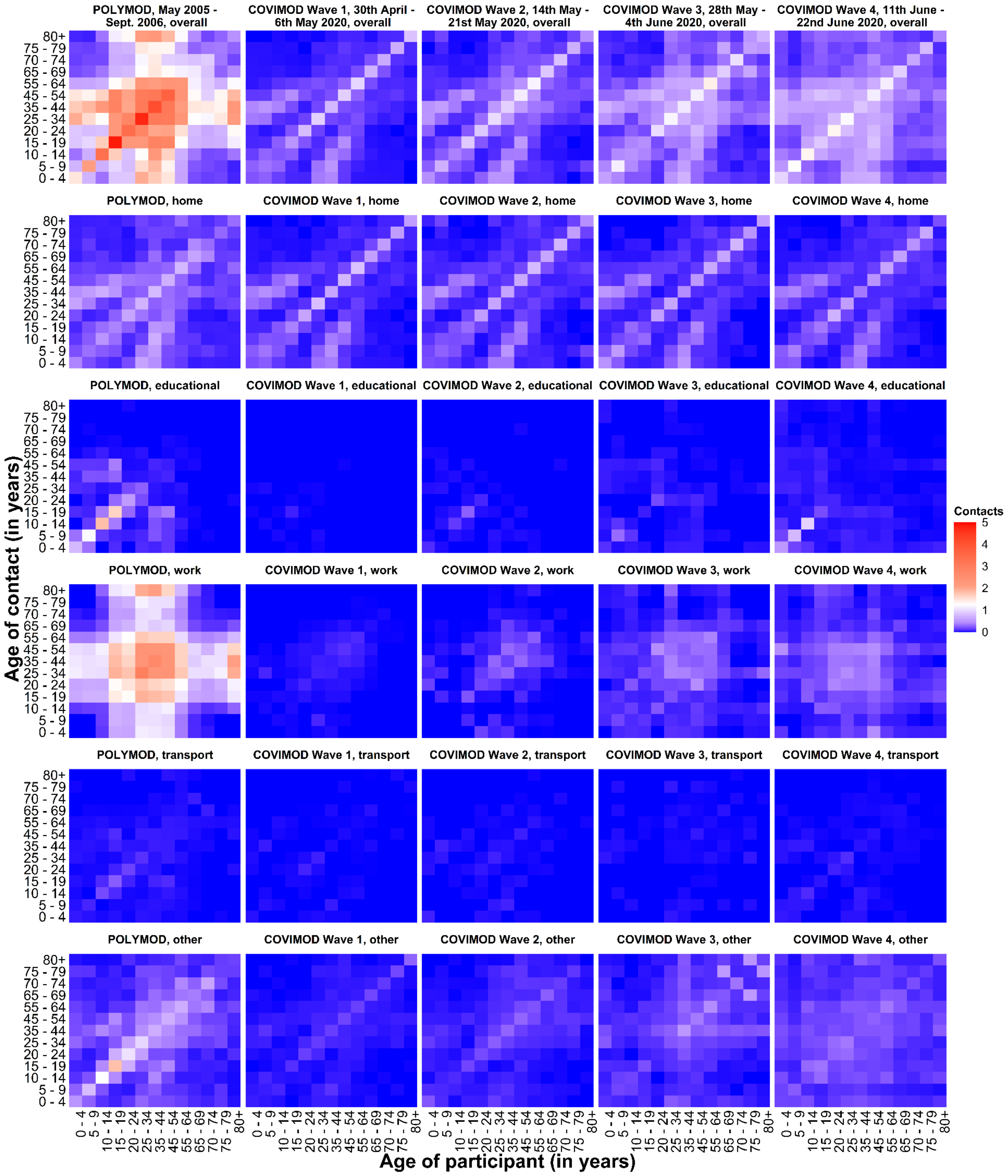

2.6.b. Figure 2.6b. Social contact matrices with the mean physical number of reported daily social contacts by participants in different age groups with individuals in other age groups in POLYMOD and COVIMOD survey waves 1 to 4 in various settings. Displayed are (A) the overall number of contacts, (B) home contacts, (C) educational contacts, (D) work contacts, (E) public transport contacts and (F) other contacts the unweighted analysis including group contacts.

**Note:** *Participants with more than more than 100 group contacts were removed in COVIMOD and POLYMOD. Specifically, 6 participants and 13 participants were removed in Wave 3 and Wave 4 respectively, while 10 participants were removed in POLYMOD*

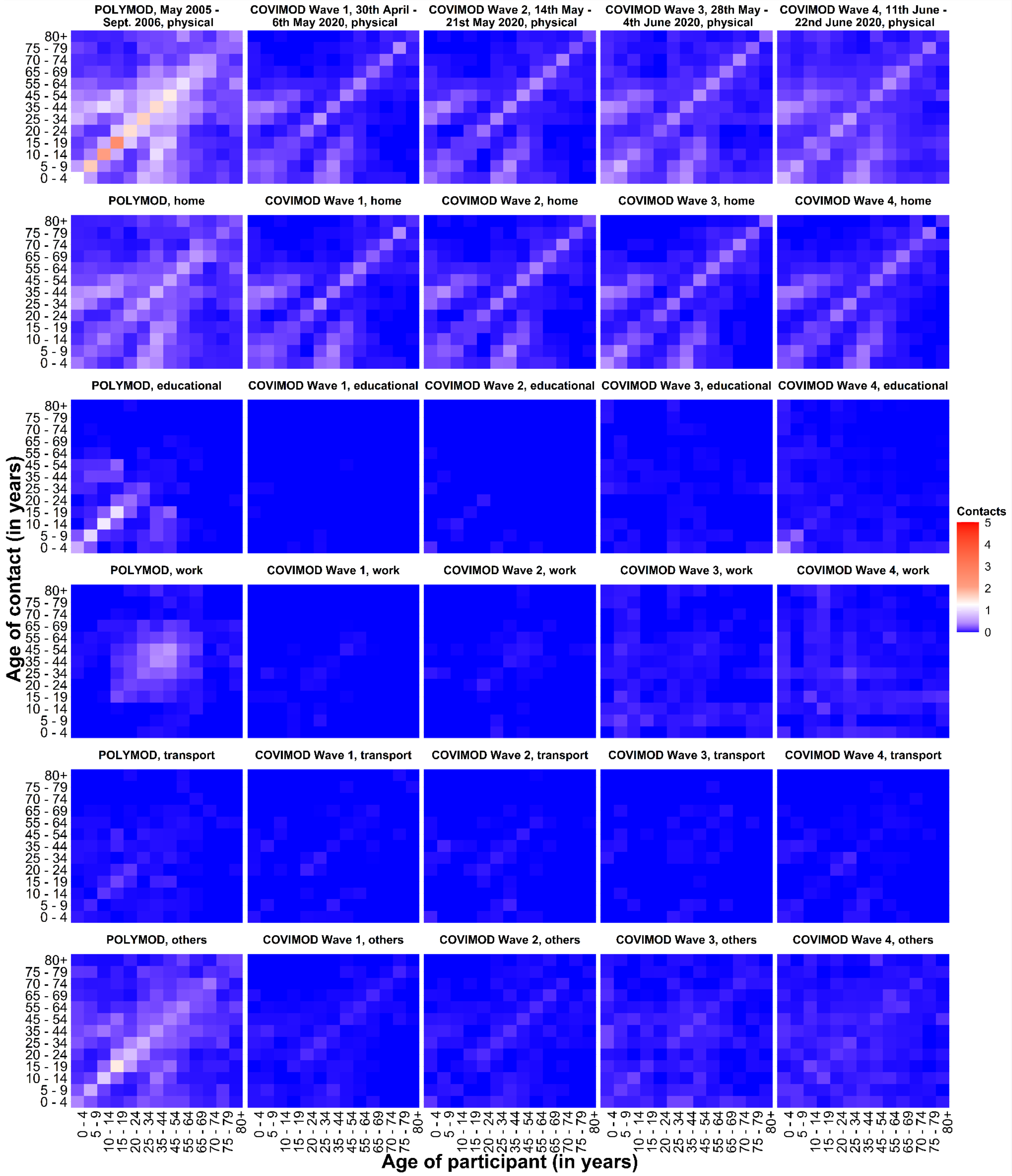

2.7. **Estimated reproduction number of SARS-CoV-2 under contact reduction measures. RKI - the percentage mean and minimum and maximum reductions in the reproduction number in the assessed time intervals, COVIMOD - the calculated percentage mean reductions in the reproduction number and the 95% confidence interval, Google and Apple mobility – the percentage mean and minimum and maximum reduction in mobility in the assessed time intervals.**

2.7.a. Figure 2.7a. Comparison of R(t) estimates obtained based on different input data. Measured effective reproduction number at the timing of COVIMOD survey Waves 1 to 4 (red), estimated reproduction number based on the reduction of social contacts at the times of COVIMOD survey Waves 1 to 4 (blue) and the reduction in mobility at the times of the COVIMOD survey Waves (yellow and green) for the unweighted analysis including group contacts. Displayed are (A) 30th April – 6th May 2020, (B) 14th May – 21st May 2020, (C) 28th May – 4th June 2020 and (D) 11th June – 22nd June 2020

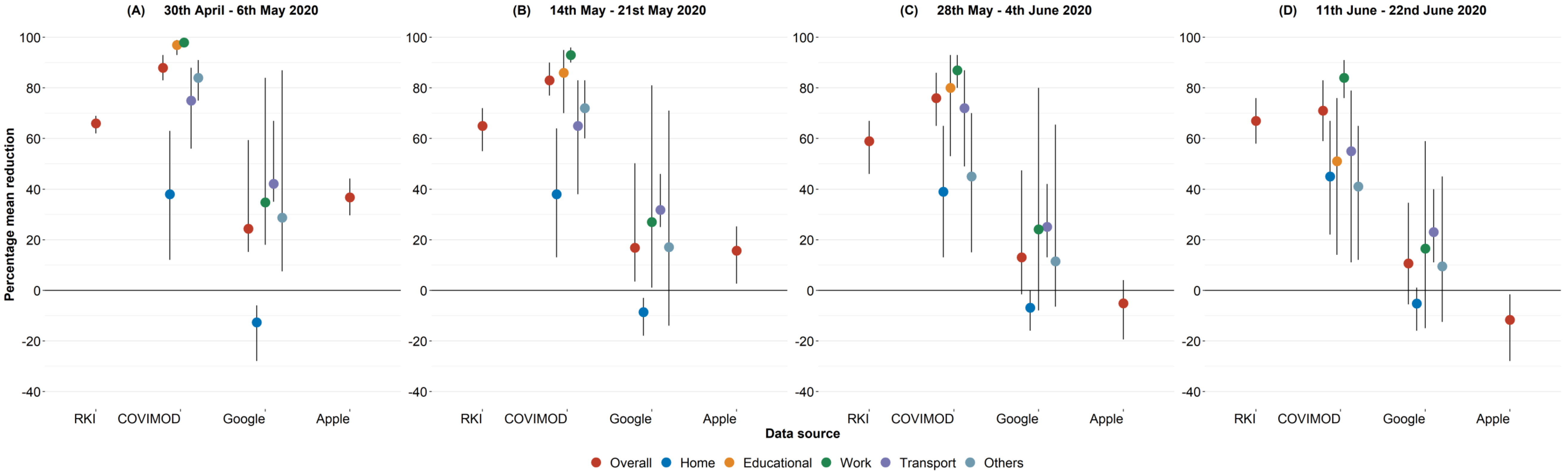


2.7.b. Figure 2.7b. Comparison of R(t) estimates obtained based on different input data. Measured effective reproduction number at the timing of COVIMOD survey Waves 1 to 4 (red), estimated reproduction number based on the reduction of social contacts at the times of COVIMOD survey Waves 1 to 4 (blue) and the reduction in mobility at the times of the COVIMOD survey Waves (yellow and green) for the unweighted analysis including group contacts.

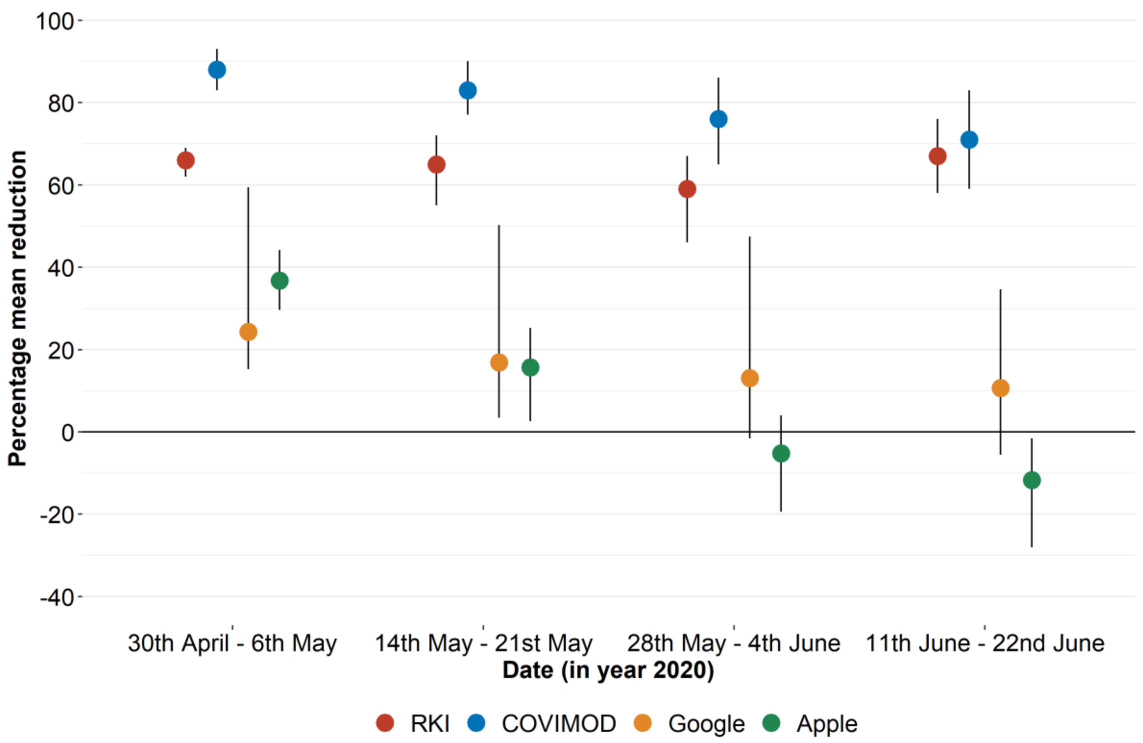

3. **WEIGHTED ANALYSIS WITHOUT GROUP CONTACTS**

3.2. **Number of social contacts.**

3.2.a. Table 3.2a. Number of recorded overall contacts per participant per day stratified by age, gender, household size, and day of the week in the COVIMOD survey Waves 1 to 4 and the POLYMOD survey for the weighted analysis without group contacts.

| | **POLYMOD** | | **COVIMOD** | | | | | | | |
|---|---|---|---|---|---|---|---|---|---|---|
| | | | **Wave1** | | **Wave 2** | | **Wave 3** | | **Wave 4** | |
| | **Mean (SD)** | **Min, Max** | **Mean (SD)** | **Min, Max** | **Mean (SD)** | **Min, Max** | **Mean (SD)** | **Min, Max** | **Mean (SD)** | **Min, Max** |
| | **7.8 (6.3)** | **1,58** | **1.9 (1.8)** | **0,16** | **3.5 (6.1)** | **0,102** | **3.4 (5.5)** | **0,101** | **3.2 (5.0)** | **0,100** |
| **Age category** | | | | | | | | | | |
| 0 - 4 | 9.4(6.2) | 2,30 | 3.1(1.7) | 0,8 | 4.6(2.5) | 0,14 | 4.8 (3.7) | 1,14 | 4.0 (3.7) | 0,33 |
| 5 - 9 | 8.5 (6.3) | 2,43 | 3.4 (1.7) | 0,8 | 3.8 (3.1) | 0,17 | 3.6 (2.8) | 0,25 | 4.7 (4.1) | 0,31 |
| 10 - 14 | 9.2 (5.8) | 1,29 | 2.9 (1.5) | 0,12 | 3.2 (2.0) | 0,9 | 3.5 (2.4) | 0,14 | 4.7 (5.5) | 0,39 |
| 15 - 19 | 9.9 (8.0) | 1,50 | 3.3 (1.9) | 0,8 | 4.3 (3.9) | 0,20 | 3.6 (3.0) | 0,14 | 4.4 (5.0) | 0,39 |
| 20 - 24 | 7.9 (5.0) | 1,35 | 1.6 (2.1) | 0,16 | 3.3 (3.1) | 0,20 | 4.9 (7.6) | 0,27 | 4.2 (7.2) | 0,84 |
| 25 - 34 | 9.1 (8.3) | 1,58 | 1.7 (1.7) | 0,13 | 3.0 (3.3) | 0,18 | 2.7 (3.4) | 0,34 | 3.4 (5.4) | 0,36 |
| 35 - 44 | 8.3 (5.4) | 1,38 | 2.0 (2.0) | 0,11 | 3.3 (3.7) | 0,33 | 3.4 (4.1) | 0,18 | 2.4 (3.3) | 0,42 |
| 45 - 54 | 8.8 (7.4) | 1,55 | 1.7 (1.6) | 0,9 | 3.7 (9.7) | 0,102 | 3.4 (4.8) | 0,36 | 3.0 (7.2) | 0,100 |
| 55 - 64 | 6.2 (3.9) | 1,25 | 1.7 (1.6) | 0,12 | 3.9 (5.5) | 0,42 | 3.8 (6.3) | 0,46 | 3.0 (4.0) | 0,39 |
| 65 - 69 | 6.4 (6.1) | 1,40 | 1.4 (1.6) | 0,11 | 4.3 (9.9) | 0,93 | 4.2 (10.7) | 0,101 | 2.9 (3.9) | 0,43 |
| 70 - 74 | 5.1 (4.4) | 1,20 | 1.2 (1.6) | 0,8 | 2.7 (3.3) | 0,25 | 2.5 (1.8) | 0,11 | 2.3 (2.2) | 0,16 |
| 75 - 79 | 4.8 (4.0) | 1,14 | 1.5 (1.2) | 0,5 | 2.6 (1.8) | 0,9 | 3.3 (4.2) | 0,32 | 2.0 (1.7) | 0,8 |
| 80 + | 3.9 (1.9) | 2,8 | 0.7 (1.0) | 0,3 | 1.2 (2.3) | 0,9 | 0.4 (0.8) | 0,4 | 0.5 (1.0) | 0,4 |
| **Sex of participants** | | | | | | | | | | |
| Female | 7.5 (5.5) | 1,45 | 1.9 (1.8) | 0,16 | 3.6 (6.6) | 0,102 | 3.7 (5.0) | 0,46 | 3.5 (5.7) | 0,100 |
| Male | 8.1 (6.9) | 1,58 | 1.9 (1.8) | 0,12 | 3.4 (5.6) | 0,93 | 3.1 (5.9) | 0,101 | 3.0 (4.2) | 0,84 |
| **Household size** | | | | | | | | | | |
| 1 | 6.3 (6.8) | 1,58 | 0.7 (1.4) | 0,12 | 1.9 (3.5) | 0,42 | 1.7 (2.8) | 0,42 | 2.3 (6.8) | 0,100 |
| 2 | 6.8 (5.8) | 1,55 | 1.3 (1.5) | 0,11 | 3.7 (8.3) | 0,101 | 3.4 (7.0) | 0,101 | 2.5 (3.9) | 0,84 |
| 3 | 7.7 (5.2) | 1,42 | 2.1 (1.4) | 0,11 | 3.5 (6.1) | 0,102 | 3.8 (5.5) | 0,46 | 3.6 (5.2) | 0,43 |
| 4 or more | 9.7 (6.6) | 1,50 | 3.3 (1.7) | 0,16 | 4.3 (3.1) | 0,32 | 4.5 (3.7) | 0,34 | 4.5 (4.4) | 0,39 |
| **Weekdays** | | | | | | | | | | |
| Monday | 7.4 (4.8) | 1,31 | 2.4 (1.7) | 0,6 | 4.1 (4.3) | 0,24 | 2.8 (3.6) | 0,42 | 3.5 (5.1) | 0,84 |
| Tuesday | 7.7 (6.3) | 1,56 | 2.5 (2.2) | 0,8 | 4.7 (9.3) | 0,102 | 4.5 (5.0) | 0,32 | 3.9 (5.2) | 0,43 |
| Wednesday | 8.0 (7.8) | 1,58 | 2.5 (2.7) | 0,13 | 3.2 (4.2) | 0,42 | 3.1 (4.2) | 0,31 | 3.2 (4.5) | 0,39 |
| Thursday | 7.1 (4.5) | 1,30 | 1.9 (1.8) | 0,16 | 3.4 (5.4) | 0,93 | 3.7 (6.8) | 0,101 | 2.5 (3.4) | 0,36 |
| Friday | 8.1 (6.5) | 1,55 | 1.9 (1.8) | 0,8 | 4.2 (4.4) | 0,32 | 3.1 (4.4) | 0,27 | 3.3 (8.3) | 0,100 |
| Saturday | 8.3 (6.4) | 1,50 | 1.7 (1.5) | 0,8 | 5.0 (14.8) | 0,101 | 2.8 (3.3) | 0,33 | 3.2 (4.2) | 0,35 |
| Sunday | 8.1 (7.6) | 1,45 | 1.8 (1.6) | 0,12 | 2.3 (2.5) | 0,21 | 2.4 (2.3) | 0,34 | 2.3 (3.8) | 0,43 |

3.2.b. Table 3.2b. Number of recorded physical contacts per participant per day stratified by age, gender, household size, and day of the week in the COVIMOD survey Waves 1 to 4 and the POLYMOD survey for the weighted analysis without group contacts.

| | **POLYMOD** | | **COVIMOD** | | | | | | | |
|---|---|---|---|---|---|---|---|---|---|---|
| | | | **Wave1** | | **Wave 2** | | **Wave 3** | | **Wave 4** | |
| | **Mean (SD)** | **Min, Max** | **Mean (SD)** | **Min, Max** | **Mean (SD)** | **Min, Max** | **Mean (SD)** | **Min, Max** | **Mean (SD)** | **Min, Max** |
| | **5.3 (4.7)** | **0,45** | **0.8 (1.2)** | **0,11** | **1.1 (3.7)** | **0,101** | **1.0 (1.6)** | **0,25** | **1.1 (1.9)** | **0,39** |
| **Age category** | | | | | | | | | | |
| 0 - 4 | 7.5 (4.9) | 0,30 | 2.7 (1.7) | 0,7 | 3.3 (2.3) | 0,8 | 3.4 (3.1) | 0,12 | 3.3 (3.0) | 0,18 |
| 5 - 9 | 6.3 (4.8) | 0,33 | 2.2 (1.7) | 0,6 | 2.2 (1.8) | 0,6 | 2.2 (1.5) | 0,5 | 3.0 (2.7) | 0,23 |
| 10 - 14 | 6.5 (4.9) | 0,25 | 1.9 (1.5) | 0,11 | 1.6 (1.7) | 0,6 | 2.1 (2.1) | 0,7 | 2.2 (1.9) | 0,8 |
| 15 - 19 | 6.5 (7.3) | 0,45 | 1.2 (1.5) | 0,8 | 1.3 (1.6) | 0,6 | 1.2 (1.4) | 0,4 | 1.5 (2.6) | 0,22 |
| 20 - 24 | 5.1 (4.1) | 0,26 | 0.6 (0.9) | 0,5 | 0.8 (1.1) | 0,3 | 0.7 (1.0) | 0,3 | 1.4 (2.9) | 0,33 |
| 25 - 34 | 5.6 (3.8) | 0,29 | 0.8 (1.0) | 0,6 | 0.8 (1.0) | 0,5 | 0.6 (0.8) | 0,4 | 0.9 (1.3) | 0,8 |
| 35 - 44 | 5.9 (4.9) | 0,36 | 0.7 (1.1) | 0,8 | 0.9 (1.1) | 0,6 | 1.4 (1.9) | 0,8 | 0.7 (1.0) | 0,6 |
| 45 - 54 | 5.9 (5.1) | 0,35 | 0.7 (1.0) | 0,4 | 1.6 (7.8) | 0,101 | 0.9 (1.2) | 0,7 | 0.9 (1.5) | 0,13 |
| 55 - 64 | 4.2 (3.4) | 0,16 | 0.6 (0.9) | 0,5 | 0.7 (1.2) | 0,10 | 0.6 (0.9) | 0,5 | 0.8 (1.6) | 0,39 |
| 65 - 69 | 4.6 (5.7) | 0,40 | 0.5 (0.8) | 0,5 | 1.0 (3.3) | 0,29 | 1.1 (2.5) | 0,25 | 0.8 (1.3) | 0,11 |
| 70 - 74 | 2.7 (3.1) | 0,13 | 0.4 (0.6) | 0,3 | 0.6 (0.8) | 0,3 | 0.6 (0.9) | 0,4 | 0.7 (0.9) | 0,4 |
| 75 - 79 | 2.7 (2.6) | 0,8 | 0.7 (0.8) | 0,3 | 0.8 (1.1) | 0,3 | 0.6 (0.6) | 0,3 | 0.6 (0.7) | 0,3 |
| 80 + | 3.0 (1.8) | 1,6 | 0.1 (0.3) | 0,1 | 0.0 (0.2) | 0,1 | 0.0 (0.1) | 0,2 | 0.0 (0.2) | 0,1 |
| **Sex of participants** | | | | | | | | | | |
| Female | 5.2 (4.7) | 0,45 | 0.9 (1.2) | 0,8 | 1.3 (4.9) | 0,101 | 1.1 (1.9) | 0,25 | 1.1 (1.8) | 0,39 |
| Male | 5.3 (4.8) | 0,44 | 0.8 (1.2) | 0,11 | 0.9 (1.8) | 0,29 | 1.0 (1.2) | 0,7 | 1.1 (1.9) | 0,33 |
| **Household size** | | | | | | | | | | |
| 1 | 3.7 (4.1) | 0,36 | 0.1 (0.4) | 0,5 | 0.2 (0.5) | 0,4 | 0.3 (0.7) | 0,7 | 0.3 (0.9) | 0,8 |
| 2 | 4.5 (4.1) | 0,45 | 0.5 (0.7) | 0,6 | 1.2 (5.9) | 0,101 | 0.8 (1.5) | 0,25 | 0.7 (1.3) | 0,33 |
| 3 | 5.4 (4.0) | 0,30 | 0.9 (1.0) | 0,8 | 1.0 (1.2) | 0,13 | 1.1 (1.4) | 0,12 | 1.4 (1.9) | 0,39 |
| 4 or more | 6.9 (5.5) | 0,44 | 1.7 (1.6) | 0,11 | 1.7 (2.0) | 0,13 | 2.0 (1.9) | 0,8 | 2.1 (2.4) | 0,23 |
| **Weekdays** | | | | | | | | | | |
| Monday | 4.9 (4.0) | 0,29 | 1.4 (1.5) | 0,6 | 1.2 (1.4) | 0,6 | 1.0 (1.1) | 0,5 | 1.3 (2.0) | 0,33 |
| Tuesday | 5.2 (4.7) | 0,40 | 1.0 (1.3) | 0,7 | 1.3 (1.7) | 0,13 | 0.9 (1.2) | 0,6 | 1.3 (2.0) | 0,39 |
| Wednesday | 5.2 (5.1) | 0,36 | 0.8 (1.5) | 0,6 | 0.9 (1.7) | 0,22 | 1.0 (1.5) | 0,6 | 1.1 (2.2) | 0,22 |
| Thursday | 5.5 (4.2) | 0,30 | 0.8 (1.1) | 0,8 | 1.0 (2.0) | 0,29 | 1.0 (1.8) | 0,25 | 0.9 (1.1) | 0,6 |
| Friday | 5.2 (4.1) | 0,23 | 0.9 (1.3) | 0,6 | 1.1 (1.8) | 0,12 | 1.0 (1.8) | 0,12 | 1.0 (1.9) | 0,23 |
| Saturday | 5.8 (5.2) | 0,44 | 1.1 (1.4) | 0,8 | 3.1 (15.0) | 0,101 | 1.2 (1.3) | 0,5 | 1.3 (2.0) | 0,13 |
| Sunday | 5.8 (6.3) | 0,45 | 0.9 (1.2) | 0,11 | 0.9 (1.2) | 0,5 | 1.1 (1.3) | 0,5 | 1.0 (1.4) | 0,8 |

3.2.c. Table 3.2c. Number of recorded overall contacts per different settings in the COVIMOD survey Waves 1 to 4 and the POLYMOD survey for the weighted analysis without group contacts.

**Note:** *The displayed educational contacts are based on the group of participants who attended an educational facility (kindergarten, school, university) and work contacts are based on the group of participants who reported to work full-/part-time.*

| | **POLYMOD** | | **COVIMOD** | | | | | | | |
|---|---|---|---|---|---|---|---|---|---|---|
| | | | **Wave1** | | **Wave2** | | **Wave3** | | **Wave4** | |
| | **Mean (SD)** | **Min, Max** | **Mean (SD)** | **Min, Max** | **Mean (SD)** | **Min, Max** | **Mean (SD)** | **Min, Max** | **Mean (SD)** | **Min, Max** |
| Overall | 7.8 (6.3) | 1,58 | 1.9 (1.8) | 0,16 | 3.5 (6.1) | 0,102 | 3.4 (5.5) | 0,101 | 3.2 (5.0) | 0,100 |
| Home | 2.9 (2.3) | 0,26 | 1.5 (1.5) | 0,12 | 1.6 (1.6) | 0, 9 | 1.5 (1.5) | 0, 13 | 1.5 (1.5) | 0, 23 |
| Educational | 2.6 (4.3) | 0,42 | 0.0 (0.2) | 0, 3 | 0.2 (1.0) | 0, 10 | 0.1 (0.9) | 0, 17 | 0.6 (2.3) | 0, 22 |
| Work | 2.8 (5.3) | 0,56 | 0.3 (1.0) | 0,11 | 2.0 (7.3) | 0,100 | 2.2 (6.8) | 0, 97 | 1.6 (5.8) | 0,100 |
| Transport | 0.3 (0.9) | 0, 8 | 0.0 (0.2) | 0, 3 | 0.1 (0.4) | 0, 4 | 0.0 (0.3) | 0, 8 | 0.1 (0.5) | 0, 12 |
| Others | 2.9 (3.7) | 0,45 | 0.4 (0.9) | 0,10 | 1.0 (2.2) | 0, 33 | 1.1 (2.6) | 0, 35 | 1.0 (2.3) | 0, 38 |

3.2.d. Table 3.2d. Number of recorded physical contacts per different settings in the COVIMOD survey Waves 1 to 4 and the POLYMOD survey for the weighted analysis without group contacts.

**Note:** *The displayed educational contacts are based on the group of participants who attended an educational facility (kindergarten, school, university) and work contacts are based on the group of participants who reported to work full-/part-time.*

| | **POLYMOD** | | **COVIMOD** | | | | | | | |
|---|---|---|---|---|---|---|---|---|---|---|
| | | | **Wave1** | | **Wave2** | | **Wave3** | | **Wave4** | |
| | **Mean (SD)** | **Min, Max** | **Mean (SD)** | **Min, Max** | **Mean (SD)** | **Min, Max** | **Mean (SD)** | **Min, Max** | **Mean (SD)** | **Min, Max** |
| Overall | 5.3 (4.7) | 0,45 | 0.8 (1.2) | 0,11 | 1.1 (3.7) | 0,101 | 1.0 (1.6) | 0,25 | 1.1 (1.9) | 0,39 |
| Home | 2.4 (2.2) | 0,26 | 0.8 (1.2) | 0,11 | 0.8 (1.2) | 0, 8 | 0.9 (1.3) | 0,12 | 0.9 (1.2) | 0, 8 |
| Educational | 1.4 (3.5) | 0,42 | 0.0 (0.0) | 0, 1 | 0.1 (0.4) | 0, 4 | 0.0 (0.2) | 0, 2 | 0.2 (1.3) | 0,21 |
| Work | 1.6 (3.0) | 0,30 | 0.0 (0.1) | 0, 2 | 0.5 (5.2) | 0,100 | 0.2 (1.4) | 0,25 | 0.2 (1.5) | 0,31 |
| Transport | 0.2 (0.7) | 0, 6 | 0.0 (0.2) | 0, 3 | 0.1 (0.3) | 0, 3 | 0.0 (0.2) | 0, 3 | 0.1 (0.4) | 0, 7 |
| Others | 1.9 (3.0) | 0,40 | 0.1 (0.4) | 0, 5 | 0.2 (0.6) | 0, 11 | 0.2 (0.8) | 0, 7 | 0.3 (1.0) | 0,31 |

### 3.3. Reproduction number estimates of SARS-CoV-2 under contact reduction measures.

3.3.a. Table 3.3a. Effective reproduction number at the timing of COVIMOD survey Waves 1 to 4, estimated effective reproduction number based on the reduction of social contacts at the times of COVIMOD survey Waves 1 to 4 assuming values of the basic reproduction number of Norm (2.6, SD=0.54) and the reduction in mobility at the times of the COVIMOD survey Waves, with 10000 bootstrapped samples in various settings and in different time frames for the weighted analysis without group contacts.

**Note:** *RKI - the percentage mean and minimum and maximum reductions in the reproduction number in the assessed time intervals, COVIMOD - the calculated percentage mean reductions in the reproduction number and the 95% confidence interval, Google and Apple mobility – the percentage mean and minimum and maximum reduction in mobility in the assessed time intervals.*

| | **COVIMOD** | | **RKI** | | **Google** | | **Apple** | |
|---|---|---|---|---|---|---|---|---|
| | **Mean reproduction number (Min, Max)** | **Percent Mean Reduction Number (Min, Max)** | **Mean reproduction number (Min, Max)** | **Percent Mean Reduction Number (Min, Max)** | **Mean reproduction number (Min, Max)** | **Percent Mean Reduction Number (Min, Max)** | **Mean reproduction number (Min, Max)** | **Percent Mean Reduction Number (Min, Max)** |
| **Overall** | | | | | | | | |
| 30th April - 6th May 2020 | 0.57 (0.32, 0.86) | -78.00 (-87.00, -66.00) | 0.88 (0.79, 0.97) | -66.00 (-69.00, -62.00) | - | -24.31 (-59.40, -15.20) | - | -36.77 (-44.17, -29.61) |
| 14th May - 21st May 2020 | 0.81 (0.46, 1.24) | -68.00 (-82.00, -52.00) | 0.90 (0.72, 1.15) | -65.00 (-72.00, -55.00) | - | -16.85 (-50.20, -3.40) | - | -15.64 (-25.26, -2.58) |
| 28th May - 4th June 2020 | 0.87 (0.46, 1.42) | -66.00 (-82.00, -45.00) | 1.06 (0.84, 1.39) | -59.00 (-67.00, -46.00) | - | -13.05 (-47.40, 1.60) | - | 5.18 (-4.00, 19.43) |
| 11th June - 22nd June 2020 | 0.81 (0.45, 1.23) | -68.00 (-82.00, -52.00) | 0.84 (0.60, 1.07) | -67.00 (-76.00, -58.00) | - | -10.63 (-34.60, 5.60) | - | 11.66 (1.62, 28.00) |
| **Home** | | | | | | | | |
| 30th April - 6th May 2020 | 1.40 (0.83, 2.03) | -46.00 (-68.00, -21.00) | - | - | - | 12.71 (6.00, 28.00) | - | - |
| 14th May - 21st May 2020 | 1.35 (0.79, 1.97) | -48.00 (-69.00, -24.00) | - | - | - | 8.62 (3.00, 18.00) | - | - |
| 28th May - 4th June 2020 | 1.43 (0.82, 2.12) | -45.00 (-68.00, -18.00) | - | - | - | 6.88 (0.00, 16.00) | - | - |
| 11th June - 22nd June 2020 | 1.26 (0.74, 1.82) | -51.00 (-71.00, -30.00) | - | - | - | 5.25 (-1.00, 16.00) | - | - |
| **Educational** | | | | | | | | |
| 30th April - 6th May 2020 | 0.04 (0.01, 0.11) | -98.00 (-99.00, -95.00) | - | - | - | - | - | - |
| 14th May - 21st May 2020 | 0.20 (0.04, 0.63) | -92.00 (-98.00, -75.00) | - | - | - | - | - | - |
| 28th May - 4th June 2020 | 0.12 (0.03, 0.35) | -95.00 (-98.00, -86.00) | - | - | - | - | - | - |
| 11th June - 22nd June 2020 | 0.52 (0.14, 1.37) | -80.00 (-94.00, -47.00) | - | - | - | - | - | - |
| **Work** | | | | | | | | |
| 30th April - 6th May 2020 | 0.24 (0.09, 0.53) | -90.00 (-96.00, -79.00) | - | - | - | -34.71 (-84.00, -18.00) | - | - |
| 14th May - 21st May 2020 | 0.82 (0.37, 1.59) | -68.00 (-85.00, -38.00) | - | - | - | -27.00 (-81.00, -1.00) | - | - |
| 28th May - 4th June 2020 | 1.09 (0.35, 2.47) | -58.00 (-86.00, -4.00) | - | - | - | -24.12 (-80.00, 8.00) | - | - |
| 11th June - 22nd June 2020 | 0.81 (0.36, 1.60) | -68.00 (-86.00, -38.00) | - | - | - | -16.50 (-59.00, 15.00) | - | - |
| **Transport** | | | | | | | | |
| 30th April - 6th May 2020 | 0.64 (0.12, 2.20) | -75.00 (-95.00, -15.00) | - | - | - | -42.14 (-67.00, -35.00) | - | - |
| 14th May - 21st May 2020 | 0.90 (0.15, 3.16) | -65.00 (-94.00, 22.00) | - | - | - | -31.75 (-46.00, -25.00) | - | - |
| 28th May - 4th June 2020 | 0.53 (0.11, 1.73) | -79.00 (-95.00, -33.00) | - | - | - | -25.12 (-42.00, -13.00) | - | - |
| 11th June - 22nd June 2020 | 1.24 (0.17, 4.85) | -52.00 (-93.00, 87.00) | - | - | - | -23.00 (-40.00, -11.00) | - | - |
| **Others** | | | | | | | | |
| 30th April - 6th May 2020 | 0.29 (0.14, 0.53) | -88.00 (-94.00, -79.00) | - | - | - | -28.71 (-87.00, -7.50) | - | - |
| 14th May - 21st May 2020 | 0.57 (0.27, 1.02) | -78.00 (-89.00, -60.00) | - | - | - | -17.06 (-71.00, 14.00) | - | - |
| 28th May - 4th June 2020 | 0.72 (0.30, 1.45) | -72.00 (-88.00, -44.00) | - | - | - | -11.44 (-65.50, 6.50) | - | - |
| 11th June - 22nd June 2020 | 0.58 (0.28, 1.05) | -77.00 (-89.00, -59.00) | - | - | - | -9.46 (-45.00, 12.50) | - | - |

3.4. **Figure 3.4. Boxplots of the number of overall contacts during the POLYMOD and COVIMOD survey waves 1 to 4 in various settings. The boundaries of the boxes closest to zero indicates the 25th percentiles, the lines within the boxes marks the medians, the red dots within the boxes marks the means, and the boundary of the boxes farthest from zero indicates the 75th percentiles. Whiskers above and below the boxes indicate the 10th and 90th percentiles; black dots represent outliers. Participants with no contacts are displayed as 0 on the log-scale of the y-axis.**

3.4.a. Figure 3.4a. Displayed are (A) the overall number of contacts, (B) home contacts, (C) work contacts, (D) educational contacts, (E) public transport contacts and (F) other contacts for the weighted analysis without group contacts in overall contacts.

**Note:** *The displayed educational contacts are based on the group of participants who attended an educational facility (kindergarten, school, university) and work contacts are based on the group of participants who reported to work full-/part-time.*

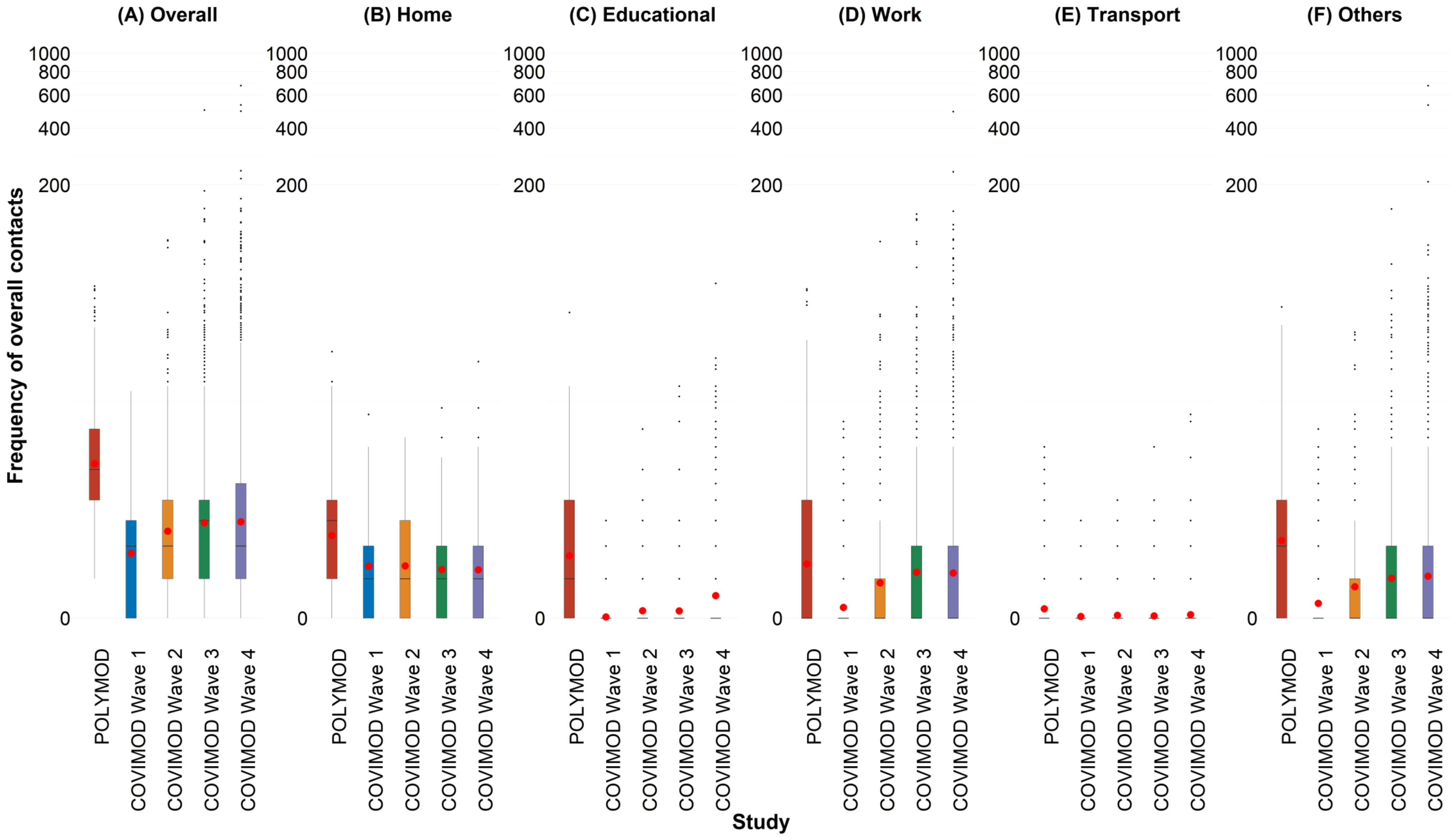

3.4.b. Figure 3.4b. Displayed are (A) 0 - 7 years, (B) 8 – 15 years, (C) 16 – 23 years, (D) 24 – 38 years, (E) 39 – 60 years and (F) 61 years or more for the weighted analysis without group contacts in overall contacts.

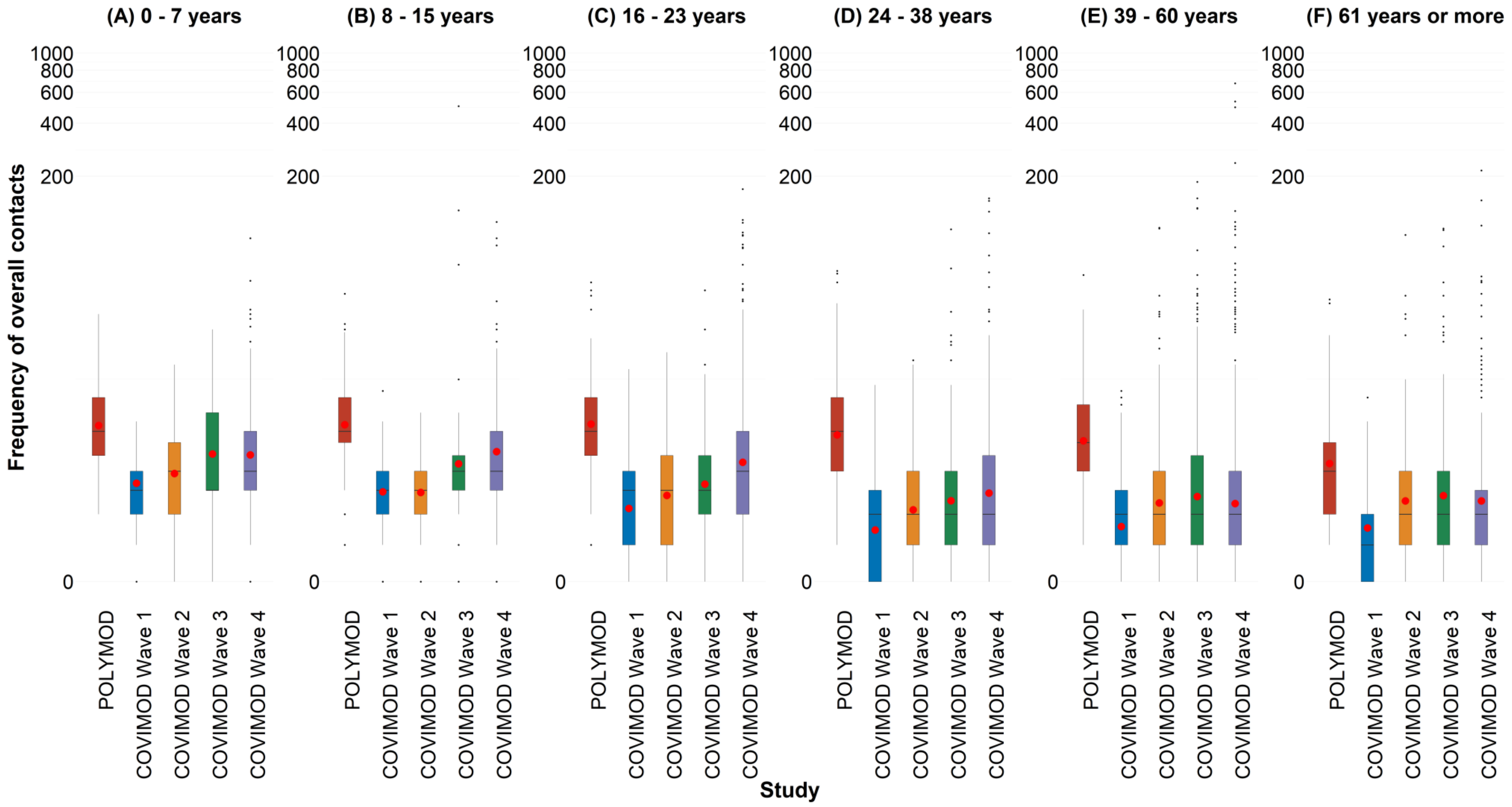

3.4.c. Figure 3.4c. Displayed are (A) Household size of 1, (B) Household size of 2, (C) Household size of 3 and (D) Household size of 4 or more for the weighted analysis without group contacts in overall contacts.

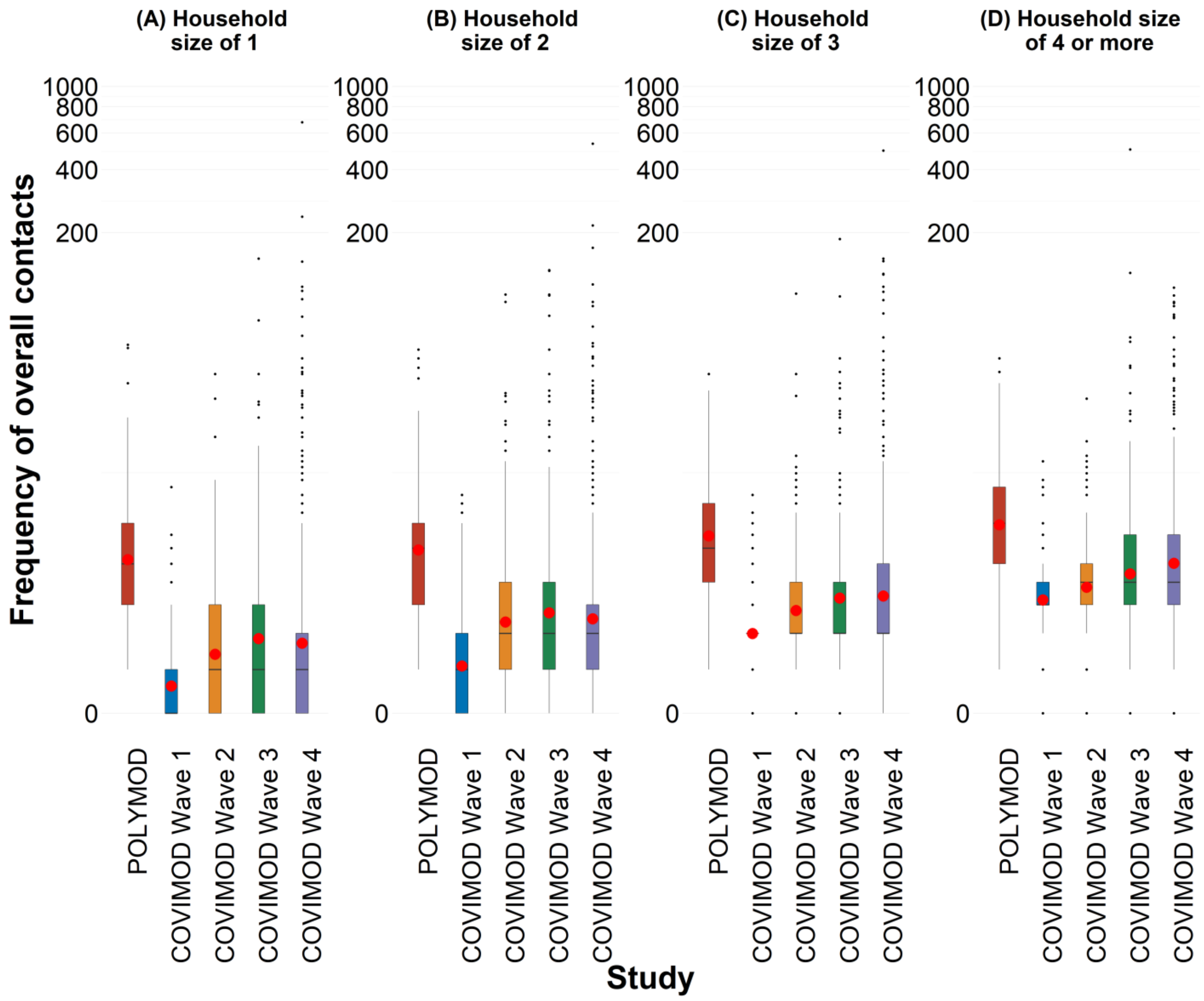

3.4.d. Figure 3.4d. Displayed are (A) Female and (B) Male for the weighted analysis without group contacts in overall contacts.

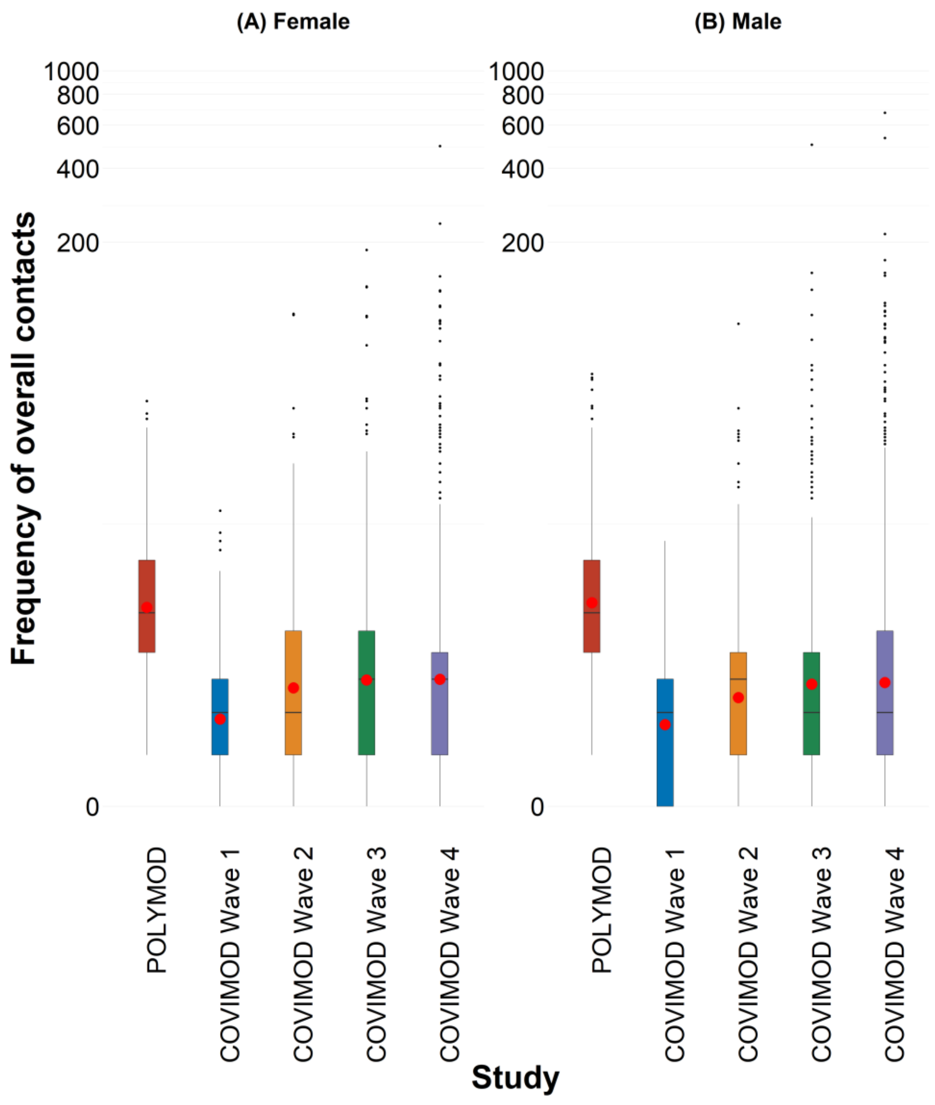

3.4.e. Figure 3.4e. Displayed are (A) Monday, (B) Tuesday, (C) Wednesday, (D) Thursday, (E) Friday, (F) Saturday and (G) Sunday for the weighted analysis without group contacts in overall contacts.

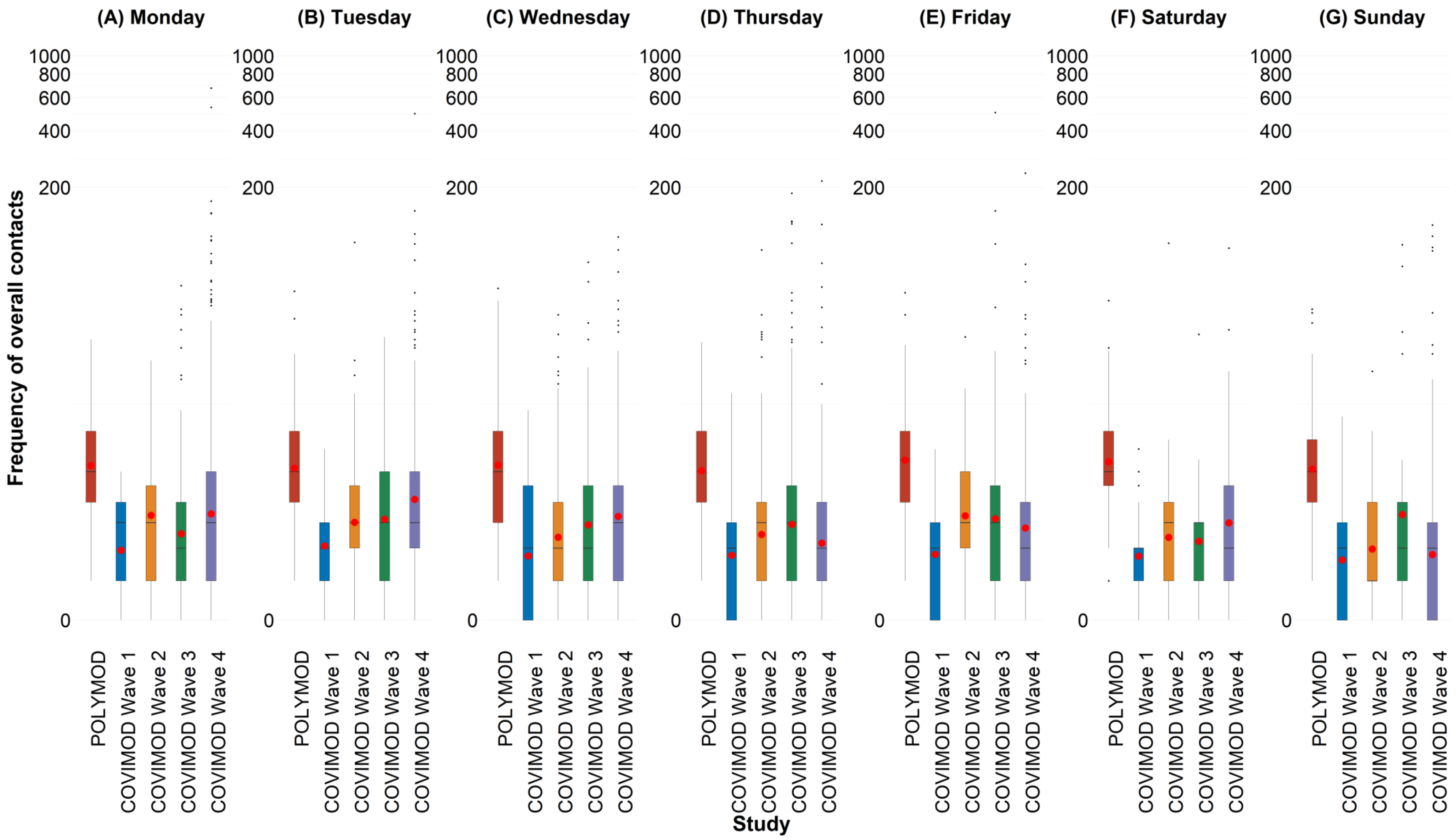

3.5. **Figure 3.5. Boxplots of the number of physical contacts during the POLYMOD and COVIMOD survey waves 1 to 4 in various settings. The boundaries of the boxes closest to zero indicates the 25th percentiles, the lines within the boxes marks the medians, the red dots within the boxes marks the means, and the boundary of the boxes farthest from zero indicates the 75th percentiles. Whiskers above and below the boxes indicate the 10th and 90th percentiles; black dots represent outliers. Participants with no contacts are displayed as 0 on the log-scale of the y-axis.**

3.5.a. Figure 3.5a. Displayed are (A) the overall number of contacts, (B) home contacts, (C) work contacts, (D) educational contacts, (E) public transport contacts and (F) other contacts for the weighted analysis without group contacts in physical contacts.

**Note:** *The displayed educational contacts are based on the group of participants who attended an educational facility (kindergarten, school, university) and work contacts are based on the group of participants who reported to work full-/part-time.*

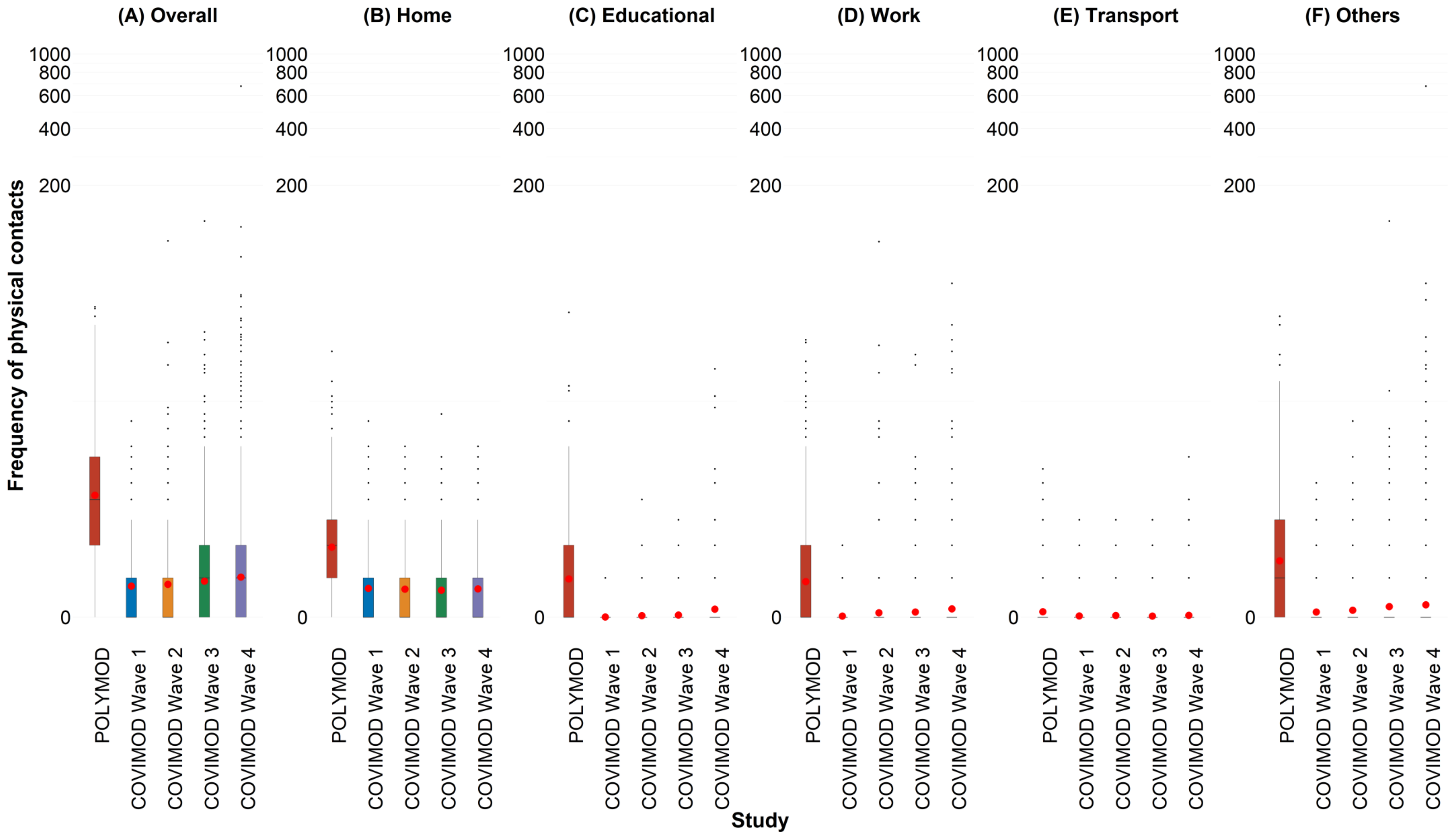

3.5.b. Figure 3.5b. Displayed are (A) 0 - 7 years, (B) 8 – 15 years, (C) 16 – 23 years, (D) 24 – 38 years, (E) 39 – 60 years and (F) 61 years or more for the weighted analysis without group contacts in physical contacts.

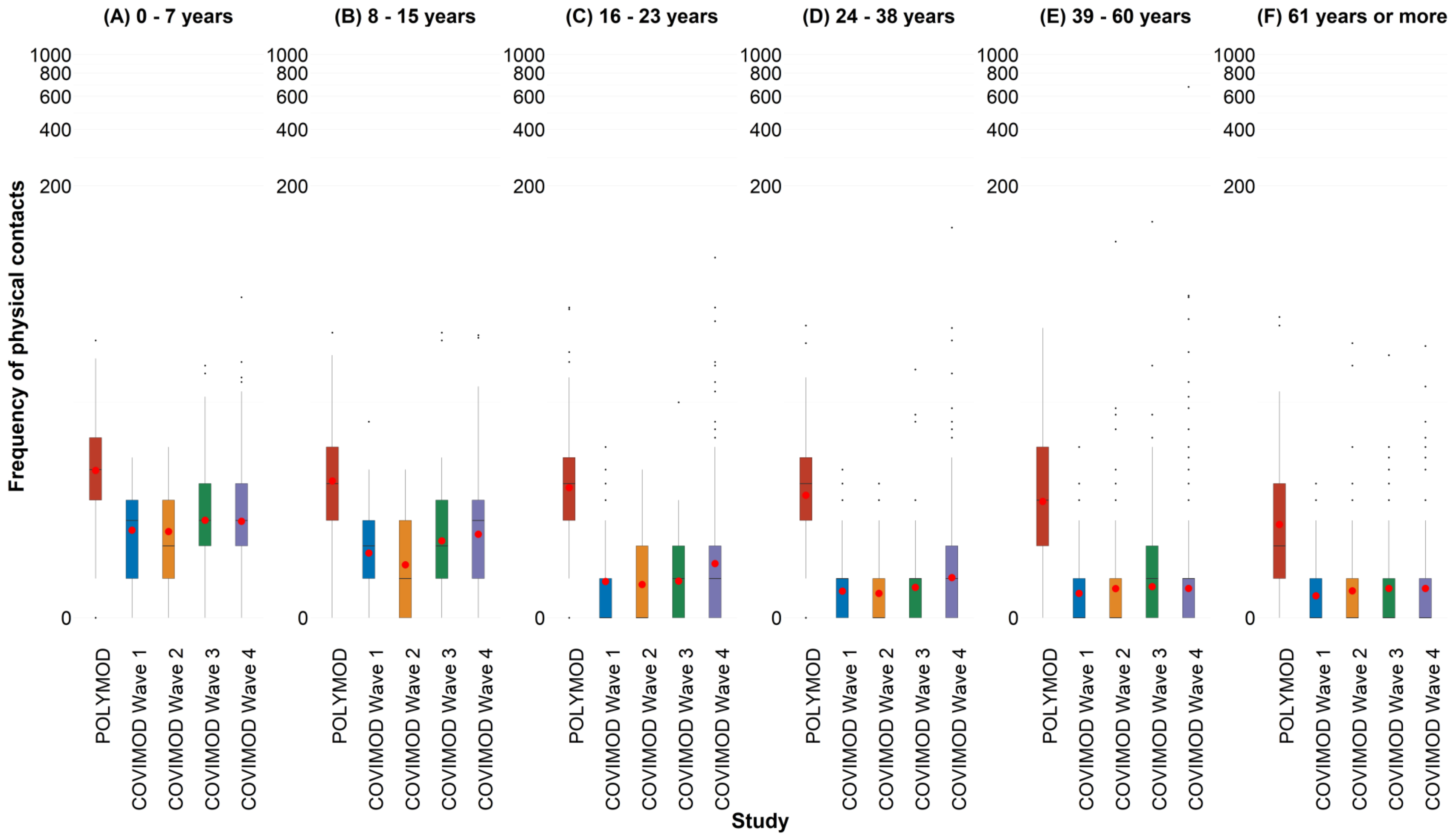

3.5.c. Figure 3.5c. Displayed are (A) Household size of 1, (B) Household size of 2, (C) Household size of 3 and (D) Household size of 4 or more for the weighted analysis without group contacts in physical contacts.

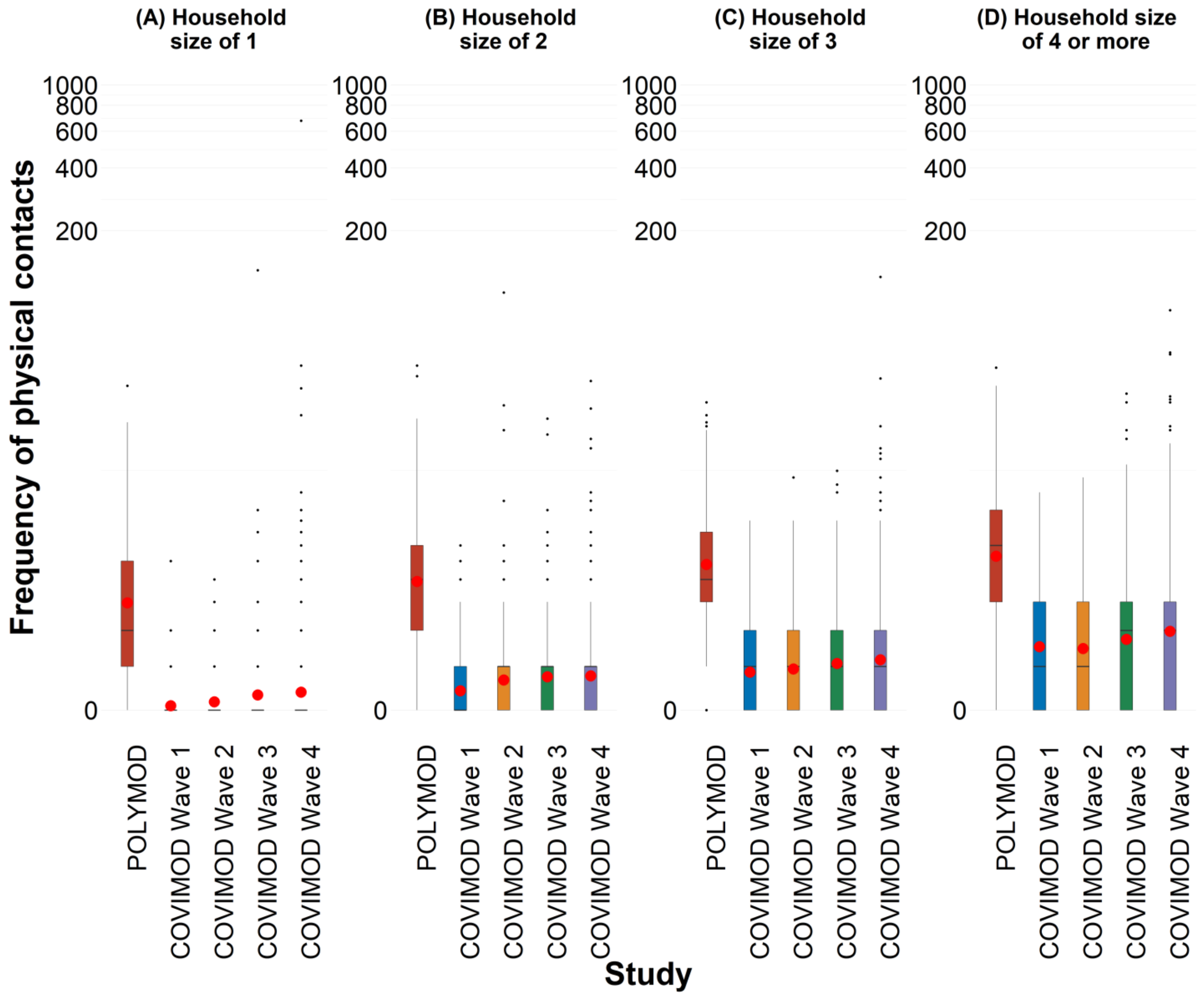

3.5.d. Figure 3.5d. Displayed are (A) Female and (B) Male for the weighted analysis without group contacts in physical contacts.

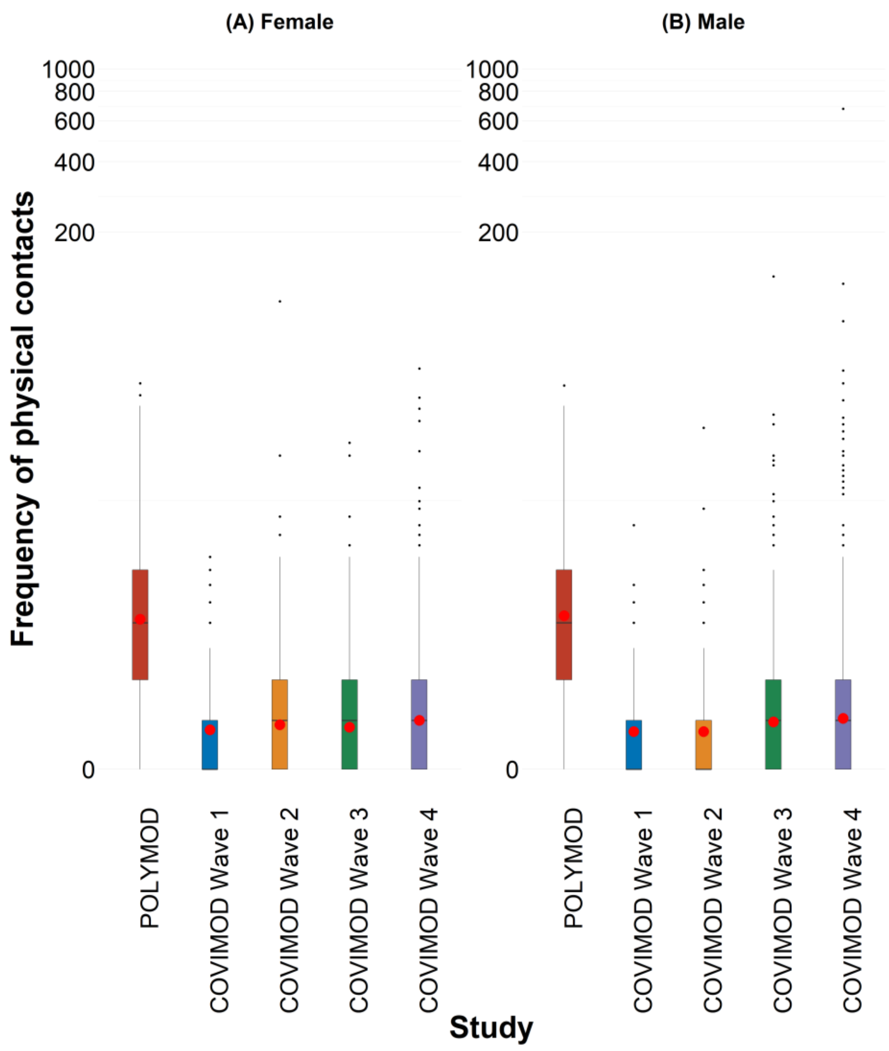

3.5.e. Figure 3.5e. Displayed are (A) Monday, (B) Tuesday, (C) Wednesday, (D) Thursday, (E) Friday, (F) Saturday and (G) Sunday for the weighted analysis without group contacts in physical contacts.

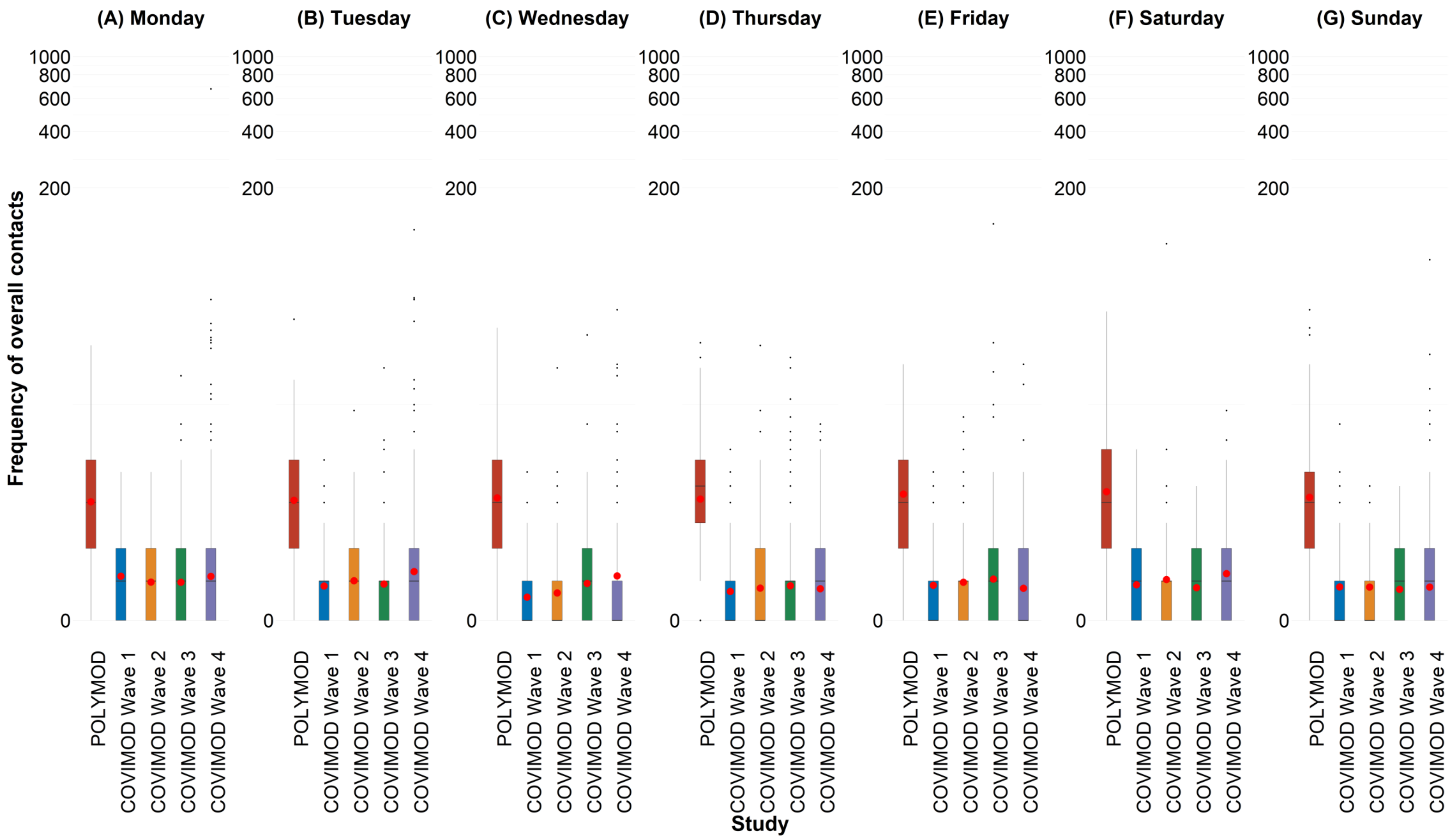

### 3.6. Social contact patterns.

3.6.a. Figure 3.6a. Social contact matrices with the mean total number of reported daily social contacts by participants in different age groups with individuals in other age groups in POLYMOD and COVIMOD survey waves 1 to 4 in various settings. Displayed are (A) the overall number of contacts, (B) home contacts, (C) educational contacts, (D) work contacts, (E) public transport contacts and (F) other contacts for the weighted analysis without group contacts.

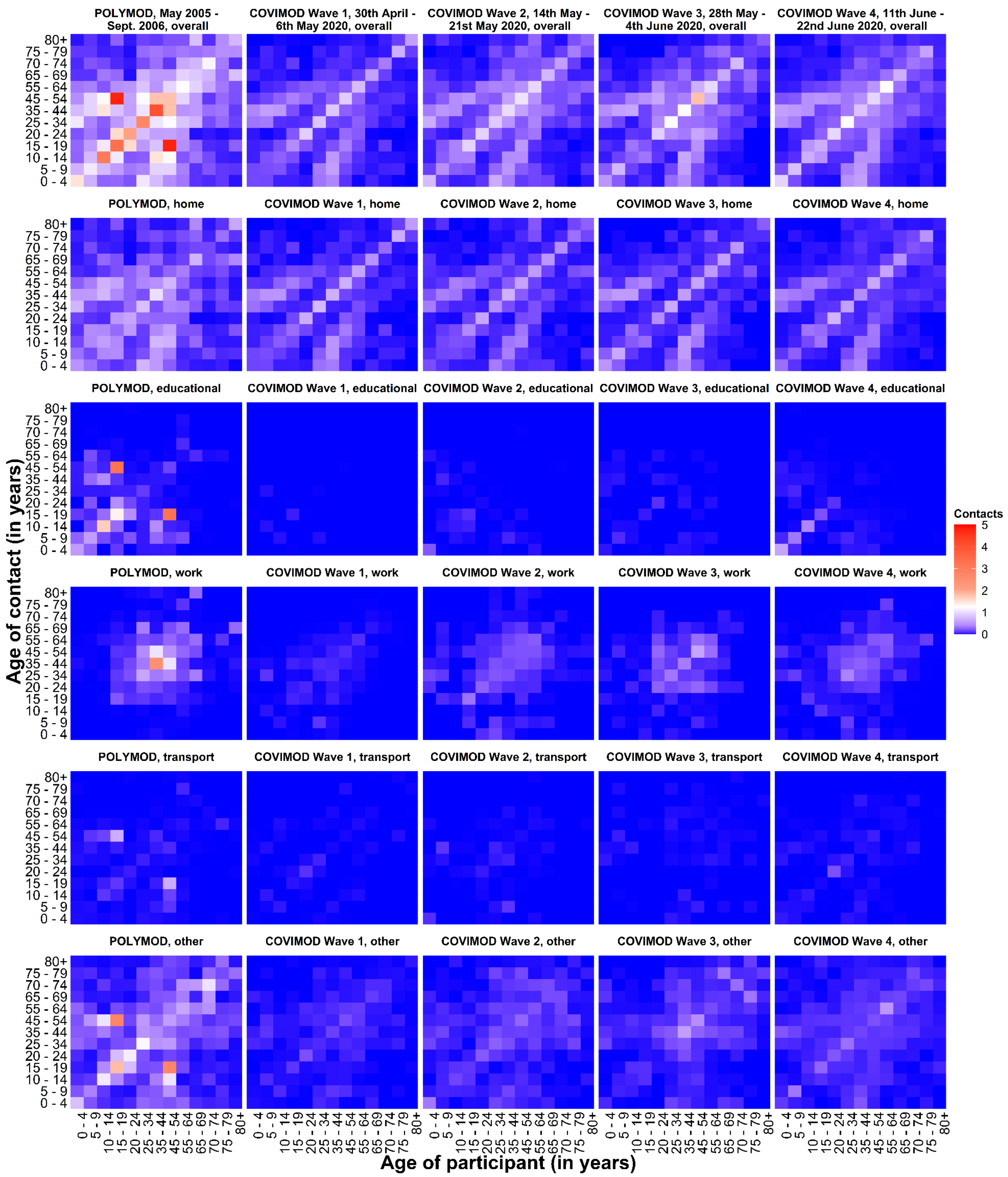

3.6.b. Figure 3.6b. Social contact matrices with the mean physical number of reported daily social contacts by participants in different age groups with individuals in other age groups in POLYMOD and COVIMOD survey waves 1 to 4 in various settings. Displayed are (A) the overall number of contacts, (B) home contacts, (C) educational contacts, (D) work contacts, (E) public transport contacts and (F) other contacts for the weighted analysis without group contacts.

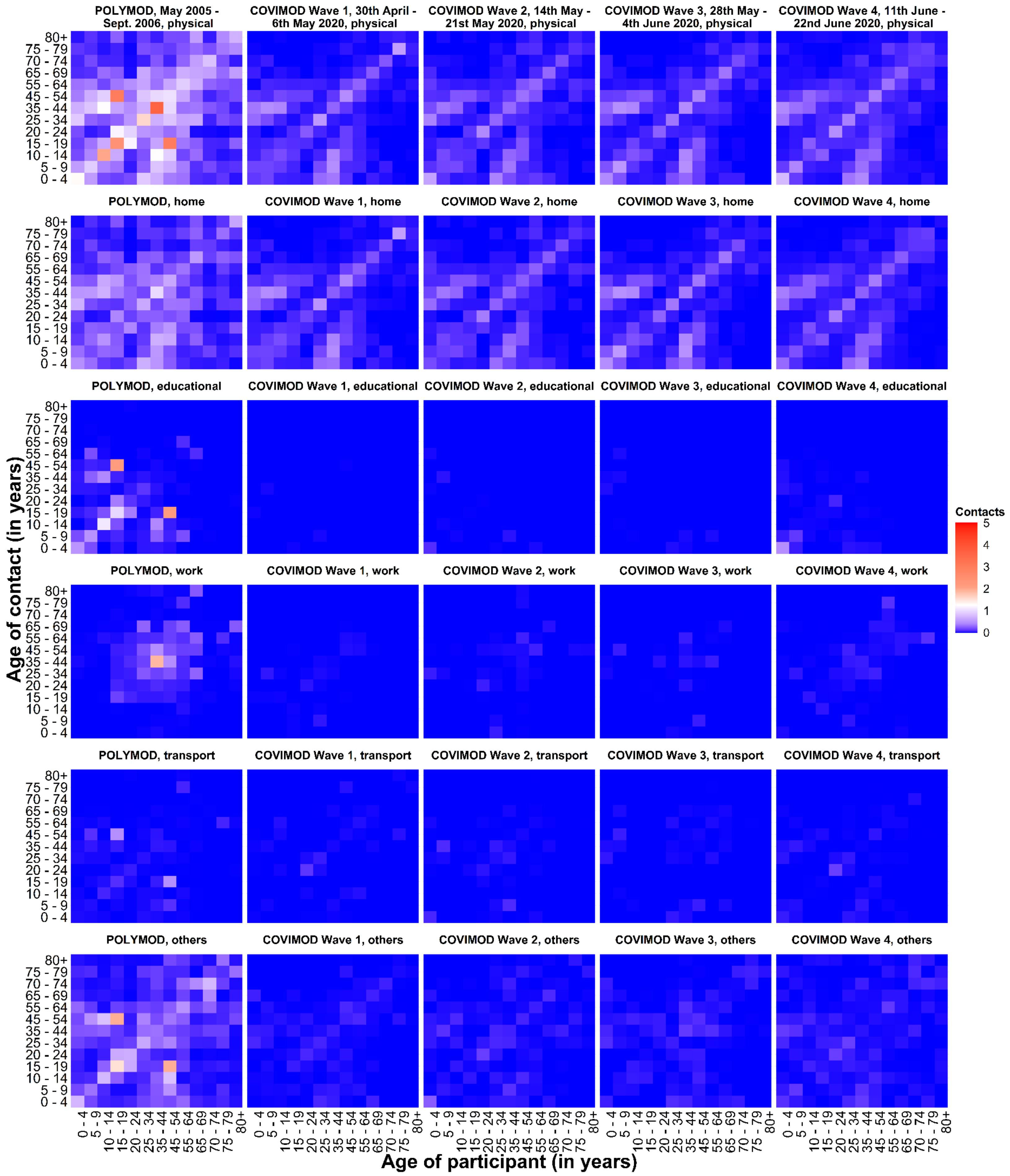

3.7. **Estimated reproduction number of SARS-CoV-2 under contact reduction measures. RKI - the percentage mean and minimum and maximum reductions in the reproduction number in the assessed time intervals, COVIMOD - the calculated percentage mean reductions in the reproduction number and the 95% confidence interval, Google and Apple mobility – the percentage mean and minimum and maximum reduction in mobility in the assessed time intervals.**

3.7.a. Figure 3.7a. Comparison of R(t) estimates obtained based on different input data. Measured effective reproduction number at the timing of COVIMOD survey Waves 1 to 4 (red), estimated reproduction number based on the reduction of social contacts at the times of COVIMOD survey Waves 1 to 4 (blue) and the reduction in mobility at the times of the COVIMOD survey Waves (yellow and green) for the weighted analysis without group contacts. Displayed are (A) 30th April – 6th May 2020, (B) 14th May – 21st May 2020, (C) 28th May – 4th June 2020 and (D) 11th June – 22nd June 2020

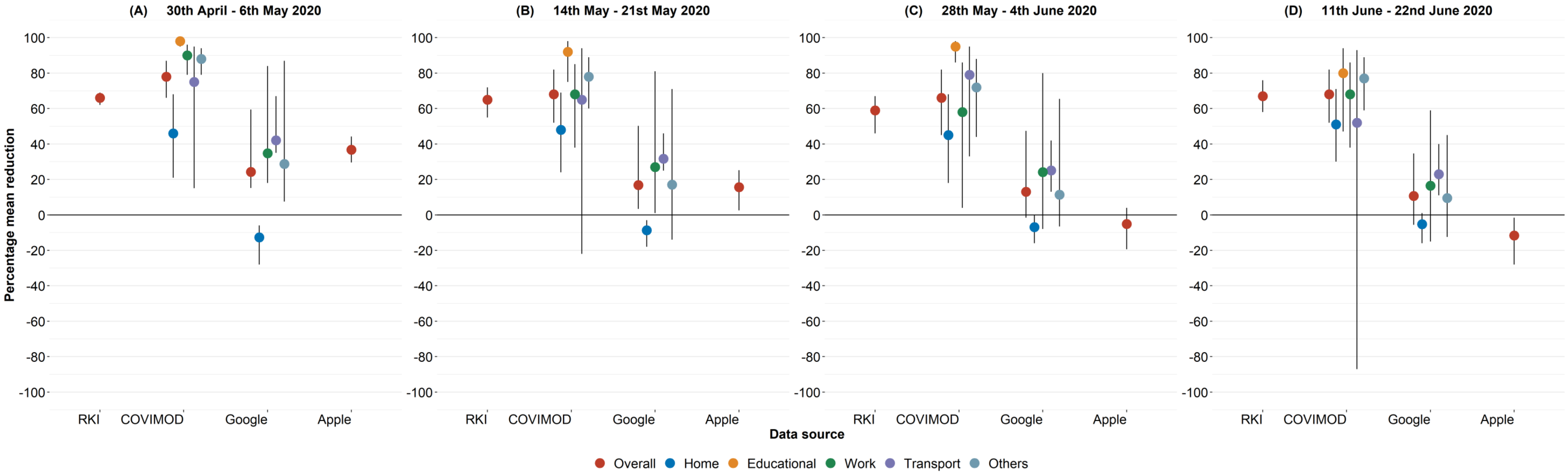


3.7.b. Figure 3.7b. Comparison of R(t) estimates obtained based on different input data. Measured effective reproduction number at the timing of COVIMOD survey Waves 1 to 4 (red), estimated reproduction number based on the reduction of social contacts at the times of COVIMOD survey Waves 1 to 4 (blue) and the reduction in mobility at the times of the COVIMOD survey Waves (yellow and green) for the weighted analysis without group contacts.

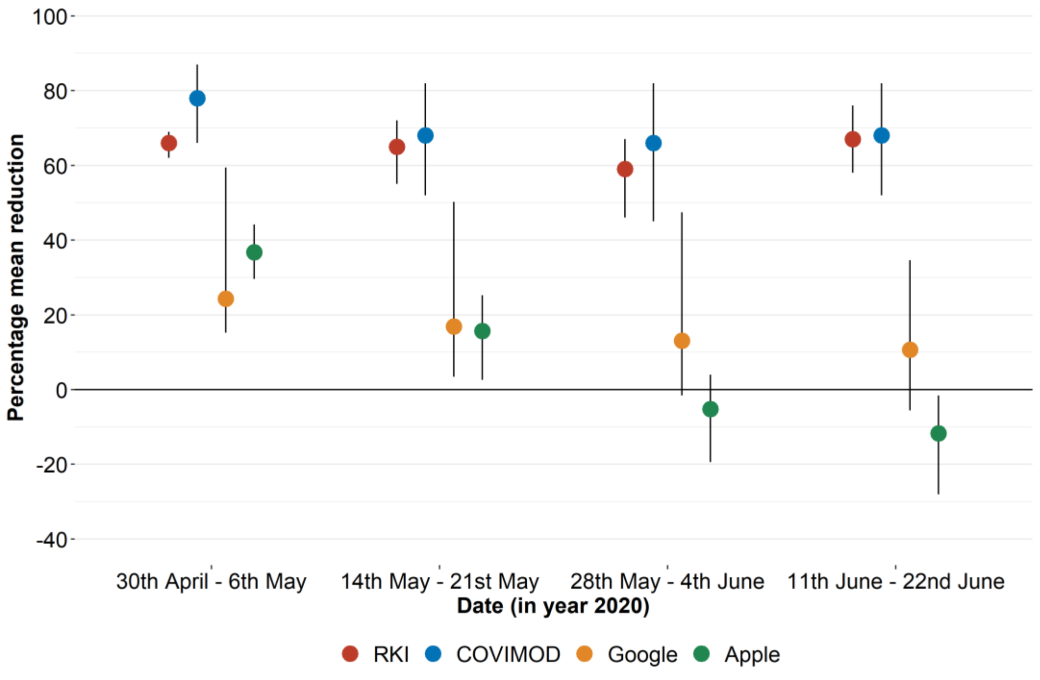

### 4. UNWEIGHTED ANALYSIS INCLUDING GROUP CONTACTS

#### 4.2. Number of social contacts.

4.2.a. Table 4.2a. Number of recorded overall contacts per participant per day stratified by age, gender, household size, and day of the week in the COVIMOD survey Waves 1 to 4 and the POLYMOD survey for the unweighted analysis without group contacts.

| | POLYMOD | | | COVIMOD | | | | | | | | | | | |
|---|---|---|---|---|---|---|---|---|---|---|---|---|---|---|---|
| | | | | Wave1 | | | Wave 2 | | | Wave 3 | | | Wave 4 | | |
| | N | Mean (SD) | Min, Max | N | Mean (SD) | Min, Max | N | Mean (SD) | Min, Max | N | Mean (SD) | Min, Max | N | Mean (SD) | Min, Max |
| | **1341** | **7.9 (6.3)** | **1,58** | **1560** | **2.1 (1.9)** | **0,16** | **1356** | **3.6 (6.1)** | **0,102** | **1081** | **3.4 (5.4)** | **0,101** | **1890** | **3.3 (5.3)** | **0,100** |
| **Age category** | | | | | | | | | | | | | | | |
| 0 - 4 | 89 | 9.1 (6.1) | 2,30 | 46 | 3.8 (1.9) | 0,8 | 36 | 4.7 (2.7) | 0,14 | 21 | 5.0 (3.7) | 1,14 | 56 | 4.7 (5.3) | 0,33 |
| 5 - 9 | 92 | 8.4 (5.7) | 2,43 | 48 | 3.5 (1.7) | 0,8 | 41 | 4.4 (3.2) | 0,17 | 30 | 4.8 (4.6) | 0,25 | 62 | 5.5 (5.1) | 0,31 |
| 10 - 14 | 110 | 9.0 (5.7) | 1,29 | 73 | 3.2 (1.8) | 0,12 | 63 | 3.3 (2.0) | 0,9 | 45 | 3.6 (2.6) | 0,14 | 87 | 5.1 (5.5) | 0,39 |
| 15 - 19 | 121 | 10.0 (7.6) | 1,50 | 95 | 3.3 (2.0) | 0,8 | 66 | 4.1 (3.7) | 0,20 | 45 | 3.7 (3.1) | 0,14 | 108 | 4.6 (5.4) | 0,39 |
| 20 - 24 | 117 | 7.9 (4.9) | 1,35 | 83 | 2.0 (2.6) | 0,16 | 60 | 3.0 (3.3) | 0,20 | 28 | 4.1 (6.0) | 0,27 | 109 | 4.6 (8.9) | 0,84 |
| 25 - 34 | 132 | 9.0 (8.5) | 1,58 | 173 | 1.7 (1.8) | 0,13 | 148 | 2.8 (3.2) | 0,18 | 96 | 2.7 (4.3) | 0,34 | 219 | 3.3 (5.3) | 0,36 |
| 35 - 44 | 156 | 8.1 (5.4) | 1,38 | 137 | 1.9 (2.2) | 0,11 | 124 | 3.0 (3.7) | 0,33 | 91 | 2.5 (3.0) | 0,18 | 164 | 2.4 (4.0) | 0,42 |
| 45 - 54 | 184 | 7.9 (7.1) | 1,55 | 235 | 1.8 (1.7) | 0,9 | 209 | 4.0 (10.3) | 0,102 | 174 | 2.9 (4.6) | 0,36 | 275 | 2.9 (7.3) | 0,100 |
| 55 - 64 | 160 | 6.0 (3.8) | 1,25 | 265 | 1.9 (1.7) | 0,12 | 244 | 3.7 (5.1) | 0,42 | 237 | 3.6 (5.7) | 0,46 | 321 | 3.0 (4.2) | 0,39 |
| 65 - 69 | 74 | 6.3 (6.6) | 1,40 | 270 | 1.8 (1.7) | 0,11 | 245 | 3.9 (7.6) | 0,93 | 199 | 3.8 (8.3) | 0,101 | 313 | 3.0 (4.1) | 0,43 |
| 70 - 74 | 33 | 5.1 (4.7) | 1,20 | 89 | 1.8 (1.5) | 0,8 | 73 | 3.1 (3.5) | 0,25 | 79 | 3.0 (2.1) | 0,11 | 118 | 2.4 (2.2) | 0,16 |
| 75 - 79 | 14 | 4.6 (3.7) | 1,14 | 35 | 1.7 (1.4) | 0,5 | 34 | 2.6 (2.1) | 0,9 | 29 | 3.7 (5.9) | 0,32 | 48 | 2.8 (2.1) | 0,8 |
| 80 + | 13 | 3.7 (1.8) | 2,8 | 11 | 1.1 (1.0) | 0,3 | 11 | 2.0 (2.8) | 0,9 | 7 | 1.7 (1.5) | 0,4 | 10 | 1.5 (1.7) | 0,4 |
| **Sex of participants** | | | | | | | | | | | | | | | |
| Female | 722 | 7.7 (5.5) | 1,45 | 748 | 2.2 (2.0) | 0,16 | 638 | 3.9 (6.8) | 0,102 | 536 | 3.6 (5.0) | 0,46 | 901 | 3.7 (6.0) | 0,100 |
| Male | 581 | 8.3 (7.2) | 1,58 | 806 | 2.0 (1.8) | 0,12 | 717 | 3.3 (5.3) | 0,93 | 544 | 3.2 (5.9) | 0,101 | 985 | 3.0 (4.6) | 0,84 |
| **Household size** | | | | | | | | | | | | | | | |
| 1 | 250 | 6.8 (6.8) | 1,58 | 232 | 0.8 (1.5) | 0,12 | 256 | 2.1 (4.2) | 0,42 | 268 | 1.9 (3.4) | 0,42 | 487 | 2.2 (6.2) | 0,100 |
| 2 | 411 | 7.3 (6.2) | 1,55 | 412 | 1.3 (1.6) | 0,11 | 351 | 3.8 (8.4) | 0,101 | 270 | 3.4 (7.6) | 0,101 | 439 | 2.8 (5.4) | 0,84 |
| 3 | 339 | 8.2 (5.4) | 1,42 | 514 | 2.2 (1.5) | 0,11 | 447 | 3.6 (6.0) | 0,102 | 343 | 3.8 (5.1) | 0,46 | 544 | 3.5 (4.5) | 0,43 |
| 4 or more | 341 | 9.3 (6.5) | 1,50 | 402 | 3.5 (1.9) | 0,16 | 302 | 4.5 (3.5) | 0,32 | 200 | 4.5 (4.1) | 0,34 | 420 | 4.9 (4.8) | 0,39 |
| **Weekdays** | | | | | | | | | | | | | | | |
| Monday | 227 | 7.6 (4.9) | 1,31 | 50 | 2.2 (1.7) | 0,6 | 60 | 4.3 (4.1) | 0,24 | 128 | 2.7 (4.3) | 0,42 | 642 | 3.6 (5.5) | 0,84 |
| Tuesday | 237 | 7.7 (5.9) | 1,56 | 63 | 2.4 (2.0) | 0,8 | 89 | 5.0 (11.2) | 0,102 | 143 | 4.0 (4.6) | 0,32 | 246 | 4.1 (4.9) | 0,43 |
| Wednesday | 222 | 8.5 (7.7) | 1,58 | 54 | 2.3 (2.5) | 0,13 | 293 | 3.3 (4.4) | 0,42 | 87 | 3.1 (3.9) | 0,31 | 172 | 3.6 (4.9) | 0,39 |
| Thursday | 179 | 7.3 (4.9) | 1,30 | 914 | 2.1 (1.9) | 0,16 | 613 | 3.4 (5.5) | 0,93 | 489 | 3.6 (6.6) | 0,101 | 320 | 2.5 (3.4) | 0,36 |
| Friday | 186 | 8.5 (6.7) | 1,55 | 144 | 2.1 (1.8) | 0,8 | 117 | 4.4 (4.5) | 0,32 | 132 | 3.0 (3.5) | 0,27 | 196 | 3.6 (8.1) | 0,100 |
| Saturday | 152 | 8.4 (6.5) | 1,50 | 88 | 2.0 (1.6) | 0,8 | 63 | 4.2 (12.6) | 0,101 | 66 | 2.8 (4.2) | 0,33 | 81 | 3.9 (5.8) | 0,35 |
| Sunday | 117 | 7.9 (7.3) | 1,45 | 247 | 1.9 (1.9) | 0,12 | 121 | 2.5 (2.8) | 0,21 | 36 | 3.2 (5.5) | 0,34 | 233 | 2.2 (4.4) | 0,43 |

4.2.b. Table 4.2b. Number of recorded physical contacts per participant per day stratified by age, gender, household size, and day of the week in the COVIMOD survey Waves 1 to 4 and the POLYMOD survey for the unweighted analysis without group contacts.

| | POLYMOD | | | COVIMOD | | | | | | | | | | | |
|---|---|---|---|---|---|---|---|---|---|---|---|---|---|---|---|
| | | | | Wave1 | | | Wave 2 | | | Wave 3 | | | Wave 4 | | |
| | N | Mean (SD) | Min, Max | N | Mean (SD) | Min, Max | N | Mean (SD) | Min, Max | N | Mean (SD) | Min, Max | N | Mean (SD) | Min, Max |
| | **1341** | **5.5 (4.8)** | **1,45** | **1560** | **0.9 (1.3)** | **0,11** | **1356** | **1.1 (3.2)** | **0,101** | **1081** | **0.9 (1.5)** | **0,25** | **1890** | **1.1 (2.1)** | **0,39** |
| **Age category** | | | | | | | | | | | | | | | |
| 0 - 4 | 89 | 7.3 (5.0) | 0,30 | 46 | 3.3 (1.9) | 0,7 | 36 | 3.3 (2.2) | 0,8 | 21 | 3.1 (2.7) | 0,12 | 56 | 3.4 (3.4) | 0,18 |
| 5 - 9 | 92 | 6.6 (4.4) | 0,33 | 48 | 2.6 (1.7) | 0,6 | 41 | 2.7 (1.8) | 0,6 | 30 | 2.4 (1.6) | 0,5 | 62 | 3.1 (3.2) | 0,23 |
| 10 - 14 | 110 | 6.4 (4.7) | 0,25 | 73 | 2.1 (1.9) | 0,11 | 63 | 1.7 (1.8) | 0,6 | 45 | 2.1 (2.1) | 0,7 | 87 | 2.3 (2.0) | 0,8 |
| 15 - 19 | 121 | 6.7 (6.9) | 0,45 | 95 | 1.4 (1.6) | 0,8 | 66 | 1.3 (1.6) | 0,6 | 45 | 1.1 (1.3) | 0,4 | 108 | 1.5 (2.7) | 0,22 |
| 20 - 24 | 117 | 5.2 (4.1) | 0,26 | 83 | 0.8 (1.1) | 0,5 | 60 | 0.7 (1.0) | 0,3 | 28 | 0.7 (0.9) | 0,3 | 109 | 1.7 (3.6) | 0,33 |
| 25 - 34 | 132 | 5.4 (3.8) | 0,29 | 173 | 0.8 (1.0) | 0,6 | 148 | 0.7 (0.9) | 0,5 | 96 | 0.6 (0.8) | 0,4 | 219 | 0.9 (1.3) | 0,8 |
| 35 - 44 | 156 | 5.8 (4.9) | 0,36 | 137 | 0.7 (1.2) | 0,8 | 124 | 0.8 (1.2) | 0,6 | 91 | 0.8 (1.3) | 0,8 | 164 | 0.6 (1.0) | 0,6 |
| 45 - 54 | 184 | 5.2 (4.8) | 0,35 | 235 | 0.7 (1.0) | 0,4 | 209 | 1.5 (7.2) | 0,101 | 174 | 0.8 (1.2) | 0,7 | 275 | 0.8 (1.3) | 0,13 |
| 55 - 64 | 160 | 4.0 (3.2) | 0,16 | 265 | 0.7 (0.9) | 0,5 | 244 | 0.8 (1.1) | 0,10 | 237 | 0.7 (1.0) | 0,5 | 321 | 0.9 (2.5) | 0,39 |
| 65 - 69 | 74 | 4.6 (6.3) | 0,40 | 270 | 0.6 (0.9) | 0,5 | 245 | 0.9 (2.5) | 0,29 | 199 | 0.9 (2.1) | 0,25 | 313 | 0.8 (1.3) | 0,11 |
| 70 - 74 | 33 | 2.6 (3.0) | 0,13 | 89 | 0.7 (0.8) | 0,3 | 73 | 0.8 (0.9) | 0,3 | 79 | 0.8 (1.0) | 0,4 | 118 | 0.6 (0.9) | 0,4 |
| 75 - 79 | 14 | 2.7 (2.6) | 0,8 | 35 | 0.8 (0.9) | 0,3 | 34 | 0.6 (1.0) | 0,3 | 29 | 0.7 (0.8) | 0,3 | 48 | 0.8 (0.9) | 0,3 |
| 80 + | 13 | 2.8 (1.8) | 1,6 | 11 | 0.3 (0.5) | 0,1 | 11 | 0.2 (0.4) | 0,1 | 7 | 0.3 (0.8) | 0,2 | 10 | 0.3 (0.5) | 0,1 |
| **Sex of participants** | | | | | | | | | | | | | | | |
| Female | 722 | 5.3 (4.8) | 0,45 | 748 | 0.9 (1.3) | 0,8 | 638 | 1.3 (4.3) | 0,101 | 536 | 0.9 (1.7) | 0,25 | 901 | 1.1 (2.1) | 0,39 |
| Male | 581 | 5.6 (4.9) | 0,44 | 806 | 0.9 (1.3) | 0,11 | 717 | 0.9 (1.7) | 0,29 | 544 | 0.9 (1.2) | 0,7 | 985 | 1.1 (2.0) | 0,33 |
| **Household size** | | | | | | | | | | | | | | | |
| 1 | 250 | 4.0 (4.4) | 0,36 | 232 | 0.1 (0.5) | 0,5 | 256 | 0.2 (0.6) | 0,4 | 268 | 0.2 (0.7) | 0,7 | 487 | 0.3 (0.9) | 0,8 |
| 2 | 411 | 4.9 (4.5) | 0,45 | 412 | 0.5 (0.7) | 0,6 | 351 | 1.1 (5.8) | 0,101 | 270 | 0.8 (1.8) | 0,25 | 439 | 0.8 (1.9) | 0,33 |
| 3 | 339 | 5.8 (4.2) | 0,30 | 514 | 0.9 (1.0) | 0,8 | 447 | 1.1 (1.2) | 0,13 | 343 | 1.1 (1.2) | 0,12 | 544 | 1.2 (2.2) | 0,39 |
| 4 or more | 341 | 6.7 (5.6) | 0,44 | 402 | 1.8 (1.8) | 0,11 | 302 | 1.8 (2.1) | 0,13 | 200 | 1.7 (1.8) | 0,8 | 420 | 2.1 (2.6) | 0,23 |
| **Weekdays** | | | | | | | | | | | | | | | |
| Monday | 227 | 5.1 (4.2) | 0,29 | 50 | 1.4 (1.6) | 0,6 | 60 | 1.1 (1.4) | 0,6 | 128 | 0.9 (1.1) | 0,5 | 642 | 1.2 (2.2) | 0,33 |
| Tuesday | 237 | 5.3 (4.4) | 0,40 | 63 | 1.0 (1.5) | 0,7 | 89 | 1.2 (1.8) | 0,13 | 143 | 0.9 (1.4) | 0,6 | 246 | 1.3 (2.9) | 0,39 |
| Wednesday | 222 | 5.6 (5.1) | 0,36 | 54 | 0.7 (1.2) | 0,6 | 293 | 0.8 (1.7) | 0,22 | 87 | 0.9 (1.3) | 0,6 | 172 | 1.3 (2.4) | 0,22 |
| Thursday | 179 | 5.4 (4.6) | 0,30 | 914 | 0.8 (1.2) | 0,8 | 613 | 1.0 (1.8) | 0,29 | 489 | 0.9 (1.7) | 0,25 | 320 | 0.8 (1.2) | 0,6 |
| Friday | 186 | 5.6 (4.2) | 0,23 | 144 | 1.0 (1.3) | 0,6 | 117 | 1.2 (1.9) | 0,12 | 132 | 0.9 (1.5) | 0,12 | 196 | 0.9 (2.0) | 0,23 |
| Saturday | 152 | 6.0 (5.4) | 0,44 | 88 | 1.1 (1.6) | 0,8 | 63 | 2.7 (12.7) | 0,101 | 66 | 0.9 (1.2) | 0,5 | 81 | 1.5 (2.2) | 0,13 |
| Sunday | 117 | 6.0 (6.7) | 0,45 | 247 | 1.0 (1.5) | 0,11 | 121 | 1.0 (1.3) | 0,5 | 36 | 0.8 (1.2) | 0,5 | 233 | 0.8 (1.3) | 0,8 |

4.2.c. Table 4.2c. Number of recorded overall contacts per different settings in the COVIMOD survey Waves 1 to 4 and the POLYMOD survey for the unweighted analysis without group contacts.

**Note:** *The displayed educational contacts are based on the group of participants who attended an educational facility (kindergarten, school, university) and work contacts are based on the group of participants who reported to work full-/part-time.*

| | POLYMOD | | | | | | | | | | | | | | |
|---|---|---|---|---|---|---|---|---|---|---|---|---|---|---|---|
| | | | | Wave 1 | | | Wave 2 | | | Wave 3 | | | Wave 4 | | |
| | N | Mean (SD) | Min, Max | N | Mean (SD) | Min, Max | N | Mean (SD) | Min, Max | N | Mean (SD) | Min, Max | N | Mean (SD) | Min, Max |
| Overall | 1341 | 7.9 (6.3) | 1,58 | 1560 | 2.1 (1.9) | 0,16 | 1356 | 3.6 (6.1) | 0,102 | 1081 | 3.4 (5.4) | 0,101 | 1890 | 3.3 (5.3) | 0,100 |
| Home | 1341 | 2.8 (2.3) | 0,26 | 1560 | 1.6 (1.5) | 0,12 | 1356 | 1.6 (1.6) | 0,9 | 1081 | 1.5 (1.5) | 0,13 | 1890 | 1.5 (1.7) | 0,23 |
| Educational | 199 | 2.8 (4.7) | 0,42 | 310 | 0.0 (0.2) | 0,3 | 247 | 0.2 (1.0) | 0,10 | 179 | 0.2 (1.6) | 0,17 | 385 | 0.7 (2.5) | 0,22 |
| Work | 715 | 2.4 (5.2) | 0,56 | 690 | 0.4 (1.2) | 0,11 | 613 | 2.0 (7.2) | 0,100 | 476 | 1.8 (6.3) | 0,97 | 809 | 1.6 (5.7) | 0,100 |
| Transport | 1341 | 0.3 (0.8) | 0,8 | 1560 | 0.0 (0.3) | 0,3 | 1356 | 0.1 (0.4) | 0,4 | 1081 | 0.1 (0.4) | 0,8 | 1890 | 0.1 (0.6) | 0,12 |
| Others | 1341 | 3.0 (3.8) | 0,45 | 1560 | 0.4 (1.0) | 0,10 | 1356 | 1.1 (2.4) | 0,33 | 1081 | 1.1 (2.9) | 0,35 | 1890 | 1.1 (2.5) | 0,38 |

4.2.d. Table 2d. Number of recorded physical contacts per different settings in the COVIMOD survey Waves 1 to 4 and the POLYMOD survey for the unweighted analysis without group contacts.

**Note:** *The displayed educational contacts are based on the group of participants who attended an educational facility (kindergarten, school, university) and work contacts are based on the group of participants who reported to work full-/part-time.*

| | POLYMOD | | | COVIMOD | | | | | | | | | | | |
|---|---|---|---|---|---|---|---|---|---|---|---|---|---|---|---|
| | | | | Wave 1 | | | Wave 2 | | | | Wave 3 | | Wave 4 | | |
| | N | Mean (SD) | Min, Max | N | Mean (SD) | Min, Max | N | Mean (SD) | Min, Max | N | Mean (SD) | Min, Max | N | Mean (SD) | Min, Max |
| Overall | 1341 | 5.5 (4.8) | 0,45 | 1560 | 0.9 (1.3) | 0,11 | 1356 | 1.1 (3.2) | 0,101 | 1081 | 0.9 (1.5) | 0,25 | 1890 | 1.1 (2.1) | 0,39 |
| Home | 1341 | 2.3 (2.2) | 0,26 | 1560 | 0.8 (1.3) | 0,11 | 1356 | 0.8 (1.2) | 0, 8 | 1081 | 0.8 (1.2) | 0,12 | 1890 | 0.8 (1.2) | 0, 8 |
| Educational | 199 | 1.5 (3.8) | 0,42 | 310 | 0.0 (0.1) | 0, 1 | 247 | 0.0 (0.3) | 0, 4 | 179 | 0.0 (0.2) | 0, 2 | 385 | 0.2 (1.5) | 0,21 |
| Work | 715 | 1.4 (3.0) | 0,30 | 690 | 0.0 (0.2) | 0, 2 | 613 | 0.4 (4.4) | 0,100 | 476 | 0.1 (1.3) | 0,25 | 809 | 0.2 (1.5) | 0,31 |
| Transport | 1341 | 0.2 (0.6) | 0, 6 | 1560 | 0.0 (0.2) | 0, 3 | 1356 | 0.0 (0.3) | 0, 3 | 1081 | 0.0 (0.2) | 0, 3 | 1890 | 0.0 (0.3) | 0, 7 |
| Others | 1341 | 2.1 (3.2) | 0,40 | 1560 | 0.1 (0.5) | 0, 5 | 1356 | 0.2 (0.7) | 0, 11 | 1081 | 0.2 (0.7) | 0, 7 | 1890 | 0.3 (1.2) | 0,31 |

### 4.3. Reproduction number estimates of SARS-CoV-2 under contact reduction measures.

4.3.a. Table 4.3. Effective reproduction number at the timing of COVIMOD survey Waves 1 to 4, estimated effective reproduction number based on the reduction of social contacts at the times of COVIMOD survey Waves 1 to 4 assuming values of the basic reproduction number of Norm (2.6, SD=0.54) and the reduction in mobility at the times of the COVIMOD survey Waves, with 10000 bootstrapped samples in various settings and in different time frames for the unweighted analysis without group contacts.

**Note:** *RKI - the percentage mean and minimum and maximum reductions in the reproduction number in the assessed time intervals, COVIMOD - the calculated percentage mean reductions in the reproduction number and the 95% confidence interval, Google and Apple mobility – the percentage mean and minimum and maximum reduction in mobility in the assessed time intervals.*

| | COVIMOD | | RKI | | Google | | Apple | |
|---|---|---|---|---|---|---|---|---|
| | Mean reproduction number (Min, Max) | Percent Mean Reduction Number (Min, Max) | Mean reproduction number (Min, Max) | Percent Mean Reduction Number (Min, Max) | Mean reproduction number (Min, Max) | Percent Mean Reduction Number (Min, Max) | Mean reproduction number (Min, Max) | Percent Mean Reduction Number (Min, Max) |
| **Overall** | | | | | | | | |
| 30th April - 6th May 2020 | 0.70 (0.42, 0.99) | -73.00 (-83.00, -61.00) | 0.88 (0.79, 0.97) | -66.00 (-69.00, -62.00) | - | -24.31 (-59.40, -15.20) | - | -36.77 (-44.17, -29.61) |
| 14th May - 21st May 2020 | 0.99 (0.59, 1.41) | -61.00 (-77.00, -45.00) | 0.90 (0.72, 1.15) | -65.00 (-72.00, -55.00) | - | -16.85 (-50.20, -3.40) | - | -15.64 (-25.26, -2.58) |
| 28th May - 4th June 2020 | 0.96 (0.56, 1.37) | -63.00 (-78.00, -47.00) | 1.06 (0.84, 1.39) | -59.00 (-67.00, -46.00) | - | -13.05 (-47.40, 1.60) | - | 5.18 (-4.00, 19.43) |
| 11th June - 22nd June 2020 | 1.00 (0.59, 1.44) | -61.00 (-77.00, -44.00) | 0.84 (0.60, 1.07) | -67.00 (-76.00, -58.00) | - | -10.63 (-34.60, 5.60) | - | 11.66 (1.62, 28.00) |
| **Home** | | | | | | | | |
| 30th April - 6th May 2020 | 1.60 (0.95, 2.27) | -38.00 (-63.00, -12.00) | - | - | - | 12.71 (6.00, 28.00) | - | - |
| 14th May - 21st May 2020 | 1.58 (0.93, 2.25) | -39.00 (-64.00, -13.00) | - | - | - | 8.62 (3.00, 18.00) | - | - |
| 28th May - 4th June 2020 | 1.56 (0.91, 2.26) | -40.00 (-64.00, -13.00) | - | - | - | 6.88 (0.00, 16.00) | - | - |
| 11th June - 22nd June 2020 | 1.41 (0.84, 2.01) | -45.00 (-67.00, -22.00) | - | - | - | 5.25 (-1.00, 16.00) | - | - |
| **Educational** | | | | | | | | |
| 30th April - 6th May 2020 | 0.07 (0.02, 0.17) | -97.00 (-99.00, -93.00) | - | - | - | - | - | - |
| 14th May - 21st May 2020 | 0.34 (0.12, 0.76) | -86.00 (-95.00, -70.00) | - | - | - | - | - | - |
| 28th May - 4th June 2020 | 0.42 (0.08, 1.23) | -83.00 (-96.00, -52.00) | - | - | - | - | - | - |
| 11th June - 22nd June 2020 | 1.10 (0.49, 1.99) | -57.00 (-81.00, -23.00) | - | - | - | - | - | - |
| **Work** | | | | | | | | |
| 30th April - 6th May 2020 | 0.28 (0.15, 0.43) | -89.00 (-94.00, -83.00) | - | - | - | -34.71 (-84.00, -18.00) | - | - |
| 14th May - 21st May 2020 | 0.96 (0.53, 1.53) | -63.00 (-79.00, -41.00) | - | - | - | -27.00 (-81.00, -1.00) | - | - |
| 28th May - 4th June 2020 | 0.96 (0.49, 1.58) | -63.00 (-81.00, -39.00) | - | - | - | -24.12 (-80.00, 8.00) | - | - |
| 11th June - 22nd June 2020 | 1.03 (0.55, 1.70) | -60.00 (-78.00, -34.00) | - | - | - | -16.50 (-59.00, 15.00) | - | - |
| **Transport** | | | | | | | | |
| 30th April - 6th May 2020 | 0.64 (0.31, 1.12) | -75.00 (-88.00, -56.00) | - | - | - | -42.14 (-67.00, -35.00) | - | - |
| 14th May - 21st May 2020 | 0.91 (0.44, 1.64) | -64.00 (-83.00, -36.00) | - | - | - | -31.75 (-46.00, -25.00) | - | - |
| 28th May - 4th June 2020 | 0.71 (0.33, 1.31) | -72.00 (-87.00, -49.00) | - | - | - | -25.12 (-42.00, -13.00) | - | - |
| 11th June - 22nd June 2020 | | | | | | | | |
| **Others** | | | | | | | | |
| 30th April - 6th May 2020 | 0.40 (0.22, 0.64) | -84.00 (-91.00, -75.00) | - | - | - | -28.71 (-87.00, -7.50) | - | - |
| 14th May - 21st May 2020 | 0.72 (0.42, 1.04) | -72.00 (-83.00, -60.00) | - | - | - | -17.06 (-71.00, 14.00) | - | - |
| 28th May - 4th June 2020 | 0.91 (0.49, 1.55) | -64.00 (-81.00, -40.00) | - | - | - | -11.44 (-65.50, 6.50) | - | - |
| 11th June - 22nd June 2020 | 0.76 (0.44, 1.12) | -70.00 (-83.00, -56.00) | - | - | - | -9.46 (-45.00, 12.50) | - | - |

4.4. **Figure 4.4. Boxplots of the number of overall contacts during the POLYMOD and COVIMOD survey waves 1 to 4 in various settings. The boundaries of the boxes closest to zero indicates the 25th percentiles, the lines within the boxes marks the medians, the red dots within the boxes marks the means, and the boundary of the boxes farthest from zero indicates the 75th percentiles. Whiskers above and below the boxes indicate the 10th and 90th percentiles; black dots represent outliers. Participants with no contacts are displayed as 0 on the log-scale of the y-axis.**

4.4.a. Figure 4.4a. Displayed are (A) the overall number of contacts, (B) home contacts, (C) work contacts, (D) educational contacts, (E) public transport contacts and (F) other contacts for the unweighted analysis without group contacts in overall contacts.

**Note:** *The displayed educational contacts are based on the group of participants who attended an educational facility (kindergarten, school, university) and work contacts are based on the group of participants who reported to work full-/part-time.*

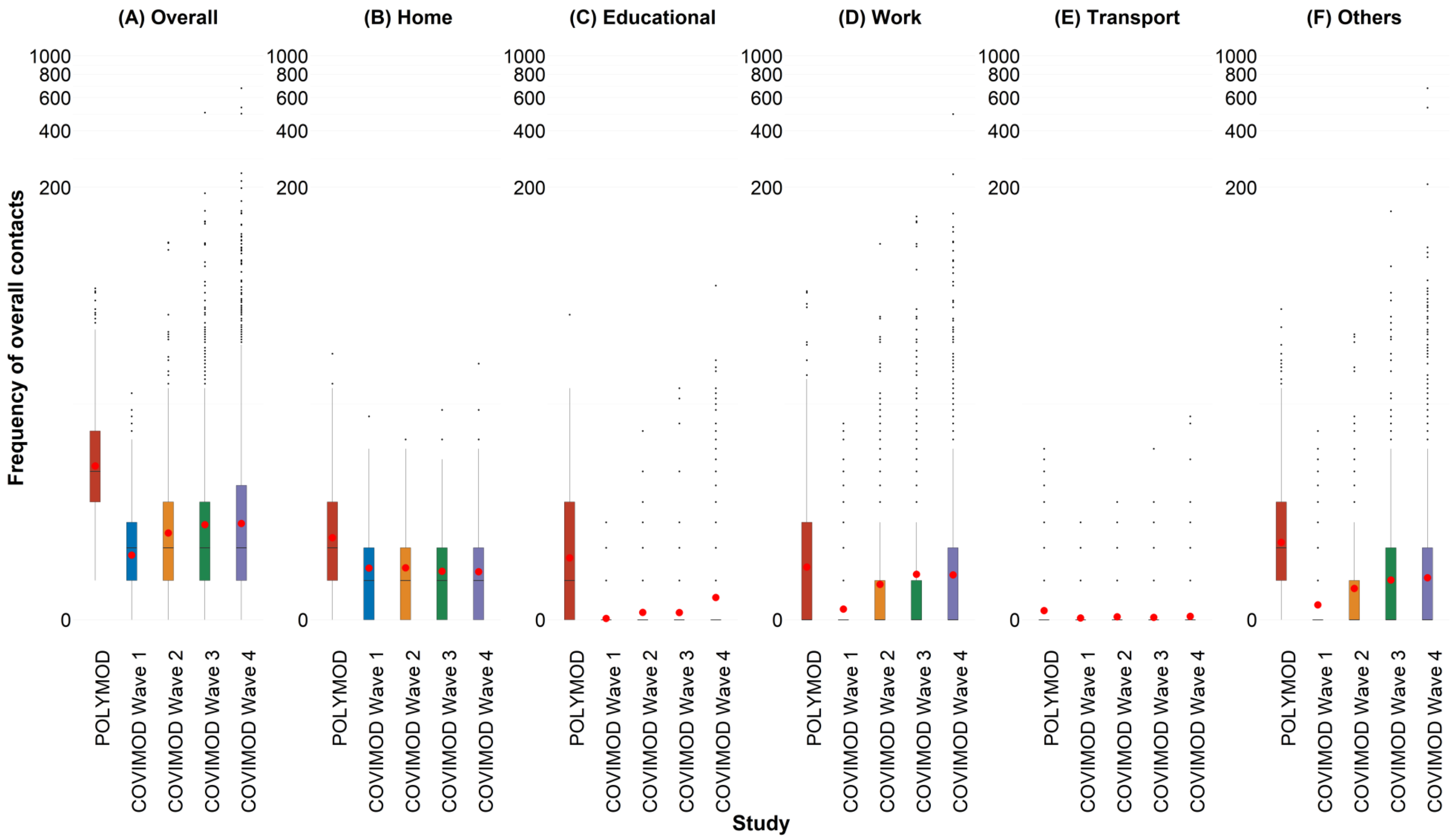

4.4.b. Figure 4.4b. Displayed are (A) 0 - 7 years, (B) 8 – 15 years, (C) 16 – 23 years, (D) 24 – 38 years, (E) 39 – 60 years and (F) 61 years or more for the unweighted analysis without group contacts in overall contacts.

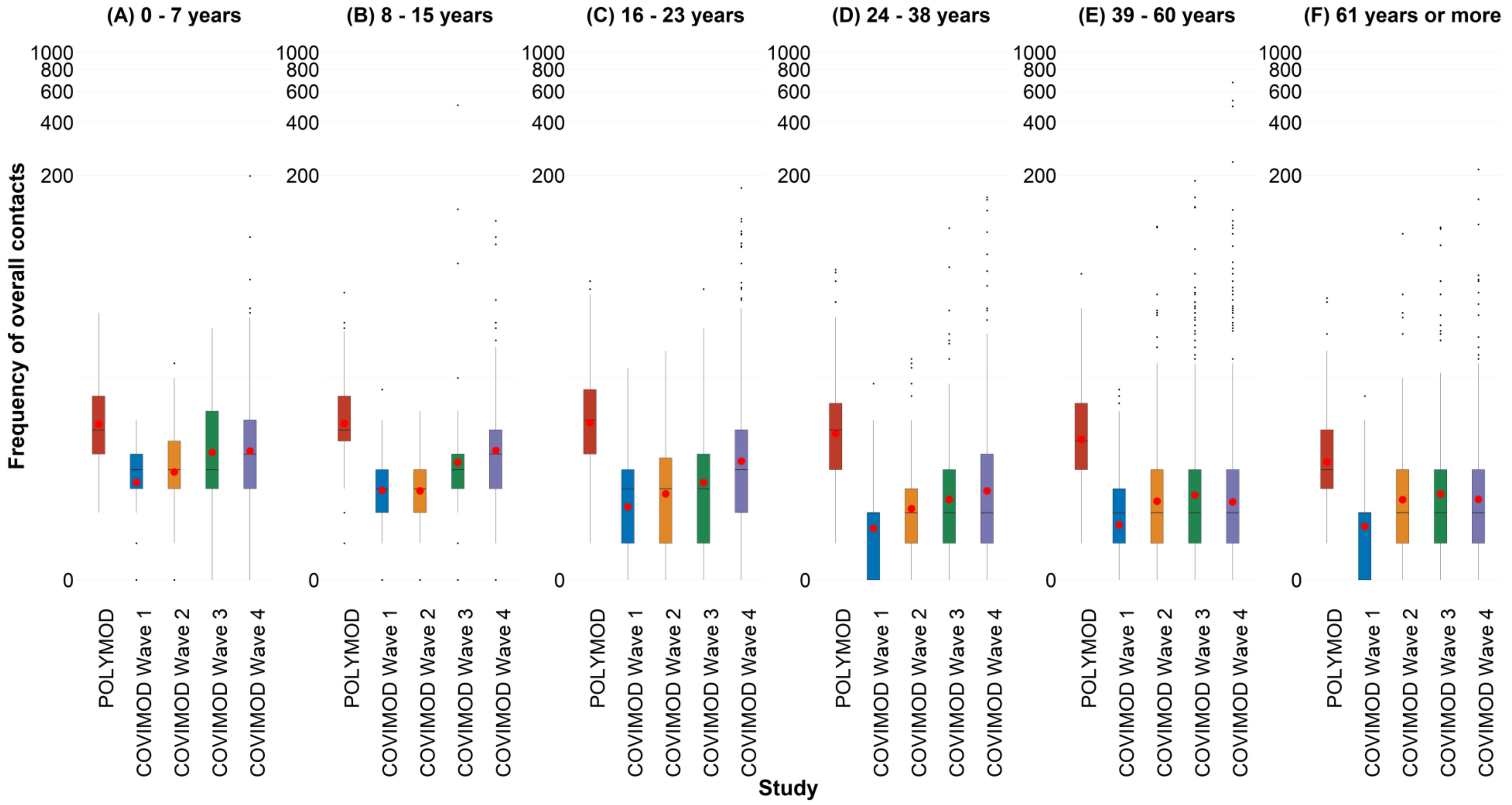

4.4.c. Figure 4.4c. Displayed are (A) Household size of 1, (B) Household size of 2, (C) Household size of 3 and (D) Household size of 4 or more for the unweighted analysis without group contacts in overall contacts.

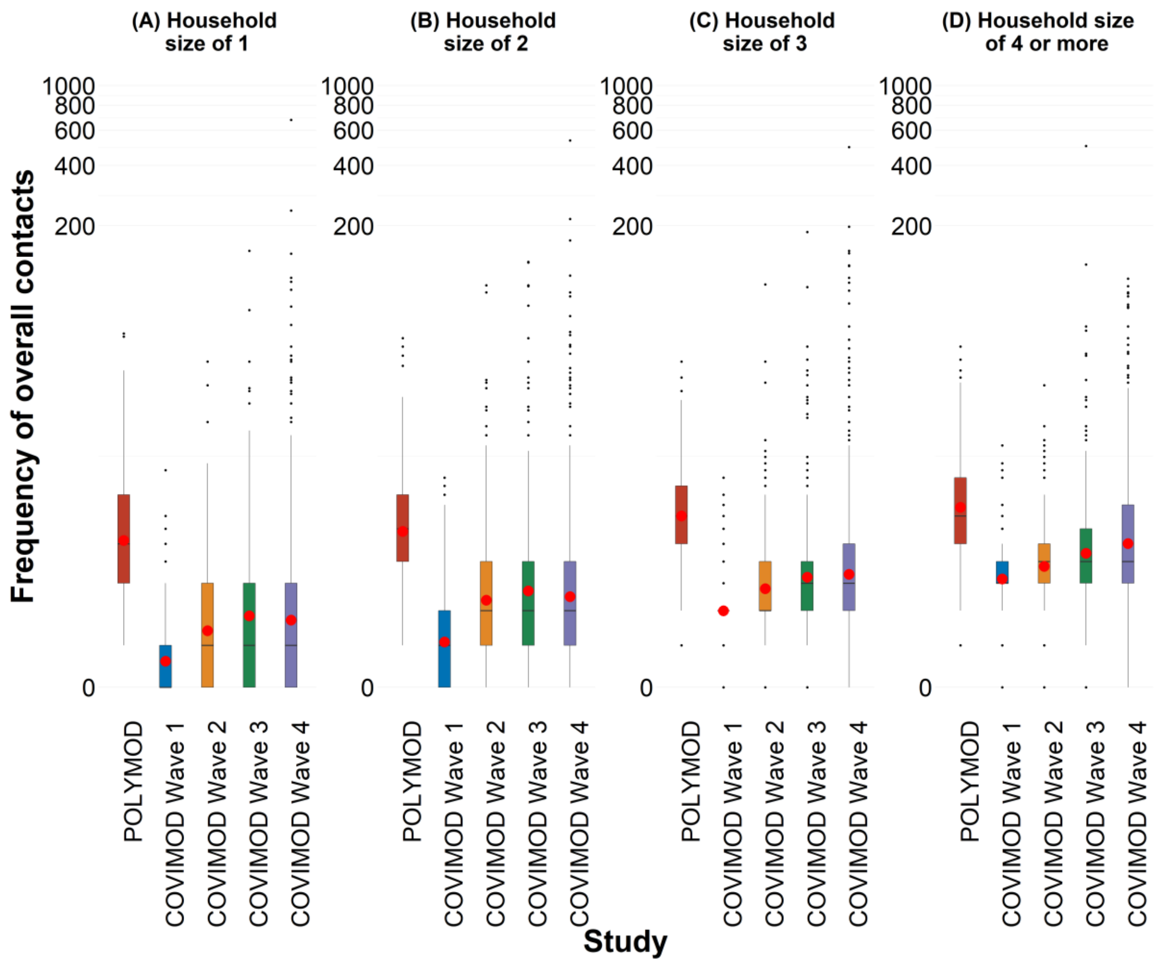

4.4.d. Figure 4.4d. Displayed are (A) Female and (B) Male for the unweighted analysis without group contacts in overall contacts.

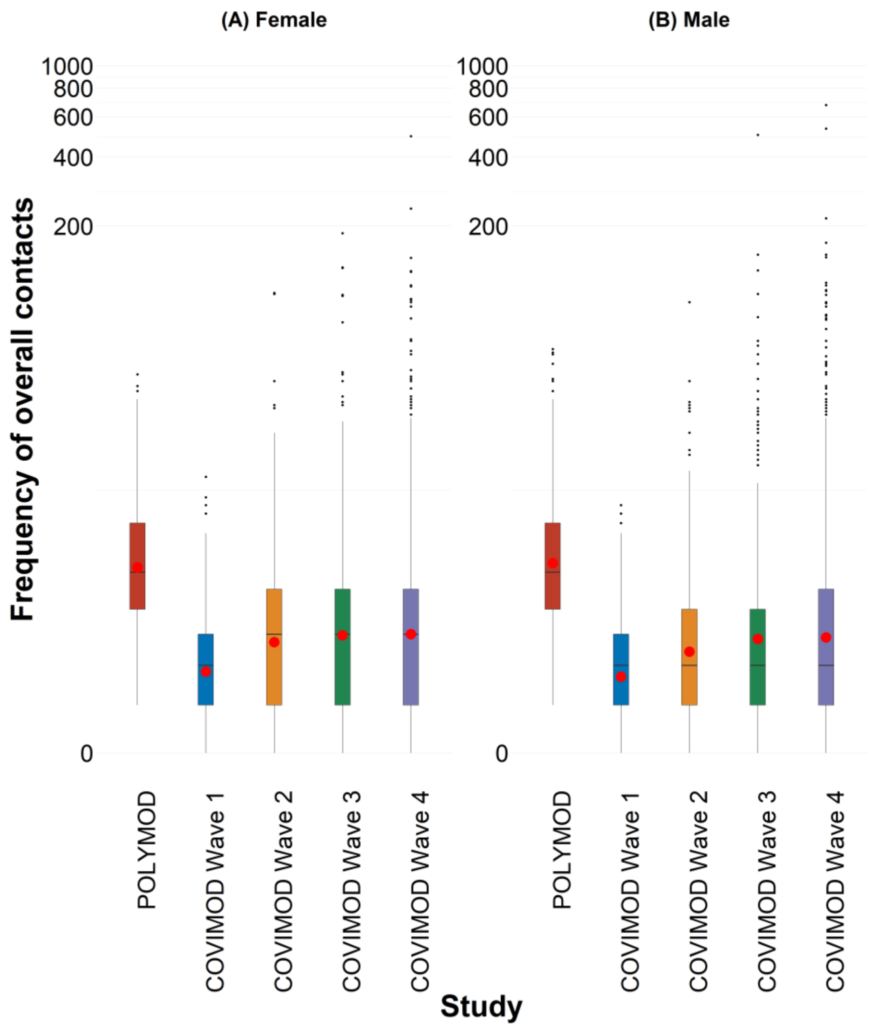

4.4.e. Figure 4.4e. Displayed are (A) Monday, (B) Tuesday, (C) Wednesday, (D) Thursday, (E) Friday, (F) Saturday and (G) Sunday for the unweighted analysis without group contacts in overall contacts.

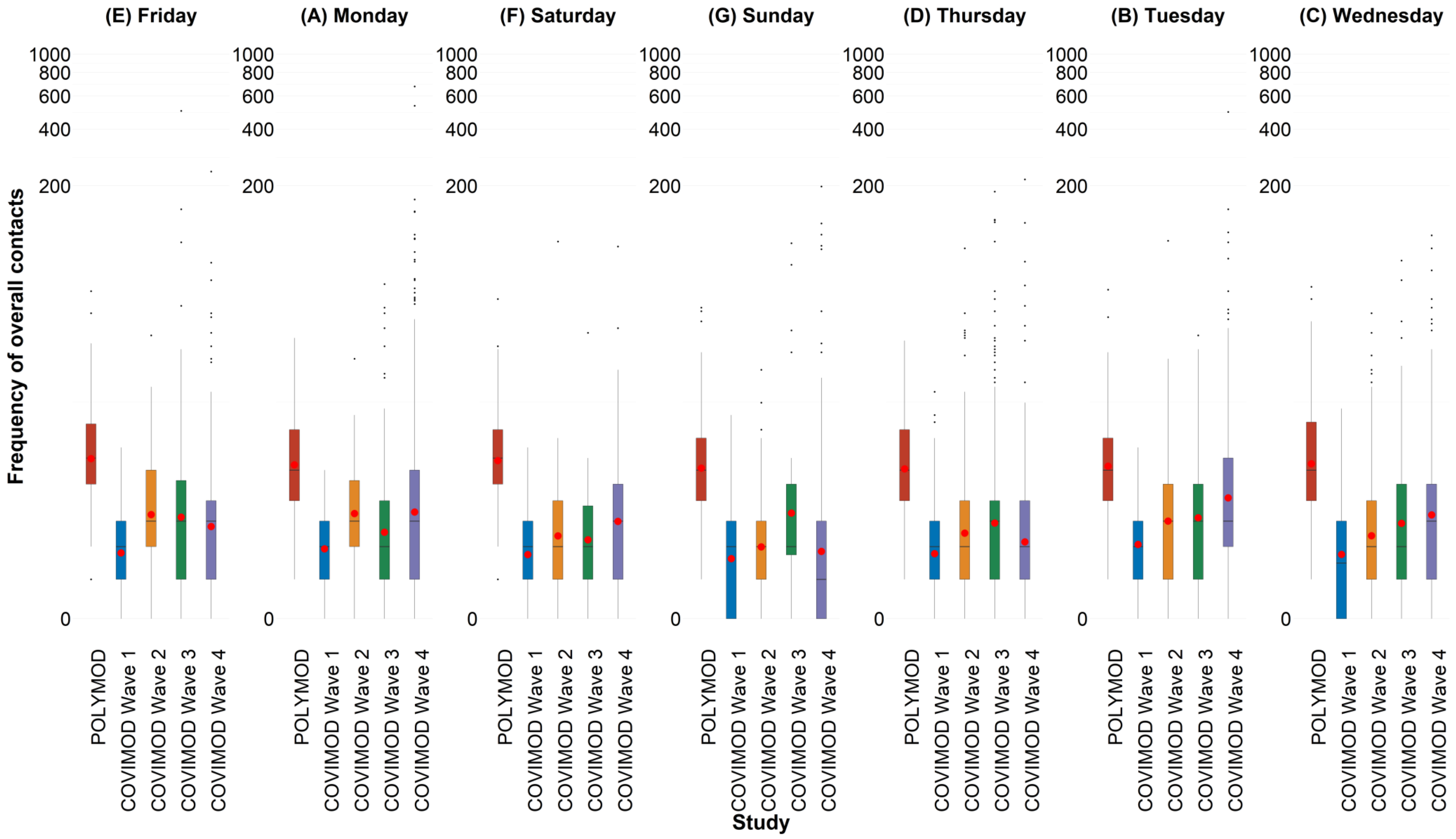

4.5. **Figure 4.5. Boxplots of the number of physical contacts during the POLYMOD and COVIMOD survey waves 1 to 4 in various settings. The boundaries of the boxes closest to zero indicates the 25th percentiles, the lines within the boxes marks the medians, the red dots within the boxes marks the means, and the boundary of the boxes farthest from zero indicates the 75th percentiles. Whiskers above and below the boxes indicate the 10th and 90th percentiles; black dots represent outliers. Participants with no contacts are displayed as 0 on the log-scale of the y-axis.**

4.5.a. Figure 4.5a. Displayed are (A) the overall number of contacts, (B) home contacts, (C) work contacts, (D) educational contacts, (E) public transport contacts and (F) other contacts for the unweighted analysis without group contacts in physical contacts.

**Note:** *The displayed educational contacts are based on the group of participants who attended an educational facility (kindergarten, school, university) and work contacts are based on the group of participants who reported to work full-/part-time.*

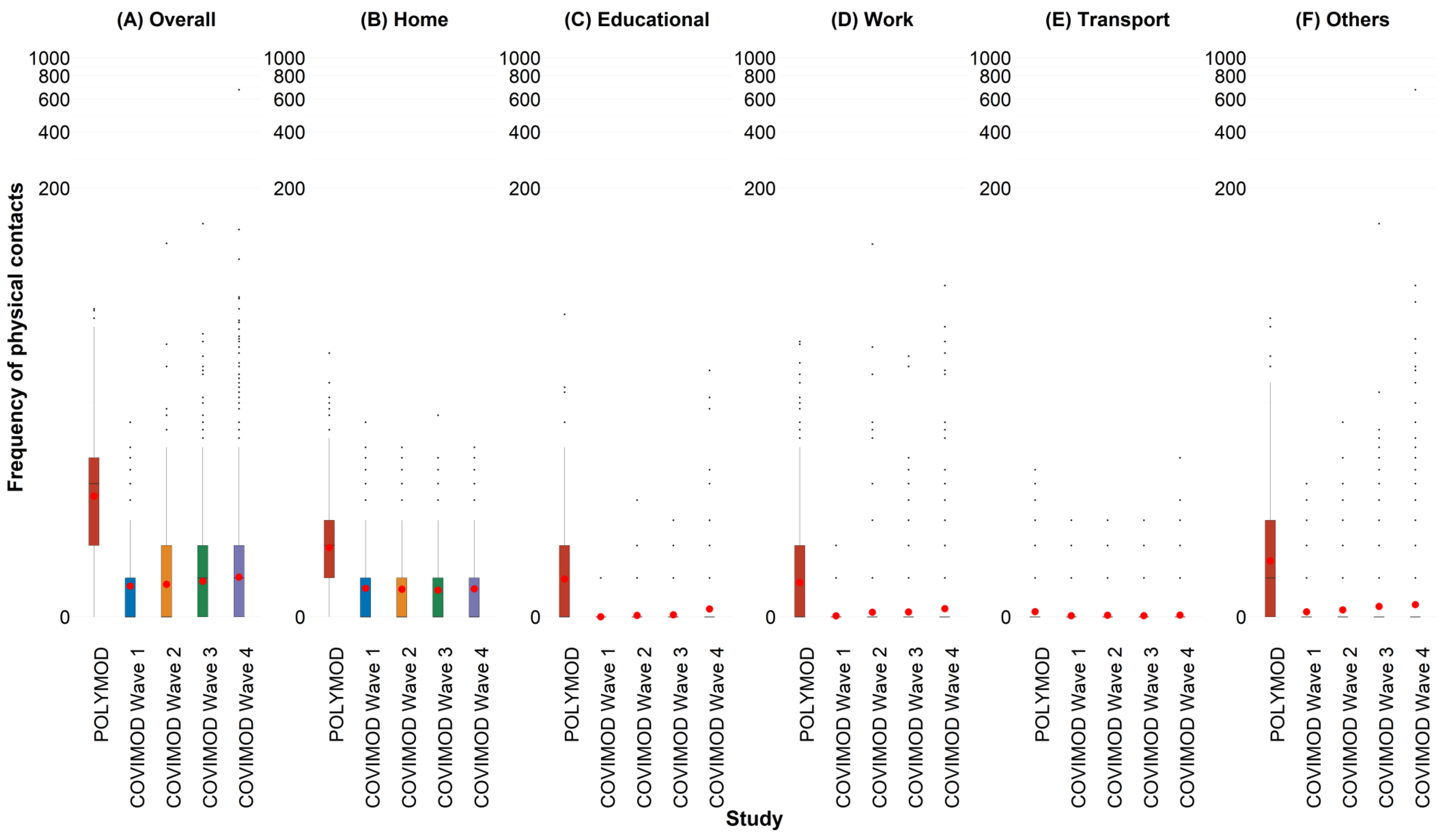

4.5.b. Figure 4.5b. Displayed are (A) 0 - 7 years, (B) 8 – 15 years, (C) 16 – 23 years, (D) 24 – 38 years, (E) 39 – 60 years and (F) 61 years or more for the unweighted analysis without group contacts in physical contacts.

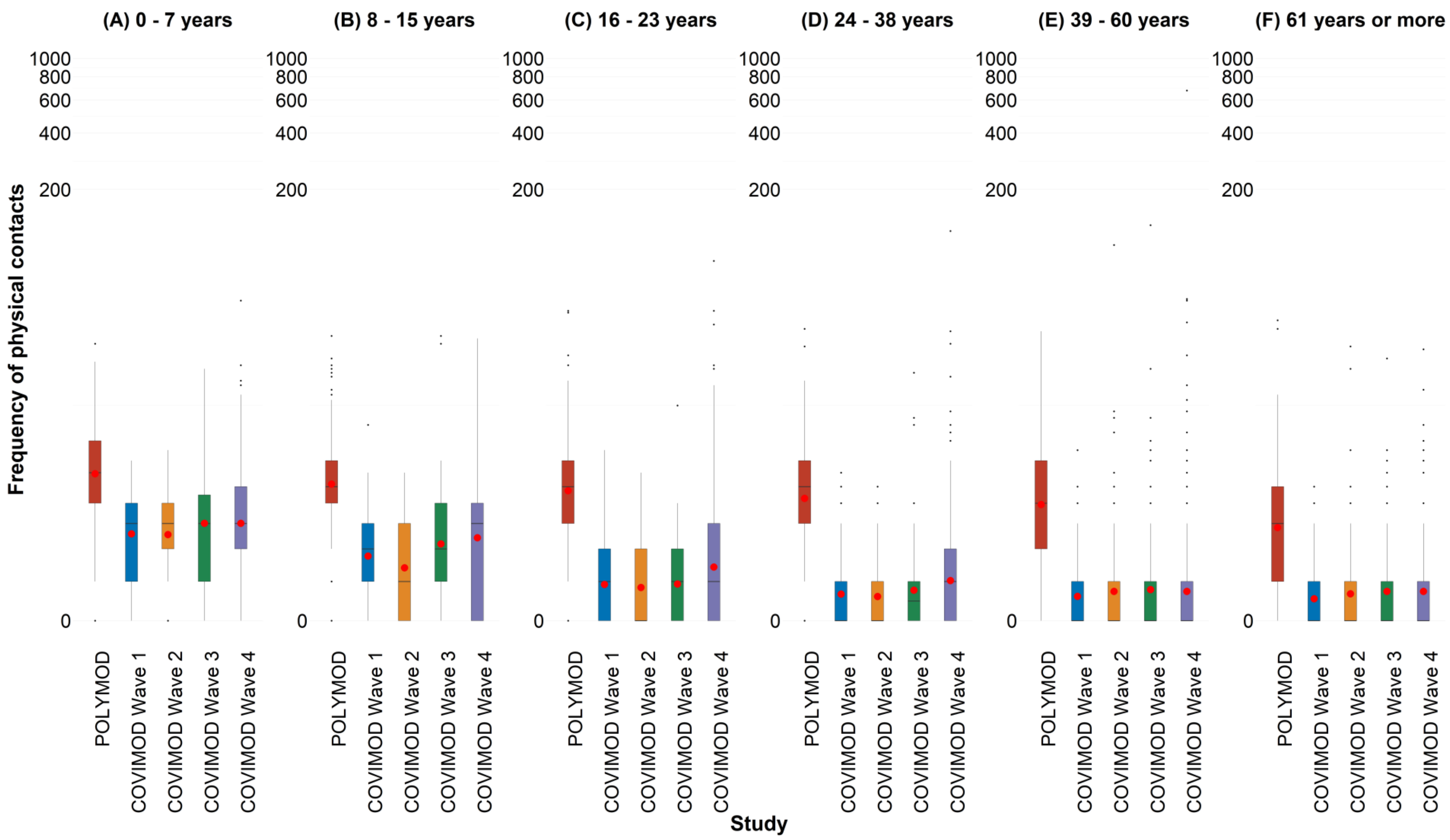

4.5.c. Figure 4.5c. Displayed are (A) Household size of 1, (B) Household size of 2, (C) Household size of 3 and (D) Household size of 4 or more for the unweighted analysis without group contacts in physical contacts.

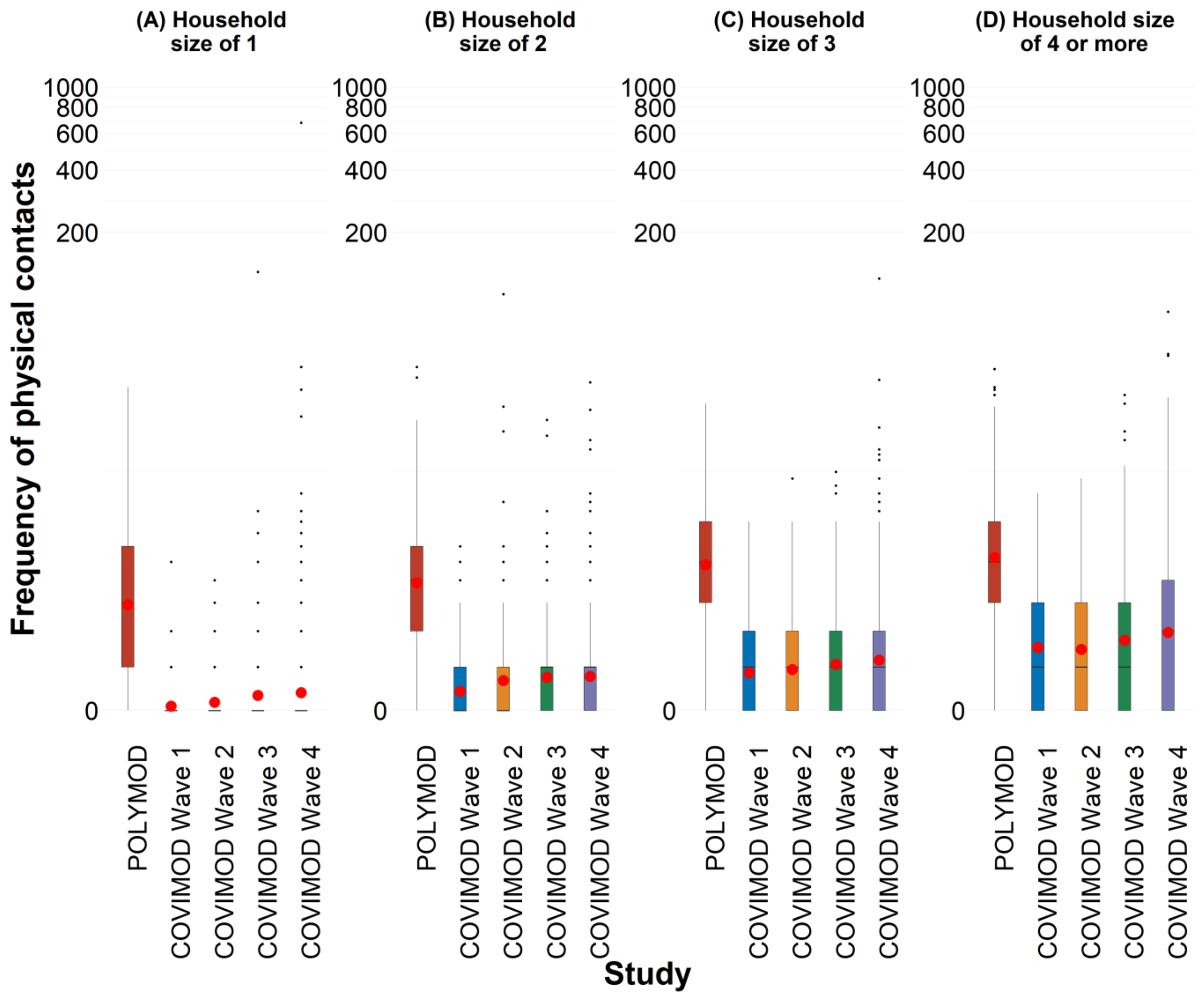

4.5.d. Figure 4.5d. Displayed are (A) Female and (B) Male for the unweighted analysis without group contacts in physical contacts.

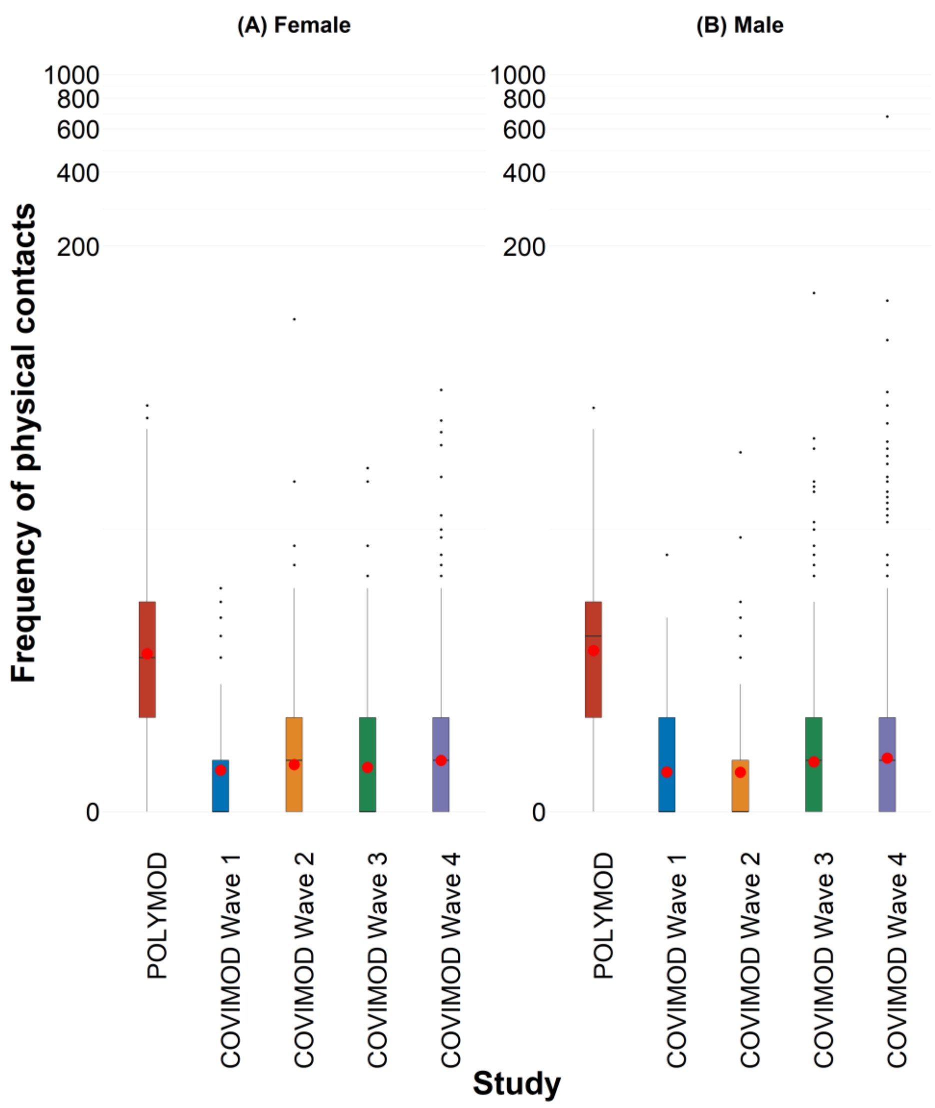

4.5.e. Figure 4.5e. Displayed are (A) Monday, (B) Tuesday, (C) Wednesday, (D) Thursday, (E) Friday, (F) Saturday and (G) Sunday for the unweighted analysis without group contacts in physical contacts.

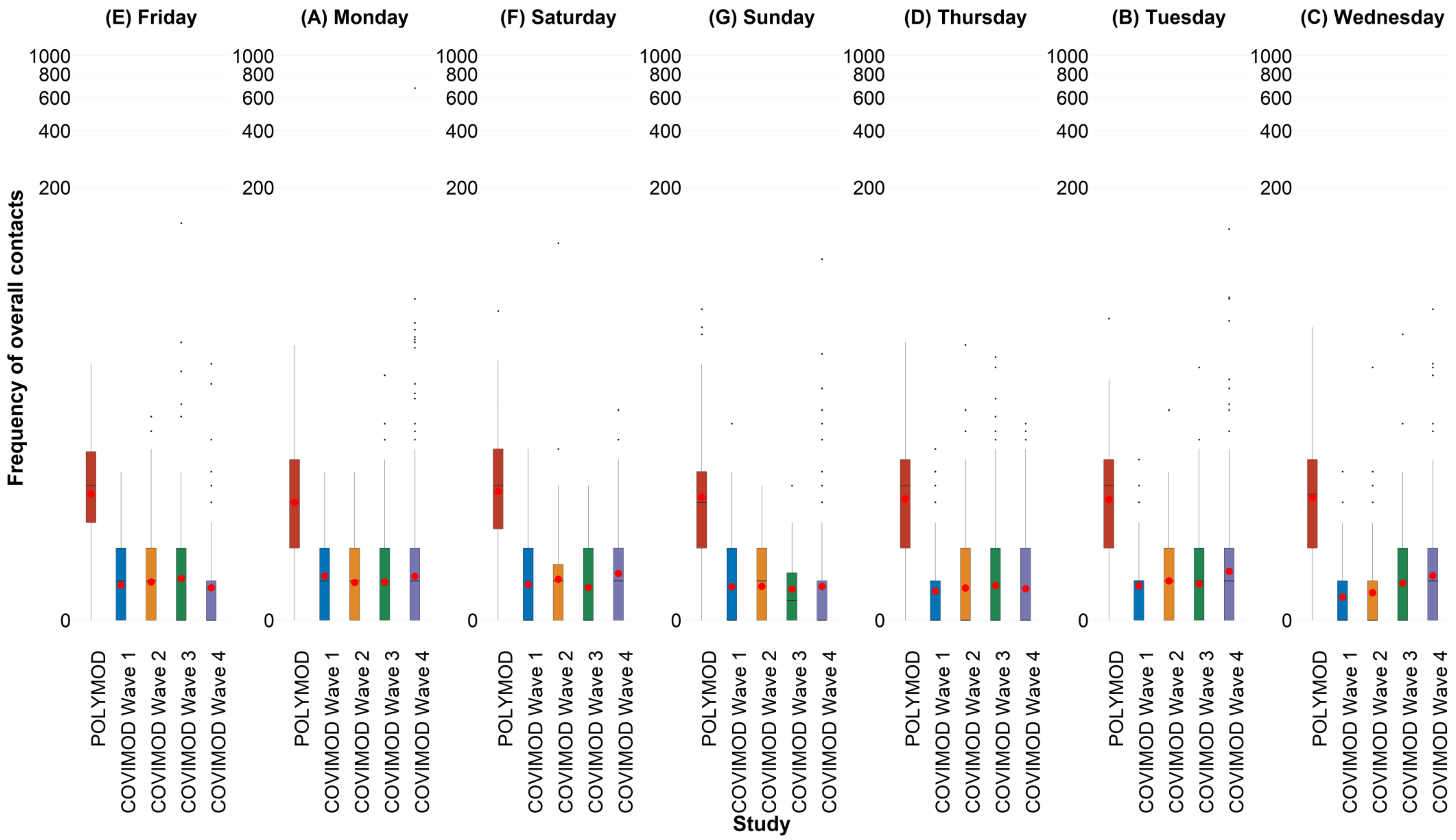

4.6. **Social contact patterns.**

4.6.a. Figure 4.6a. Social contact matrices with the mean total number of reported daily social contacts by participants in different age groups with individuals in other age groups in POLYMOD and COVIMOD survey waves 1 to 4 in various settings. Displayed are (A) the overall number of contacts, (B) home contacts, (C) educational contacts, (D) work contacts, (E) public transport contacts and (F) other contacts for the unweighted analysis without group contacts.

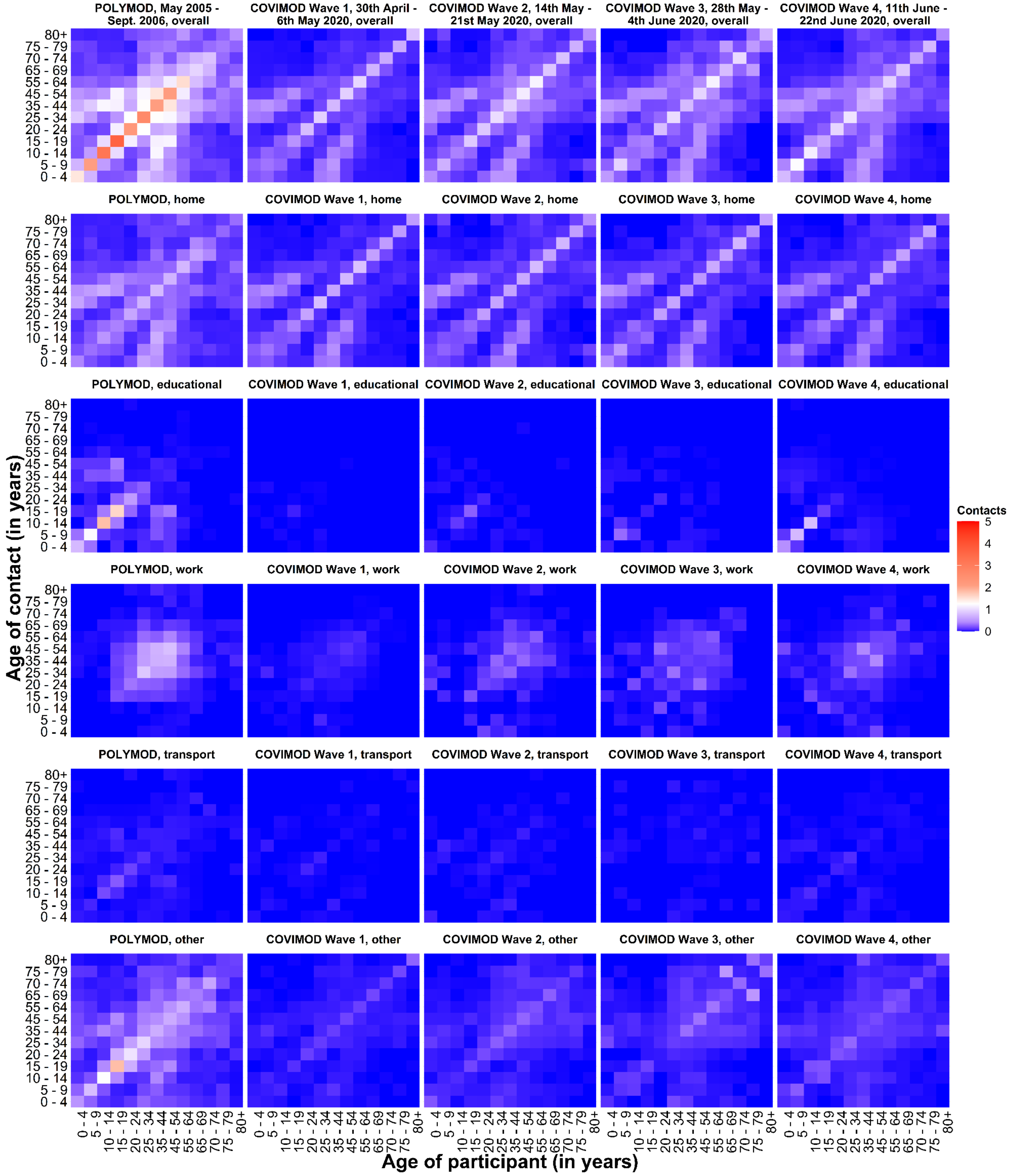

4.6.b. Figure 4.6b. Social contact matrices with the mean physical number of reported daily social contacts by participants in different age groups with individuals in other age groups in POLYMOD and COVIMOD survey waves 1 to 4 in various settings. Displayed are (A) the overall number of contacts, (B) home contacts, (C) educational contacts, (D) work contacts, (E) public transport contacts and (F) other contacts for the unweighted analysis without group contacts.

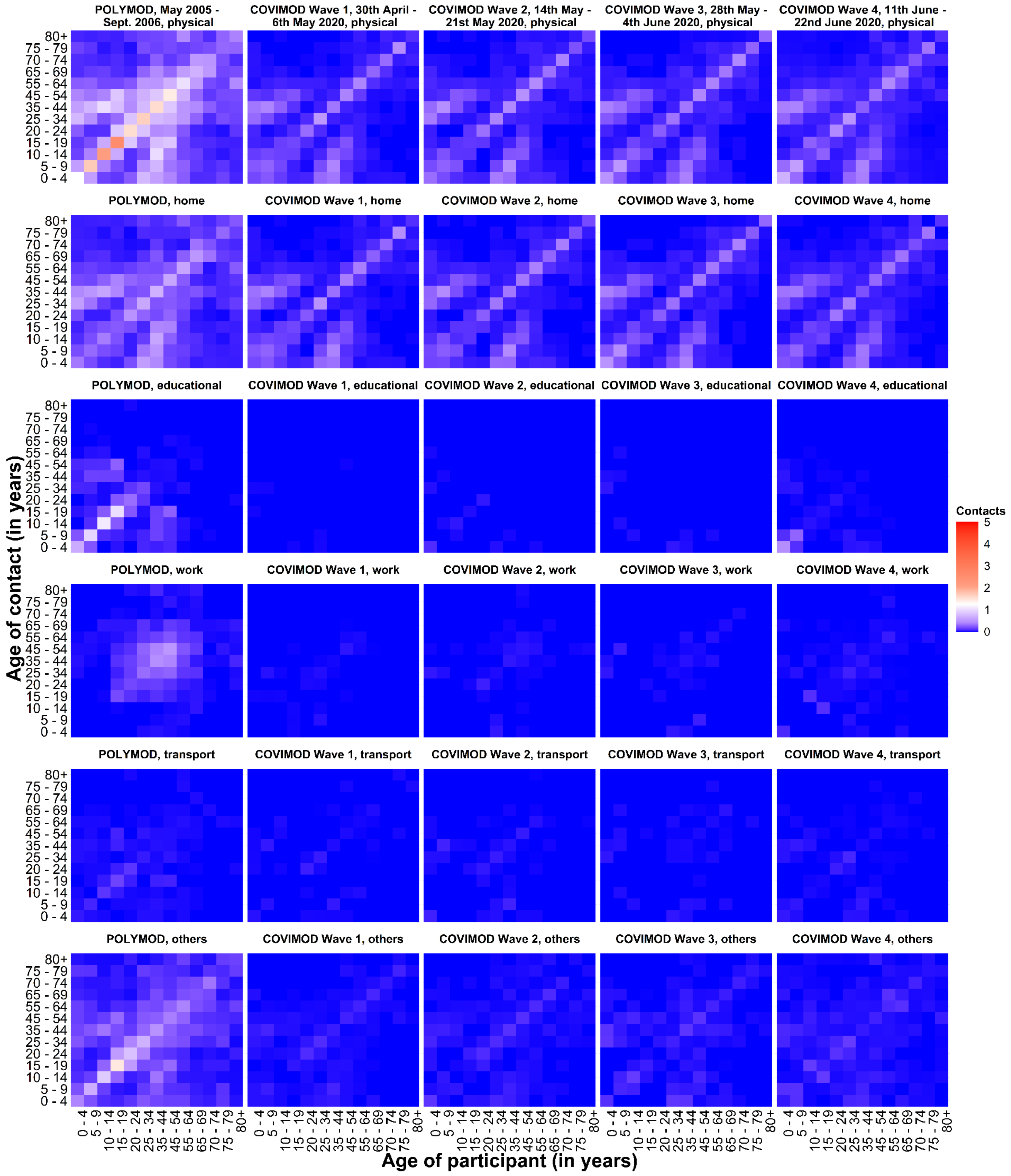

4.7. **Estimated reproduction number of SARS-CoV-2 under contact reduction measures. RKI - the percentage mean and minimum and maximum reductions in the reproduction number in the assessed time intervals, COVIMOD - the calculated percentage mean reductions in the reproduction number and the 95% confidence interval, Google and Apple mobility – the percentage mean and minimum and maximum reduction in mobility in the assessed time intervals.**

4.7.a. Figure 4.7a. Comparison of R(t) estimates obtained based on different input data. Measured effective reproduction number at the timing of COVIMOD survey Waves 1 to 4 (red), estimated reproduction number based on the reduction of social contacts at the times of COVIMOD survey Waves 1 to 4 (blue) and the reduction in mobility at the times of the COVIMOD survey Waves (yellow and green) for the unweighted analysis without group contacts. Displayed are (A) 30th April – 6th May 2020, (B) 14th May – 21st May 2020, (C) 28th May – 4th June 2020 and (D) 11th June – 22nd June 2020

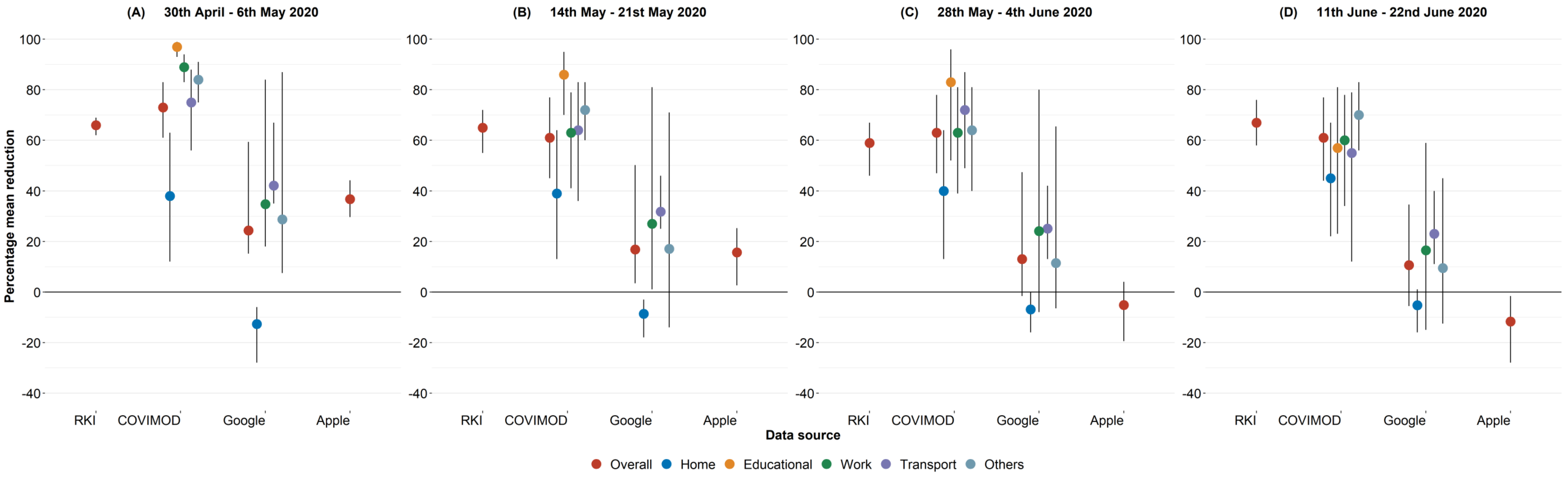


4.7.b. Figure 4.7b. Comparison of R(t) estimates obtained based on different input data. Measured effective reproduction number at the timing of COVIMOD survey Waves 1 to 4 (red), estimated reproduction number based on the reduction of social contacts at the times of COVIMOD survey Waves 1 to 4 (blue) and the reduction in mobility at the times of the COVIMOD survey Waves (yellow and green) for the unweighted analysis without group contacts.

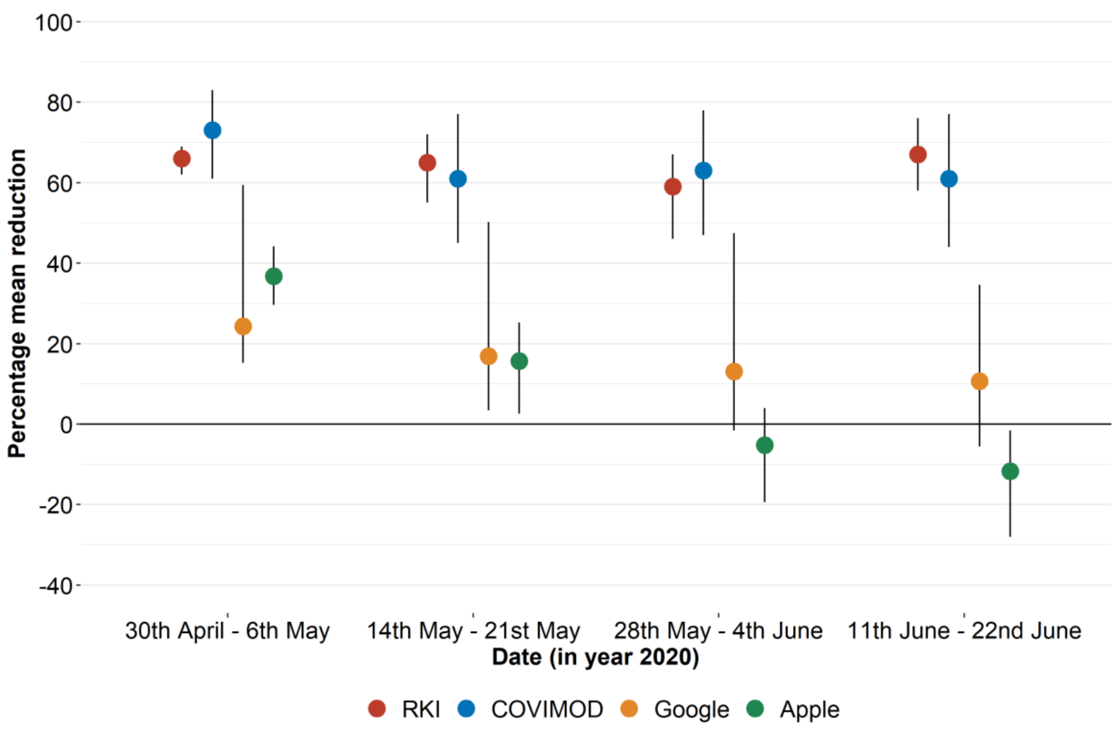
