## Supplementary File 2 for "Individual social contact data reflected SARS-CoV-2 transmission dynamics during the first wave in Germany better than population mobility data – an analysis based on the COVIMOD study"

### Questionnaire

**Demographics**
Q1 What is your age in years? YEAR/MONTH.
Which of the following describes how you think of yourself?
1. Male
2. Female
3. In another way
4. Prefer not to answer

Q3. What is your current employment status?
1. Employed full-time (34 hours or more)
2. Employed part-time (less than 34 hours)
3. Self employed
4. Unemployed but looking for a job
5. Unemployed and not looking for a job
6. Full-time parents, homemaker
7. Retired
8. Student/Pupil
9. Long-term sick or disabled

Are you the one in your household who has the highest income? [person with the largest income from employment, pensions, state benefits, investments, or other sources]
1. Yes
2. Yes, together with another household member
3. No

What is your occupation? If retired or unemployed, please indicate the category closest to your previous occupation.
In which of the following categories your occupation falls?
Legislators, senior officials, and managers

Legislators and senior officials
Corporate managers
Directors and chief executives
Production and operations department managers
Production and operations department managers in agriculture, hunting, forestry, and fishing
Production and operations department managers in manufacturing
Production and operations department managers in construction
Production and operations department managers in wholesale and retail trade
Production and operations department managers in restaurants and hotels
Production and operations department managers in transport, storage, and communications
Production and operations department managers in business services
Production and operations department managers in personal care, cleaning, and related services
Production and operations department managers not elsewhere classified
Other department managers
Finance and administration department managers
Personnel and industrial relations department managers
Sales and marketing department managers
Advertising and public relations department managers
Supply and distribution department managers
Computing services department managers
Research and development department managers
Other department managers not elsewhere classified
General managers
General managers in agriculture, hunting, forestry/ and fishing
General managers in manufacturing
General managers in construction
General managers in wholesale and retail trade
General managers of restaurants and hotels
General managers in transport, storage, and communications
General managers of business services
General managers in personal care, cleaning, and related services
General managers not elsewhere classified
Physical, mathematical, and engineering science professionals
Physicists, chemists, and related professionals
Mathematicians, statisticians, and related professionals
Computing professionals
Architects, engineers, and related professionals
Architects, town, and traffic planners
Civil engineers
Electrical engineers
Electronics and telecommunications engineers
Mechanical engineers
Chemical engineers
Mining engineers, metallurgists, and related professionals
Cartographers and surveyors
Architects, engineers, and related professionals not elsewhere classified
Life science and health professionals
Life science professionals

- Health professionals (except nursing)
- Nursing and midwifery professionals
- Teaching professionals

Other professionals
- Business professionals
- Legal professionals
- Archivists, librarians and related information professionals
- Social science and related professionals
- Writers and creative or performing artists
- Religious professionals

Physical and engineering science associate professionals
- Physical and engineering science technicians
- Computer associate professionals
- Optical and electronic equipment operators
- Ship and aircraft controllers and technicians
- Safety and quality inspectors
- Life science and health associate professionals
- Teaching associate professionals

Other associate professionals
- Finance and sales associate professionals
- Business services agents and trade brokers
- Administrative associate professionals
- Customs, tax, and related government associate professionals
- Police inspectors and detectives
- Social work associate professionals
- Artistic, entertainment and sports associate professionals
- Religious associate professionals

Clerks
- Office clerks
- Customer services clerks

Personal and protective services workers
- Travel attendants and related workers
- Housekeeping and restaurant services workers
- Personal care and related workers
- Other personal services workers
- Protective services workers
- Models, salespersons, and demonstrators

Skilled agricultural and fishery workers
- Skilled agricultural and fishery workers

Craft and related trades workers
- Extraction and building trades workers

Metal, machinery, and related trades workers
- Metal moulders, welders, sheet-metal workers, structural - metal preparers, and related trades workers
- Blacksmiths, tool-makers and related trades workers
- Machinery mechanics and fitters
- Electrical and electronic equipment mechanics and fitters

Precision, handicraft, printing, and related trades workers
- Precision workers in metal and related materials
- Potters, glass-makers and related trades workers
- Handicraft workers in wood, textile, leather and related materials

- Printing and related trades workers

Other craft and related trades workers

- Food processing and related trades workers
- Wood treaters, cabinet-makers and related trades workers
- Textile, garment, and related trades workers
- Pelt, leather, and shoemaking trades workers

Plant and machine operators and assemblers

- Plant and machine operators and assemblers

Elementary occupations

- Sales and services elementary occupations
- Agricultural, fishery and related labourers
- Labourers in mining, construction, manufacturing, and transport

Armed forces

- Armed forces

Did not work before

- Unemployed but looking for a job
- Unemployed and not looking for a job / Long-term sick or disabled
- Pupil /Student/ in full time education
- Housewife
- Retired

What is the occupation of the person with the highest income?

Legislators, senior officials, and managers

- Legislators and senior officials

Corporate managers

- Directors and chief executives

Production and operations department managers

- Production and operations department managers in agriculture, hunting, forestry,and fishing
- Production and operations department managers in manufacturing
- Production and operations department managers in construction
- Production and operations department managers in wholesale and retail trade
- Production and operations department managers in restaurants and hotels
- Production and operations department managers in transport, storage, and communications
- Production and operations department managers in business services
- Production and operations department managers in personal care, cleaning, and related services
- Production and operations department managers not elsewhere classified

Other department managers

- Finance and administration department managers
- Personnel and industrial relations department managers
- Sales and marketing department managers
- Advertising and public relations department managers
- Supply and distribution department managers
- Computing services department managers
- Research and development department managers
- Other department managers not elsewhere classified

General managers

- General managers in agriculture, hunting, forestry/ and fishing
- General managers in manufacturing
- General managers in construction
- General managers in wholesale and retail trade
- General managers of restaurants and hotels
- General managers in transport, storage, and communications
- General managers of business services
- General managers in personal care, cleaning and related services
- General managers not elsewhere classified

Physical, mathematical, and engineering science professionals
- Physicists, chemists, and related professionals
- Mathematicians, statisticians, and related professionals
- Computing professionals

Architects, engineers, and related professionals
- Architects, town, and traffic planners
- Civil engineers
- Electrical engineers
- Electronics and telecommunications engineers
- Mechanical engineers
- Chemical engineers
- Mining engineers, metallurgists, and related professionals
- Cartographers and surveyors
- Architects, engineers, and related professionals not elsewhere classified

Life science and health professionals
- Life science professionals
- Health professionals (except nursing)
- Nursing and midwifery professionals
- Teaching professionals

Other professionals
- Business professionals
- Legal professionals
- Archivists, librarians, and related information professionals
- Social science and related professionals
- Writers and creative or performing artists
- Religious professionals

Physical and engineering science associate professionals
- Physical and engineering science technicians
- Computer associate professionals
- Optical and electronic equipment operators
- Ship and aircraft controllers and technicians
- Safety and quality inspectors
- Life science and health associate professionals
- Teaching associate professionals

Other associate professionals
- Finance and sales associate professionals
- Business services agents and trade brokers
- Administrative associate professionals
- Customs, tax, and related government associate professionals
- Police inspectors and detectives
- Social work associate professionals
- Artistic, entertainment and sports associate professionals

- Religious associate professionals

Clerks
- Office clerks
- Customer services clerks

Personal and protective services workers
- Travel attendants and related workers
- Housekeeping and restaurant services workers
- Personal care and related workers
- Other personal services workers
- Protective services workers
- Models, salespersons, and demonstrators

Skilled agricultural and fishery workers
- Skilled agricultural and fishery workers

Craft and related trades workers
- Extraction and building trades workers

Metal, machinery, and related trades workers
- Metal moulders, welders, sheet-metal workers, structural - metal preparers, and related trades workers
- Blacksmiths, tool-makers and related trades workers
- Machinery mechanics and fitters
- Electrical and electronic equipment mechanics and fitters

Precision, handicraft, printing, and related trades workers
- Precision workers in metal and related materials
- Potters, glass-makers and related trades workers
- Handicraft workers in wood, textile, leather, and related materials
- Printing and related trades workers

Other craft and related trades workers
- Food processing and related trades workers
- Wood treaters, cabinet-makers and related trades workers
- Textile, garment, and related trades workers
- Pelt, leather, and shoemaking trades workers

Plant and machine operators and assemblers
- Plant and machine operators and assemblers

Elementary occupations
- Sales and services elementary occupations
- Agricultural, fishery and related labourers
- Labourers in mining, construction, manufacturing, and transport

Armed forces
- Armed forces

Did not work before
- Unemployed but looking for a job
- Unemployed and not looking for a job / Long-term sick or disabled
- Pupil /Student/ in full time education
- Housewife
- Retired

Social Grade
1 Managerial
2 Clerical
3 Manual
4 Self employed

5 Retired / Unemployed

Q15. Are you currently pregnant?

1. Yes
2. No
3. Prefer not to answer

The next question could be taken as personal, but answering it is not mandatory. However, we assure you that your answers will be kept strictly confidential and used only for research purposes.

What is the TOTAL monthly NET INCOME (after taxes) of YOUR HOUSEHOLD earned by all members of the household?

Include ALL income earned by all household members from all sources of income, e.g.: Wages/salaries, stipends, pension/benefits, dividends, rental income, alimony, child support, etc.
Select an answer

1. €0 - €500
2. €501-€750
3. €751-€1000
4. €1001-€1250
5. €1251-€1500
6. €1501-€1750
7. €1751-€2000
8. €2001-€2500
9. €2501-€3000
10. €3001-€4000
11. €4001-€5000
12. €5001-€10.000
13. €10.001 and more
14. No answer

Q20. Not including you, how many other people live in your household? By household, we mean anyone living at the same address as you, that you share a kitchen with.

None
1
2
3
4
5
6
7

8
9
10
11 or more

Q20a What is the best way to describe your household situation?
2. two or more adult non-family members
3. couple with dependent children
4. couple with only independent children
5. couple without children
6. single parent with dependent children
7. single parent with exclusively independent children
8. household with two or more families
9. other

Q21. Please write the nickname of each other person in your household.
Note that this nickname is only needed to make it easier for you to complete the survey, so please pick a nickname that will help you identify each household member later in the questionnaire. Nicknames are not visible to anyone outside of this survey.
1. NAME 1
2. NAME 2
3. NAME 3

Q23. Which of the following age groups do they fit into?

Under 1
1-4
5-9
10-14
15-19
20-24
25-34
35-44
45-54
55-64
65-69
70-74
75-79
80-84
85 years or older
Don’t know
Prefer not to answer

Q24. As far as you know, which of the following describes how [NAME] thinks of themselves?
1. Male
2. Female
3. In another way
4. Prefer not to answer
5. Don't know

Q25. What is [NAME]'s current employment status?
1. Employed full-time (34 hours or more)
2. Employed part-time (less than 34 hours)
3. Self employed
4. Unemployed but looking for a job
5. Unemployed and not looking for a job
6. Full-time parents, homemaker
7. Retired
8. Student/Pupil
9. Long-term sick or disabled

Q26. Does [NAME] attend any of the following as a pupil or student?
1. Nursery or pre-school
2. School
3. Higher education, e.g. university
5. None of the above
6. Don't know
7. Prefer not to answer

Q27. Is [NAME] currently pregnant?
1. Yes
2. No
3. Don't know
4. Prefer not to answer

Q28a Are you or any other household member in in a high-risk group, meaning you/they could have serious symptoms if you contracted Coronavirus (COVID-19)?
*High risk groups include individuals who: have had an organ transplant, undergoing cancer treatment, have blood or bone marrow cancer, have had a bone marrow or stem cell transplant in the past 6 months, are taking immunosuppressant medicine or high doses of steroids, have a severe lung condition (such as cystic fibrosis, severe asthma or severe COPD), have a condition that makes risk of getting infections higher (e.g. SCID or sickle cell), and/or are pregnant and have a serious heart condition*

*ROWS:*
0. Yourself
1. Name 1
2. Name 2
etc.

*COLUMNS:*
1. Yes
2. No
3. Don’t know
4. Prefer not to answer

**Symptoms**

SYMPTOMS

Q29. Have you, or anyone else in your household, had any of the following symptoms in the last seven days?

*ROWS:*
0. Yourself
1. Name 1
2. Name 2
etc.

*COLUMNS:*
1. Fever or high temperature
2. A cough that has lasted for at least several hours
3. Shortness of breath
4. Aches and pains, e.g.  in back, neck, shoulders or joints
5. Blocked nose
6. Sore throat
7. Feeling unusually tired
8. None of these
9. Don’t know
10. Prefer not to answer

Q30. Have you or another household member done any of the following to address these symptoms?
For each row, please select all that apply.

*ROWS:*
0. yourself
1. name 1
2. name 2 etc.

*COLUMNS:*
1. called the medical on-call service on 116 117 or used its online service.
2. called a general practitioner's office or an on-call service

3. visited a general practitioner's office or an on-call service
4. visited an outpatient center, emergency room, urgent care center, or minor injury department
5. visited an emergency room
6. visited a testing center outside of the previously mentioned facilities
7. received an admission to a hospital
8. don't know
9. none of the above
10. do not want to give any information on this

Q31. you have specified that you/NAME have/has [insert service]. When did you/has this person do/done this? If you do not know the exact answer, please provide an estimate.
ENTER DATE
1. do not know
2. i do not want to specify

Q32. Have you or another household member been tested for coronavirus (Covid-19)?
1. yes
2. no

Q33. Who has been tested for coronavirus (Covid-19)?

*ROWS:*
0. yourself
1. name 1
2. name 2 etc.

*COLUMNS:*
1. tested, and the test for coronavirus was positive
2. tested, and the test for coronavirus was negative
3. yes, but the test result is still pending
4. not tested
5. do not know
6. do not want to give any information

Q34. To the best of your knowledge, do you think you or anyone else in your household have been in direct contact with someone who has Coronavirus (Covid-19) in the last seven days, or know someone close to them who has Coronavirus (Covid-19)?

*ROWS:*
0. Yourself
1. Name 1
2. Name 2
etc.

*COLUMNS:*
1. Yes, currently infected
2. Yes, passed away
3. Yes, recovered
4. No
5. Don't know
6. Prefer not to answer

**Attitdues**
Q35. To what extent do you agree or disagree with each of the following statements?

1. Strongly agree
2. Tend to agree
3. Neither agree nor disagree
4. Tend to disagree
5. Strongly disagree
6. Don't know

1. Coronavirus would be a serious illness for me
2. I am likely to catch coronavirus
3. If I don't follow the government's advice, I might spread coronavirus to someone who is vulnerable

Q36. How effective do you think the following measures are in slowing the spread of coronavirus?

1. very effective
2. quite effective
3. not particularly effective
4. not effective at all
5. do not know

1. reduction of the number of people in contact
2. seven-day quarantine at home if one experiences mild symptoms such as a mild cough
3. quarantine at home for seven days if experiencing more severe symptoms such as a severe cough or fever
4. avoidance of busy places
5. 14-day quarantine at home if another person in your household experiences mild symptoms such as a mild cough
6. 14-day quarantine at home if another person in your household experiences more severe symptoms such as a severe cough or fever
7. closure of schools
8. closure of bars, restaurants, cinemas, etc.
9. prohibition of the use of public transportation
10. ban on travel abroad
11. ban on travel within

Q37. How confident are you that you would succeed in doing without the following things?

1. very confident
2. somewhat confident
3. not particularly confident
4. not at all confident
5. don't know

1. reduce the number of people you are in contact with
2. seven-day quarantine at home if you experience mild symptoms such as a mild cough
3. quarantine at home for seven days if you experience more severe symptoms such as a severe cough or fever
4. avoid busy places
5. 14-day quarantine at home if another person in your household experiences mild symptoms such as a mild cough
6. 14-day quarantine at home if another person in your household experiences more severe symptoms such as a severe cough or fever
7. refraining from using public transportation

Q38. To what extent do you agree or disagree with each of the following statements?

1. fully agree
2. rather agree
3. neither agree nor disagree
4. rather disagree
5. do not agree at all
6. do not know

1. other people expect me to show up at work even when I am sick
2.if I could no longer work because of the coronavirus, I would still continue to receive my salary
3. If I had to isolate myself for seven days because of the coronavirus, someone else would be available to take care of my children
4. If I had to isolate myself for seven days, this would be problematic for other people I do not know
5. i would have enough food and supplies for seven days if i had to isolate myself

**Behaviors**

Q39. You may have been asked or volunteered to take several measures for coronavirus (Covid-19).
Please think back over the past seven days and select the appropriate response for each of the actions listed below.

In the past seven days were you/was NAME asked to, …

*ROWS:*
1. Quarantine
*Quarantine is staying indoors after possible contact with an infected person. When you are in quarantine, you are allowed to leave the house, but your movement is restricted.*
2. Isolate
*Isolation is the physical separation from uninfected persons, including all household members. Isolation can occur in one's own home or in a health care facility.*
3. work from home or limit the time spent at work due to coronavirus *OR* limit the time spent at educational institution ([university or college] OR [pre-school or nurseryOR [school] due to coronavirus (Covid-19)).

*COLUMNS:*
1. yes
2. no
3. not applicable ONLY SHOW FOR THIRD STATEMENT
4. don't know
5. do not want to specify

Q40. In the past seven days, was

*ROWS:*
1. Your/NAME's workplace closed for at least one day due to coronavirus (Covid-19)?
2. Your/ NAMEs university or college OR pre-school or nursery OR school closed for at least one day?

*COLUMNS:*
1. yes
2. no
3. not applicable
4. don't know
5. do not want to specify

Q41. Have you/NAME in the past seven days

*ROWS:*
1. been in quarantine for at least one day? *Quarantine means staying indoors after possible contact with an infected person. When you are in quarantine, you are allowed to leave the house, but your movement is restricted.*
2. been isolated for at least one day? *Isolation is physical separation from uninfected persons, including all household members. Isolation can occur in the person's own home or in a health care facility.*
3. absent from work for at least one day due to coronavirus (Covid-19)?
4. absent from educational institution university or college OR pre-school or nursery OR school for at least one day due to coronavirus (Covid-19)?

*COLUMNS:*
1. yes

2. no
3. not applicable ONLY SHOW FOR STATEMENT c) 4. don't know
5. do not want to give any information

Q42. You have indicated that [you/NAME] has/have been in quarantine for at least one day. On what date did this quarantine period begin?

1. ENTER DATE
2. do not know
3. i do not want to specify

Q43. And when did/has [you/NAME] quit quarantine?

1. ENTER DATE
2. quarantine is still ongoing
3. do not know
4. i do not want to give any information on this

Q44. You indicated that [you/NAME] was/were isolated for at least one day. On what date did/does [you/they] start this isolation period?

1. ENTER DATE
2. do not know
3. i do not want to specify

Q45. Und wann haben/hat [you/NAME] die Isolation beendet?

1. ENTER DATE
2. the isolation is still going on.
3. do not know
4. i do not want to give any information

Q46. You indicated that [your/NAME's] workplace OR university or college OR pre-school or nursery OR school was closed for at least one day in the past seven days due to coronavirus (Covid-19). Please select the date on which the first closure occurred.

1. ENTER DATE
2. do not know
3. I do not want to give any information on this

Q47. And when was the closure lifted again?
Please enter in the format "DD/MM
1. ENTER DATE
2. the closure is still ongoing
3. do not know
4. i do not want to give any information

Q48. You indicated that [you/NAME] did/has not visit the workplace OR university or college OR pre-school or nursery OR school due to coronavirus (Covid-19), although no closure occurred. Please select the days within the past seven days when [you/NAME] did not visit the workplace OR university or college OR pre-school or nursery OR school due to coronavirus (Covid-19). Indicate only the days when [you/NAME] usually would have appeared there.

1. SHOW DATES FOR LAST SEVEN DAYS
2. do not know
3. i do not want to give any information

Q48A. What was the main reason for the absence (the workplace OR university or college OR pre-school or nursery OR school)?
1. Seven-day isolation due to symptoms suggestive of coronavirus.
4. Fourteen-day quarantine because another person in your household showed symptoms suggestive of coronavirus or because of contact with a known coronavirus case
5. Other illness (not coronavirus) within the household (including yourself
6. Care of a person not a member of the household in whom coronavirus (Covid-19) has been detected
7. Care of a person not a member of the household in whom coronavirus (Covid-19) has not been confirmed
8. At least one child in my household is staying home due to school closure
9. Other

Q49. You indicated that [you/NAME] was/is absent from work/work for at least one day due to coronavirus (Covid-19) or that this is not applicable. Did this have an adverse effect on your household income?

1. no, [I/NAME] was able to work from home
2. no, [I/NAME] was able to take caregiver leave
3. no, but [I/NAME] had to take annual leave
4. no, because [I/NAME] was fully compensated by the employer
5. no, because [I/NAME] was fully compensated by the state
6. yes, but [I/NAME] received partial compensation from the employer
7. yes, but [I/NAME] received partial compensation from the state
8. yes, but [I/NAME] did not receive compensation for loss of income [SINGLE CODE ONLY].
9. other (please specify)
10. do not know

11. do not want to specify

Q50. You indicated that NAME's educational institution (pre-school or nursery OR school) was closed for at least one day due to coronavirus (Covid-19). Who provided care for the child/children during this time?

1. one parent who is not employed
2. one parent who worked from home
3. a parent who works half-time
4. a parent who has taken annual leave
5. a parent who has taken caregiver leave
6. a parent who has taken leave without pay
7. siblings
8. grandparents
9. a babysitter, a childminder, an au-pair or a nanny (paid)
10. a babysitter, childminder, au-pair or nanny (unpaid)
11. a neighbor, friend, uncle or aunt
12. not necessary
13. other (please specify)
14. people at school because my child was eligible for child care at school

Q51. You indicated that NAME did not attend the educational institution (pre-school or nursery OR school OR secondary school) for at least one day due to coronavirus (Covid-19). Who provided care for the child/children during this time?

1. one parent who is not employed
2. one parent who worked from home
3. a parent who works half-time
4. a parent who has taken annual leave
5. a parent who has taken caregiver leave
6. a parent who has taken leave without pay
7. siblings
8. grandparents
9. a babysitter, a childminder, an au-pair or a nanny (paid)
10. a babysitter, childminder, au-pair or nanny (unpaid)
11. a neighbor, friend, uncle or aunt
12. not necessary [PN: Exclusive]?
13. other (please specify)
14. people at school because my child was eligible for child care at school

Q39 In the last seven days have you been in isolation or quarantine due to coronavirus (Covid-19)?
*This includes whether you have stayed at home after potential exposure to an infected case, or were asked to quarantine on return from a trip abroad, or separated yourself from people who*

*are not infected, including any household members. You can be in isolation in your house or in a health facility.*

1. Yes
2. No
3. Prefer not to say

Q41a. In the last seven days, was your/NAME'S workplace…

1. Fully open as normal
2. Partially open i.e. open some days, or for certain hours, or only open to some members of staff
3. Not applicable – I/they do not have a workplace
4. Don't know
5. Prefer not to answer

Q41b. And, in the last seven days how often did you/NAME go into your/their workplace?
1. Every day
2. Most days
3. About once or twice per week
4. No days – they/I only worked from home
5. No days – they/I did not work this week for another reason
6. Don't know
7. Prefer not to answer

Q41c. Did you/NAME go into your/their workplace yesterday?

1. Yes
2. No
3. Prefer not to answer

Q42a. In the last seven days, was your/NAME'S university or college OR pre- school or nursery OR school

1. Fully open as normal
2. Partially open i.e. open some days, or for certain hours, or only open to some children/pupils/students
3. Not applicable – it is closed for the holidays
4. Don't know
5. Prefer not to answer

Q42b. And, in the last seven days how often did you/NAME go into [university or college] OR [pre- school or nursery] OR [school]

1. Every day
2. Most days
3. About once or twice per week
4. No days – I/NAME only did the university or college OR pre- school or nursery OR school work/activities at home
5. No days – I/NAME did not attend university or college OR pre- school or nursery OR school for another reason
6. Don't know
7. Prefer not to answer

Q42c. And, did you go into university or college yesterday?

1. Yes
2. No
3. Prefer not to answer

Q52. Did you visit, or intend to visit, any of the following events or locations in the last seven days?

*ROWS:*
1. Someone else's house (including their garden)
5. A shop for non-essential items, eg a DIY shop, or a clothes or furniture shop
7. A place for sports such as a gym
9. A healthcare setting, eg hospital, GP, A&E, outpatient facility, dentist, physiotherapist, optometrist, etc
10. A hair dresser, barber, nail salon, beauty parlor or similar location
8. Outside, for example in a park, on the street or in the countryside

*COLUMNS:*
1. Yes, I visited this event or location
2. I intended to visit but it was cancelled because of the coronavirus (covid-19) epidemic
3. I intended to visit but chose not to go because of the coronavirus (covid-19) epidemic
4. I intended to visit but I had to cancel/it was cancelled for reasons unrelated to the coronavirus (covid-19) epidemic
5. No, I did not visit or intend to visit this event or location

Q53. You said that you visited / intended to visit [a INSERT EVENT OR LOCATION] but it was cancelled because of coronavirus / but chose not to go because of coronavirus / but you had to cancel or it was cancelled for reasons unrelated to coronavirus]. How many times did that happen in the last seven days?
1. INSERT NUMBER OF TIMES

2. Don’t know

**Individual preventive measures**

Q54. Did you use a face mask yesterday?
1. Yes
2. No

Q55. How long did you wear this mouth-nose mask in total?
ENTER NUMBER OF HOURS
ENTER NUMBER OF MINUTES

Q56. Where did you use your face mask?
1. Everywhere outside my house
2. When walking on the street
3. When cycling
4. On public transport
5. In supermarkets/shops
6. In cinema/bar/restaurant
8. At home
9. At work/school/college/university
7. Other (please specify)

Q57. How often did you wash your hands in the last three hours?
NUMBER OF ENTRIES

Q58. How often did you use hand sanitizer in the last three hours?
NUMBER OF ENTRIES

Q59. Did you travel on any public transport yesterday?
1. No
2. Train/tube
3. Bus/tram
4. Taxi, Uber, or similar ride-hailing app
5. Aeroplane

Q60. And how much time in total did you spend on the journey?
INSERT NUMBER OF HOURS

INSERT NUMBER OF MINUTES

**Contact survey**

We will now ask you to remember who [you have /NAME has] been in contact with yesterday, between 5am yesterday and 5am today.

These questions are voluntary but they are really important in helping us understand the spread of COVID-19 and the impact of different public health interventions. It will not be possible to identify you or any member of your household in the published findings.

We are only interested in direct contacts, which are **people who [you/NAME] met in person** and with whom [you/NAME] exchanged at least a few words, or with whom [you/NAME] had physical contact (e.g. a handshake, embracing, kissing, contact sports).
**Note that if [you/NAME] only spoke to someone over the phone or internet, they should not be included in this section.**

Q62. Which of the following people did **[you/NAME]** have direct contact with in person, between 5am yesterday and 5am today, in person?

*We are only interested in direct contacts, which are people who you met in person and with whom you exchanged at least a few words, or with whom you had physical contact (e.g. a handshake, embracing, kissing, contact sports).*
***Note that if you only spoke to someone over the phone or internet, they should not be included in this section.***

*COLUMNS:*
1. Yes
2. No
999_ You, the person completing the survey

CONTACT2
Q63. And what other people, **outside of your household,** did **[you/NAME]** have direct contact with in person, between 5am yesterday and 5am today? This could include friends, family, work colleagues, or people **[you/NAME]** spoke to in shops and so on.

Please write the nickname of each person **[you/NAME]** had direct contact with below. Note that this nickname is only needed to make it easier for you to complete the survey, so please pick a nickname that is easy to remember. Your individual response will not be shared with anyone outside of this survey.

*We are only interested in people who you met in person and* ***with whom [you/NAME] exchanged at least a few words, or with whom [you/NAME] had physical contact*** *(e.g. a handshake, embracing, contact sports)***.**

It is easiest list names in chronological order, e.g. After [I/NAME] had breakfast at home, [I/NAME] went to work where [I/NAME] met with Jack, Deborah, and two clients. On the way back home, [I/NAME] chatted with the shop assistant at the petrol station (give a nickname like “shop assistant”). When [I/NAME] returned home, [I/NAME] accepted a package from the delivery person, and [I/NAME] spoke to a friend, Fatima, in the garden. Etc.

**Please do not list yourself, anyone that you listed as being a part of your household, or anyone who you only spoke to over the phone or internet.**

*PN: SHOW IFQ62 for any coded 1  You have already indicated contact with the following household members, add additional contacts in the textboxes below:*

LIST HOUSEHOLD MEMBERS FROM Q21
PLEASE LIST ALL OTHER CONTACTS YOU HAVE OUTSIDE OF YOUR HOUSEHOLDE, FOR EXAMPLE, J, D, CLIENT 1, CLIENT 2, SHOP ASSISTANT, DELIVERY PERSON, FRIEND F

Q66. Which of the following age groups does NAME fit into? Please give an estimate if you are not sure

1. Under 1
2. 1-4
3. 5-9
4. 10-14
5. 15-19
6. 20-24
7. 25-34
8. 35-44
9. 45-54
10. 55-64
11. 65-69
12. 70-74
13. 75-79
14. 80-84
15. 85+
16. Don’t know
17. Prefer not to answer

Q67. As far as you know, which of the following describes how [NAME] thinks of themselves?

1. _1 Male [KEEP]
2. _2 Female [KEEP]
3. _3 In another way [KEEP]
4. _4 Prefer not to answer [KEEP]
5. _5 Don’t know
6. _5 Weiß nicht

Q68. What is [NAME]'s relationship to **[you/NAME]**?
1. They are a family member who is not in my household 2. They are someone [I work/NAME works] with
3. They are someone [I go to school, college or university with/NAME goes to school, college or university with]
4. a boyfriend/girlfriend
5. other
6. i do not want to give any information

Q69. Before the coronavirus epidemic started, how often did **[you/NAME]** usually have direct contact with [*NAME*]?
A direct contact is when you **meet with this person in person** and when you exchange at least a few words, or when you have physical contact (e.g. handshake, embracing, kissing, contact sports).
**Please do not include times that you speak to them over the phone or internet.**

1. Every day or almost every day
2. About once or twice a week
3. Every 2-3 weeks
4. About once per month
5. Less often than once per month
6. Never met them before
7. Prefer not to answer

Q70. When [you/NAME] had/had an immediate contact with [NAME] yesterday, [you/NAME] had/had a ...

1. physical contact (any kind of skin contact) (such as shaking hands, hugging, or kissing)?
2. non-physical contact (you did not touch the person)?
3. I do not want to give any information on this

Q71. And where did [you/NAME] have direct contact with [NAME]?

1. At home (including at your door, in your garden, and within entrances to your home such as stairways, lifts, and corridors)
2. At someone else's house
3. At work
4. At a place of worship
5. On any form of transport
6. At university, school, pre-school, or nursery
7. At a shop for essentials, eg a supermarket, grocery store, market, pharmacist, or bicycle shop
8. At a shop for non-essential items, eg a gardening center, or a clothes, electronics, furniture, or DIY shop
9. At a place of entertainment such as a restaurant, bar, cinema

10. At a place for sports such as a gym or sports club/match [PN. DO NOT SHOW IN BE AND NL]
11. Outside, for example in a park, on the street or in the countryside
14. In a healthcare setting, eg hospital, GP, A&E, outpatient facility, dentist, physiotherapist, optometrist, etc
15. At a hair dresser, barber, nail salon, beauty parlor or similar location
12. Somewhere else (please specify)

Q72. Please estimate the total amount of time [you/NAME] spent with [NAME] in person yesterday.

Q73 Was the time [you/NAME] spent with [NAME] yesterday inside or outside?
Please tick all that apply
Inside
Outside

Q74 We do ask you to individually include every contact you had, but if you were unable to include every single contact (for instance, because you work in a shop and have a large number of contacts in a day), please could you indicate this?

1. I individually included every person I had contact with.
2. I did not individually include every person I had contact with.
3. I did not have any contacts (PN: ONLY SHOW IF NO CONTACTS LISTED AT Q63 I.E. ALL TEXT BOXES WERE LEFT EMPTY AND NO CONTACTS ARE CODED AS 1 AT Q62)

*Q75 Approximately how many people have you had contact with that you did not specify individually? Please provide as accurate an estimate as possible, including age and environment.*

*For the purpose of this question, we are only interested in people with whom there was direct contact and with whom you exchanged at least a few words OR with whom you had physical contact (e.g., shaking hands, hugging, contact sports).*

*Please indicate the number of contacts for each age group and setting.*

ROWS
Under 18 years
18 to 64 years
65 years and older

COLUMNS
At the workplace
At school or other educational institution
Elsewhere

*Q76 And approximately how many people did you have <b>**physical contact**</b> with that you did not specify individually? Please provide as accurate an estimate as possible, including age and environment*
*For the purpose of this question, we are only interested in people with whom there was direct contact* ***AND with whom you had physical contact (e.g., shaking hands, hugging, playing contact sports).***

*Please indicate the number of contacts for each age group and setting.*

ROWS
Under 18 years
18 to 64 years
65 years and older

COLUMNS
At the workplace
At school or other educational institution
Elsewhere
