## Supplementary File 3 for "Individual social contact data reflected SARS-CoV-2 transmission dynamics during the first wave in Germany better than population mobility data – an analysis based on the COVIMOD study"

### How additional contacts were considered in COVIMOD

In some rare cases, participants did not enter one contact per line in the contact diary, but instead entered “5 colleagues” or “grandparents”. In this case, if the number of additional contacts is known (e.g. “3 colleagues”), we considered 3 contacts (in the same age group/contact setting/physical contact as the original contact) instead of 1 contact.

> **Example**
> Consider a hypothetical participant, J, who reported “3 colleagues” as one contact in the age group “30-40 years”. J had contact a non-skin-to-skin contact with them at work. In this case, we considered that participant J had 3 contacts, each contact aged “30-40 years”, met at work and non-physical.

If the number of additional contacts is unknown, e.g. the participant only stated “friends” or “grandparents”, we assumed the number of contacts provided in Table 1.

**Table 1.** Assumed number of additional contacts**.**

| | |
|---|---|
| Work colleagues/colleagues | Assumed 10 |
| Friends | Assumed 5 |
| Patients | Assumed 20 |
| School class | Assumed 15 |
| Physician's assistants | Assumed 3 |
| Residents of the nursing home | Assumed 20 |
| Persons cared for at work | Assumed 20 |
| Kindergarten | Assumed 15 |
| Clients | Assumed 30 |
| Grandchildren | Assumed 3 |
| Supermarket | Assumed 5 |
| Suppliers | Assumed 5 |
| Children | Assumed 3 |
| Persons from my rented apartment | Assumed 3 |
| Bank employee | Assume 2 |
